# Direct and Indirect Genetic Effects of Parental Liabilities to Mental Health Conditions and Related Traits on Children’s Behavioural Difficulties: A Multi-Cohort Study

**DOI:** 10.64898/2026.02.10.26345985

**Authors:** Li Tian, Mina Shahisavandi, Adrian Dahl Askelund, René Pool, Ellen Verhoef, Svenja Müller, Theresa Rohm, Marius Lahti-Pulkkinen, Josef Frank, Eric Zillich, Charlotte Pahnke, Alicia Schowe, Johanna Tuhkanen, Lidia Fortaner-Uyà, Benedetta Vai, Francesco Benedetti, Andreas J. Forstner, Darina Czamara, Christian Kandler, Maria Gilles, Stephanie H Witt, Lianne de Vries, Dorret I. Boomsma, Meike Bartels, Katri Räikkönen, Helga Ask, Ole A. Andreassen, Jean-Baptiste Pingault, Beate St Pourcain, Charlotte A. M. Cecil, Alexandra K. S. Havdahl, Alexander Neumann, Jari Lahti

## Abstract

**Background:** Parental genetics matters for children’s behavioural difficulties, but the extent to which this is due to direct genetic transmission versus environmentally mediated indirect genetic effects remains unclear.

**Methods:** We studied eight European birth cohorts with over 33,000 family-based trio samples. We analysed polygenic scores (PGSs) for 13 mental health and neurodevelopmental conditions and their composite indices (PC1 and mean) representing general neuropsychiatric liabilities, as well as educational attainment (EA) and alcohol and cigarette use, from children (PGSc), mothers (PGSm), and fathers. Child internalising, externalising, and total difficulties reported by mothers and/or fathers were examined at preschool and school ages. We then conducted multivariate meta-analyses to combine cohort-level results.

**Findings:** We observed several direct genetic effects on externalising difficulties, while indirect genetic influences were mainly identified for internalising difficulties. Specifically, child PGSs for attention-deficit/hyperactivity disorder (ADHD) and EA predicted higher and lower levels, respectively, of child externalising and total difficulties (all *p*_FDR_<0·001; for school-aged externalising difficulties, PGSc-ADHD: β=0·121 [95% CI 0·091 to 0·151], *p*_FDR_<0·0001; PGSc-EA: β=−0·095 [95% CI −0·127 to −0·063], *p*_FDR_<0·0001), whereas maternal PGSs for major depressive disorder (MDD) and general neuropsychiatric liabilities were associated with internalising and total difficulties across parental raters and child ages (all *p*_FDR_<0·05; for school-aged internalising difficulties, PGSm-MDD: β=0·049 [95% CI 0·017 to 0·081], *p*_FDR_=0·016; PGSm-PC1: β=0·056 [95% CI 0·022 to 0·091], *p*_FDR_=0·011). No statistically significant effects from paternal PGSs were identified.

**Interpretation:** In this multi-cohort study, findings across multiple traits, raters, and ages supported several direct genetic effects of ADHD and EA on child externalising difficulties and indirect genetic effects on internalising difficulties, especially maternal depression and general neuropsychiatric liabilities. These suggest that child internalising difficulties are not solely driven by direct genetic transmission. More comprehensive research is needed to better understand the mechanisms involved, and ultimately how to ameliorate child behavioural difficulties.

**Funding:** EU, ERC, RCN, RCF, UKRI, SERI, DFG

**Research in context:** *Evidence before this study:* Indirect genetic effects (IGEs) refer to the influence of parental genotypes on offspring outcomes beyond direct genetic effects (DGEs), for example via environmental pathways. While IGEs on offspring cognitive traits are well-established for educational attainment, evidence for IGEs of parental liabilities to mental health and neurodevelopmental conditions remains limited. To assess the current state of evidence, we conducted a systematic search of published studies applying trio-based polygenic score (PGS) designs to child and adolescent mental health outcomes. We identified 141 primary studies in MEDLINE, Embase, PsycInfo, and Web of Science, by 6 March 2025, after removing duplicates; following screening, 12 studies met inclusion criteria (see supplement for a full description including results). Ten out of the 12 studies focused on externalising outcomes, with little or inconsistent support for IGEs. When observed, IGEs were mainly driven by maternal liabilities to autism, educational attainment, and cognitive performance on child outcomes. The current evidence was too limited and heterogeneous to synthesize findings quantitatively, therefore a qualitative synthesis was conducted. Many studies were statistically underpowered, and the observed IGEs were in all cases sample-specific. There were no published multi-cohort studies.

*Added value of this study:* We integrated information across over 33,000 mother-father-child trios from eight European cohorts, investigating 18 PGSs from parents and children, using maternal and paternal ratings of offspring’s internalising, externalising, and total difficulties as outcomes at both preschool and school age. We mainly observed DGEs on externalising difficulties, consistent with previous studies. Some evidence of IGEs was found for internalising and total difficulties. IGEs were often found to be maternally driven, with the most robust evidence across ages and raters emerging for maternal depression and general neuropsychiatric liabilities.

*Implications of all the available evidence:* The current evidence suggests that children’s behavioural difficulties, especially internalising difficulties, may be partly driven by the environment shaped by maternal neuropsychiatric liabilities. Ours and previous findings highlight a pressing need for more comprehensive studies across different cohorts, raters, outcomes, and time points to understand the true extent of IGEs in the intergenerational transmission of mental health.

## Introduction

Globally, 6–12% of children experience neurodevelopmental and mental health conditions that may persist throughout their lifespan.^1^ Since the risk for developing these conditions increases steadily across childhood and early adolescence,^2^ this developmental period may represent a window of opportunity for early intervention. Therefore, identifying targetable and malleable risk and protective factors for behavioral difficulties early in life is a major public health priority.

Numerous studies have demonstrated that neurodevelopmental and mental health conditions run within families.^3^ For childhood internalising (e.g., anxiety and depression) and externalising difficulties (e.g., hyperactivity, impulsivity, and conduct problems), twin-based heritability estimates have typically been 0·4–0·5, comparable to many major adulthood mental health conditions.^4^ A core aim of intergenerational psychiatry is to disentangle the roles of genes and environments in the transmission of mental health and neurodevelopmental conditions across generations.^5^ This transmission can occur via two primary pathways: 1) inheritance of genetic predisposition to a psychiatric condition affecting offspring’s development, also termed direct genetic effects (DGEs) and 2) indirect genetic effects (IGEs) going via for instance parents’ own lifestyles and parenting behaviours due to their psychiatric liabilities, which can then affect offspring’s environment (“genetic nurture”).^6^ In addition to genetic nurture, IGEs may also stem from other mechanisms including intrauterine or hormonal pathways at the fetal stage, broader forms of gene-environment correlations such as those arising from social deprivation or neighbourhood selection, assortative mating, and cross-generation population stratification processes.^7^ To understand the role of nature and nurture in the intergenerational transmission of psychiatric conditions, family-based designs such as children-of-twins and adoption studies have traditionally been used.^8^ The advent of genotyped parent-offspring trios^5^ offers another way to clearly delineate nature and aspects of nurture, conceptualised as DGEs and IGEs, respectively (figure 1).

**Figure 1.**
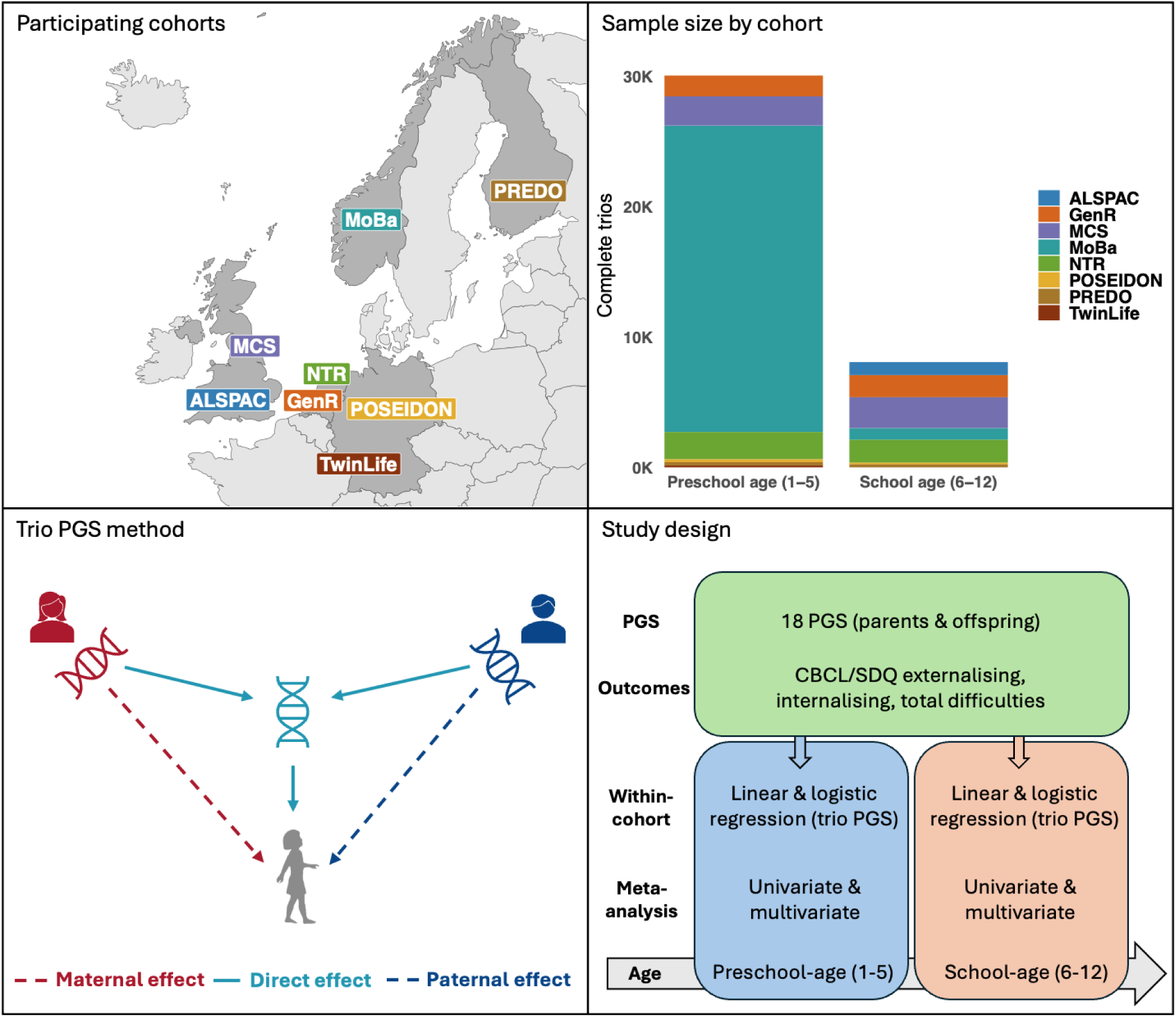
Overview of the participating cohorts, trio PGS method, and study design.

Recent within-family and multi-generation research using polygenic scores (PGSs) to examine DGEs and IGEs on child behavioural outcomes has yielded mixed findings. Some studies found evidence of primarily DGEs,^9–15^ such as on attention-deficit/hyperactivity disorder (ADHD)-related traits,^9,11,15^ internalising difficulties,^12^ aggression,^13^ conduct problems,^14^ and autism.^10^ Meanwhile, other studies showed evidence of IGEs on ADHD-related traits,^16–18^ and externalising,^20^ internalizing,^19,21^ or total^22^ behavioural difficulties. These inconsistencies may be due to limited sample size, single cohorts, as well as narrow selection of predictors and outcomes. Additionally, most previous studies only focused on maternal ratings of child behaviours, which may cause inflated IGEs and hence misinterpretation of findings.

To overcome the above limitations, we established the “characterizing intergenerational genetic transmission of mental health (TRITONE)” consortium (figure 1), joined by eight birth cohorts from five European countries, with mother- and/or father-reported offspring outcomes at preschool- and school-ages, reaching maximally ∼33,000 complete genotyped trio samples. Our goal was to examine DGEs and IGEs of liabilities to mental health conditions and related traits on children’s behavioural difficulties.

## Materials and information

### Patient and public involvement

To inform interpretation and dissemination of our research, we consulted four parents with lived experience, recruited through Mental Health Norway. Participants highlighted the sensitivity of the research topic and the importance of careful communication to avoid deterministic interpretations and unintended stigma. Their feedback informs our emphasis on group-level risk rather than individual-level conclusions, including in lay summaries for the public.

### Participating studies

Eight European cohorts in the TRITONE consortium participated in this meta-analysis: the *Avon Longitudinal Study of Parents and Children* (ALSPAC, United Kingdom), the *Generation R Study* (GenR, Netherlands), the *Millennium Cohort Study* (MCS, United Kingdom), the *Norwegian Mother, Father and Child Cohort study* (MoBa, Norway), the *Netherlands Twin Register* (NTR, Netherlands), the *Pre-, Peri-, and Postnatal Stress: Epigenetic Impact on Depression study* (POSEIDON, Germany), the *Prediction and Prevention of Preeclampsia and Intrauterine Growth Restriction study* (PREDO, Finland), and the *TwinLife study* (TwinLife, Germany). We included complete parent-offspring trios with available genotype and offspring outcome data. For siblings, only one was included at random, or clustering was accounted for in the models. Samples were restricted to genetically inferred European-associated ancestry because the PGSs were trained predominantly in European-associated ancestry samples. All cohorts performed statistical analyses according to a predefined analysis plan. All studies were approved by their institutional ethics review committees and all participants provided written informed consent (see supplement).

### Genotype data

Each cohort genotyped their DNA samples and processed their genotype data internally using single nucleotide polymorphism (SNP) arrays. Quality control (QC) meeting the minimum imputation criteria was performed in all cohorts.

### PGS predictors and calculation

We used SNP weights based on the summary statistics available from the most recently published GWASs, to calculate PGSs classified as 1) general neuropsychiatric liabilities, represented by the cross-disorder genetic liability (CDG) as well as the mean (Psych-Mean) and the first principal component (Psych-PC1) scores aggregating PGSs for CDG along with 12 psychiatric and neurodevelopmental conditions (listed in groups 2–5 below) and insomnia, which is a symptom closely associated with these conditions; 2) emotional conditions and traits, including anxiety disorders (ANX), major depressive disorder (MDD), neuroticism (NEUROT), postpartum depressive disorder (PPD), and post-traumatic stress disorder (PTSD); 3) compulsive conditions, including anorexia nervosa (AN) and obsessive-compulsive disorder (OCD); 4) neurodevelopmental conditions, including ADHD and autism spectrum disorder (ASD); 5) psychotic conditions, including bipolar disorder (BD) and schizophrenia (SCZ); 6) substance use, including daily consumption of alcohol (ALC) and cigarettes (CIG); and 7) other associated traits, including educational attainment (EA) and insomnia (INSOM).

Leave-one-out summary statistics were used for BD, EA, and PPD in MoBa and ALC, CIG, and NEUROT in NTR since these cohorts participated in the original GWASs. Moreover, NTR was excluded from our analysis of ANX.

Before calculating the PGSs, QC of GWAS summary statistics was performed by removing the SNPs with standard deviations (SD) of allele frequencies either above or below an expected value as inferred from the UK Biobank reference panel. SNP weights were computed using LDpred2-auto. PGSs for each mother (PGSm), father (PGSf), and child (PGSc) were then calculated by Plink software (v1.9). All PGSs were standardized to a mean of 0 and an SD of 1 within each cohort. Outliers were winsorized according to the +/− 3*interquartile range (IQR) criterion to reduce false positives when analysing human population samples.

### Behavioural outcome measures and covariates

The Child Behaviour Checklist (CBCL) or Strengths and Difficulties Questionnaire (SDQ) were used to measure child behavioural difficulties. All cohorts collected mother-reported behavioural measures. Additionally, GenR, NTR, PREDO, and TwinLife collected father-reported measures.

Summative CBCL T-scores or SDQ raw scores for three main scales, i.e., externalising, internalising, and total difficulties were calculated. By applying a cut-off value of 60 for the CBCL T-score or 14 for the SDQ raw score for total difficulties, a dichotomized variable indicating a clinically relevant psychopathological risk was also calculated.

The child’s age and sex and genetic ancestry PC1–PC10 were included as covariates. Additionally, we adjusted for maternal educational level and maternal age at childbirth to improve the specificity of potential IGEs and control for wider dynastic effects beyond nuclear families. Both outcomes and covariates were standardized and winsorized as described above (table 1).

**Table 1.**
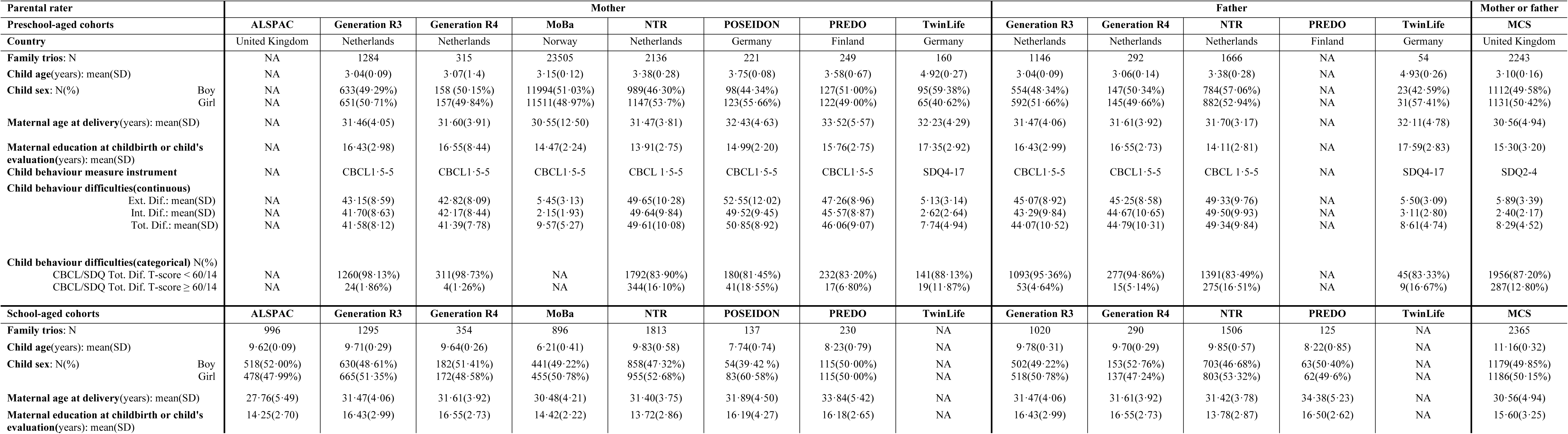
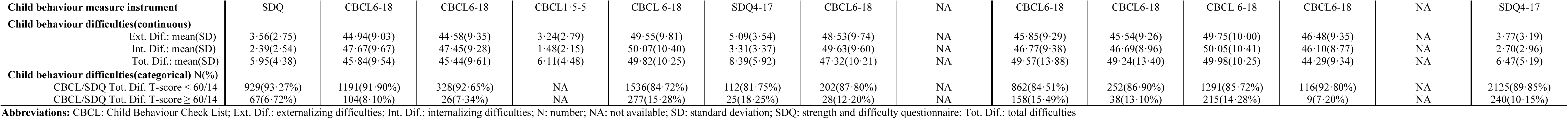
Demographics and child behavioural characteristics in the participating study cohorts.

### Cohort-level statistical models and analyses

All cohorts used the same scripts in R for statistical analyses. Linear and logistic regression models were set up for individual PGS or trio-PGSs (leveraging PGSc, PGSm, and PGSf simultaneously) on mother- or father-reported child behavioural outcomes. Multicollinearity among predictors and covariates and Pearson’s correlation coefficients among the trio-PGSs were examined (table S2).

### Meta-analyses

We applied a random-effect and inverse variance-based multivariate meta-analysis (MVMA) using the rma.mv function in the *metafor* package in R, pooling summary statistical results across cohorts and raters for each age (N=33,202 preschool age/10,854 school age). To account for dependent observations between ratings from different informants for the same child, we modelled this dependency using a variance-covariance matrix derived from Pearson’s correlation coefficients between mother- and father-reported outcomes. For verification of MVMA results, we additionally conducted univariate meta-analyses (UVMA) on parental ratings separately, excluding MCS due to its mixed parental rating (mother-rating: N=27,828 preschool-age/5,669 school-age; father-rating: N=3,131 preschool-age/2,820 school-age). Standardized beta coefficients and confidence intervals (CI) were used to evaluate overall effects. For logistic regressions, cohorts having a sample size <10 in the high-risk group were excluded in meta-analyses. All reported *p*-values are two-sided and were corrected for multiple comparisons using the False Discovery Rate (FDR) among each set of the 18 PGSs (*i.e.*, 18 tests) for child, mother, and father, respectively. The I^2^ statistic was used to evaluate cohort-level heterogeneity (tables S3-S28).

### Sensitivity analyses

Since genotyping for fathers was different from mothers and offspring in ALSPAC, this cohort was excluded from the MVMA for trio-PGSs models in the school-aged group for sensitivity analysis.

## Results

### Cohort characteristics

Table 1 presents descriptive statistics including family demographics and child behavioural outcomes of the participating cohorts. Altogether, 33,202 mother-father-child trios in the preschool-aged group and 10,854 in the school-aged group were included in meta-analyses, each including 7 cohorts. Children were 1·5–5 years old (mean=3·53) in the preschool-aged group and 6–12 years old (mean=9·14) in the school-aged group. Female children represented 40·6–60·6% of the samples.

PGSc, PGSm, and PGSf were calculated, and their correlations were analysed in each cohort (table S1). Significant correlations between PGSm and PGSf for some traits, especially EA, were observed in several cohorts (figure S1; table S2).

### Associations of each individual PGS with child behavioural difficulties

We first examined associations of each PGS individually with child outcomes reported by both mothers and fathers in MVMA (tables S3–S6) as well as by either mothers or fathers separately in UVMA (tables S11–S18). These analyses indicated numerous associations of either children’s or parental PGSs with child behavioural difficulties, when unadjusted for the other family members’ PGSs. The largest effects came from both PGSc and PGSm for general neuropsychiatric liabilities as well as PGSc for ADHD and EA at both preschool- and school-ages (all |β|>0·035; all *p*_FDR_<0·001; tables S3–S4).

### Evidence of direct and indirect genetic effects on child behaviours

To partition DGEs from parental IGEs, we regressed trio-PGSs on child behavioural difficulties (figures 2–4, tables S7–S10, and figures S2–S27 for MVMA; tables S19–S26 for UVMA).

**Figure 2.**
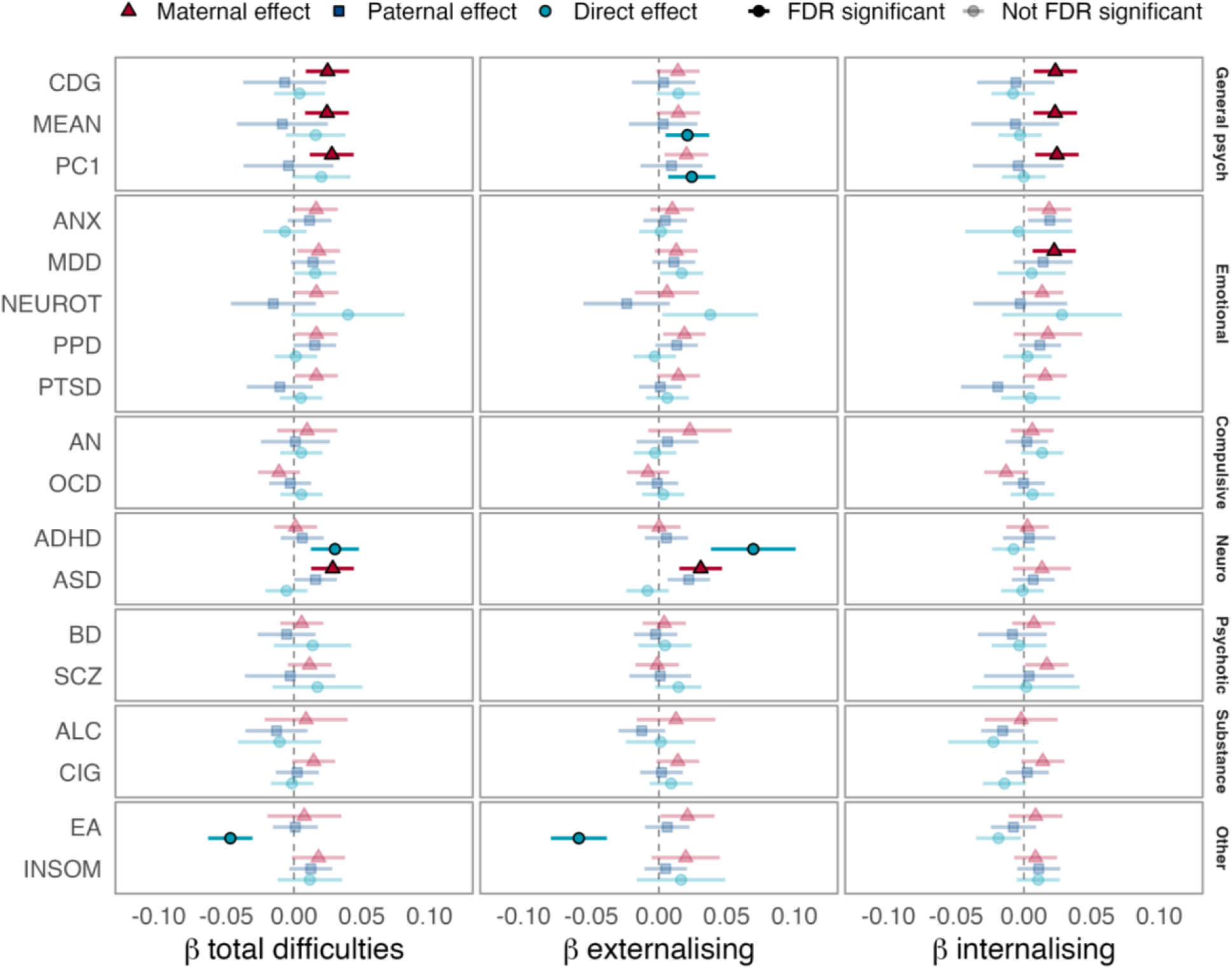
Direct and indirect genetic effects of liability to mental health conditions and related traits on child behavioural outcomes at preschool age (N=33,202, 7 cohorts). GenR, MCS, MoBa, NTR, POSEIDON, PREDO, and TwinLife were included in the analysis. Effect sizes and 95% CIs are shown. PGSs from children, mothers, and fathers were entered simultaneously as predictors of child total, externalising, and internalising difficulties, using mother- or father-reported CBCL- or SDQ-scores. PGSs are clustered within categories corresponding to 1) general neuropsychiatric liabilities, including cross-disorder genetic psychiatric liability (CDG), Psych-Mean (MEAN) of neuropsychiatric PGSs and their first principal component Psych-PC1 (PC1); 2) emotional conditions and traits, including anxiety disorders (ANX), major depressive disorder (MDD), neuroticism (NEUROT), postpartum depressive disorder (PPD), and post-traumatic stress disorder (PTSD); 3) compulsive conditions, including anorexia nervosa (AN) and obsessive-compulsive disorder (OCD); 4) neurodevelopmental conditions, including attention-deficit/hyperactivity disorder (ADHD) and autism spectrum disorder (ASD); 5) psychotic conditions, including bipolar disorder (BD) and schizophrenia (SCZ); 6) substance use, including daily consumption of alcohol (ALC) and cigarettes (CIG); and 7) other associated traits, including educational attainment (EA) and insomnia (INSOM).

**Figure 3.**
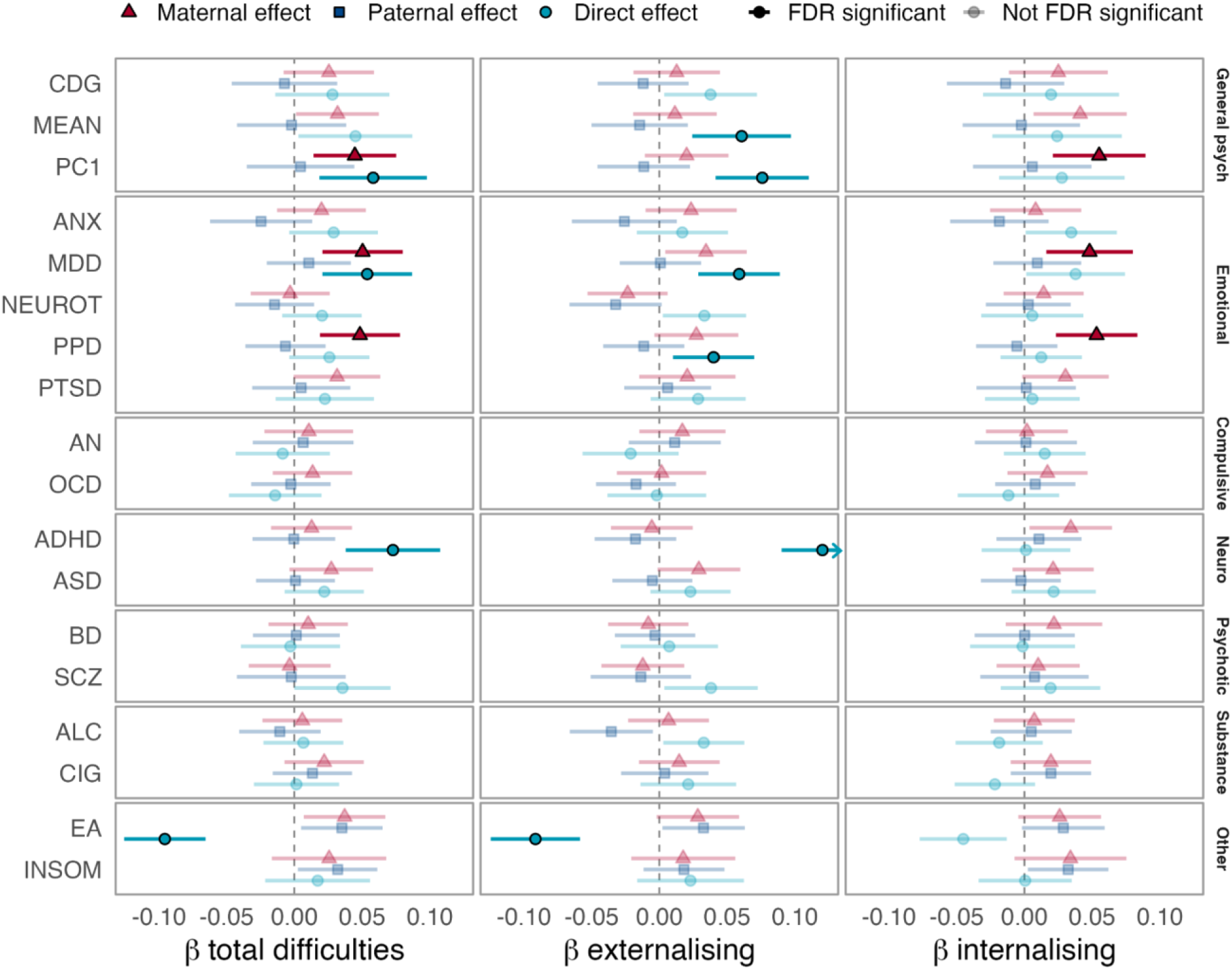
Direct and indirect genetic effects of liability to mental health conditions and related traits on child behavioural outcomes at school age (N=10,854, 7 cohorts). ALSPAC, GenR, MCS, MoBa, NTR, POSEIDON, and PREDO were included in the analysis. Effect sizes and 95% CIs are shown. PGSs from children, mothers, and fathers were entered simultaneously as predictors of child total, externalising, and internalising difficulties, using mother- or father-reported CBCL- or SDQ-scores. PGSs are clustered within categories corresponding to 1) general neuropsychiatric liabilities, including cross-disorder genetic liability (CDG), Psych-Mean (MEAN) of neuropsychiatric PGSs and their first principal component Psych-PC1 (PC1); 2) emotional conditions and traits, including anxiety disorders (ANX), major depressive disorder (MDD), neuroticism (NEUROT), postpartum depressive disorder (PPD), and post-traumatic stress disorder (PTSD); 3) compulsive conditions, including anorexia nervosa (AN) and obsessive-compulsive disorder (OCD); 4) neurodevelopmental conditions, including attention-deficit/hyperactivity disorder (ADHD) and autism spectrum disorder (ASD); 5) psychotic conditions, including bipolar disorder (BD) and schizophrenia (SCZ); 6) substance use, including daily consumption of alcohol (ALC) and cigarettes (CIG); and 7) other associated traits, including educational attainment (EA) and insomnia (INSOM).

**Figure 4.**
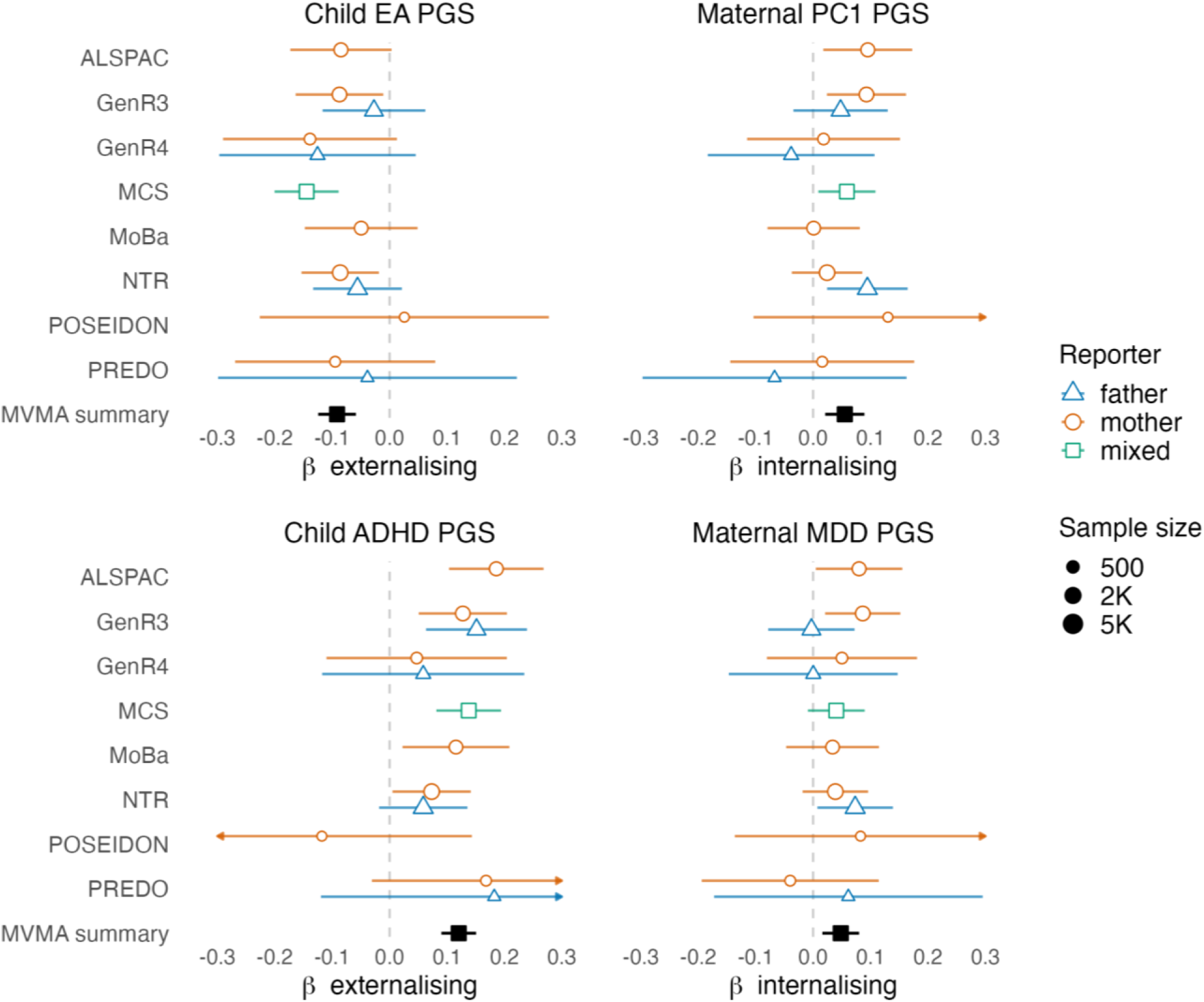
The most robust direct and indirect genetic effects on child externalising and internalising difficulties at school age, by cohort and rater (N=10,854, 7 cohorts). Effect sizes and 95% CIs are shown. PGSs from children, mothers, and fathers were entered simultaneously as predictors of child externalising or internalising difficulties, using mother- or father-reported (or mixed) CBCL- or SDQ-scores. PGS = polygenic score; ADHD = attention-deficit/hyperactivity disorder; MDD = major depressive disorder; EA = educational attainment; PC1 = first principal component indexing general neuropsychiatric liability.

#### General neuropsychiatric genetic liabilities

We first examined aggregated liability to neuropsychiatric conditions represented by the CDG, Psych-PC1, and Psych-Mean PGS scores. In preschool-aged children (figure 2; table S7), PGSm-CDG but not PGSc-CDG was associated with both internalising (β=0·023 [95% CI 0·007 to 0·039]; *p*_FDR_=0·027) and total difficulties (β=0·025 [95% CI 0·009 to 0·041]; *p*_FDR_=0·014). Maternal Psych-Mean and Psych-PC1 scores behaved similarly (Psych-Mean on internalising: β=0·023 [95% CI 0·007 to 0·039]; *p*_FDR_=0·027; Psych-Mean on total: β=0·024 [95% CI 0·008 to 0·040]; *p*_FDR_=0·014; Psych-PC1 on internalising: β=0·024 [95% CI 0·008 to 0·041]; *p*_FDR_=0·027; Psych-PC1 on total: β=0·028 [95% CI 0·012 to 0·044]; *p*_FDR_=0·007). These overall indicate IGEs of maternal general neuropsychiatric liabilities on internalising difficulties in children.

In contrast, child Psych-Mean and Psych-PC1 scores but not corresponding parental scores showed significant associations with externalising difficulties (Psych-Mean: β=0·021 [95% CI 0·005 to 0·037]; *p*_FDR_=0·048; Psych-PC1: β=0·024 [95% CI 0·007 to 0·042]; *p*_FDR_=0·039), thereby indicating DGEs of general neuropsychiatric liabilities on externalising difficulties.

Likewise, at school age (figures 3–4; table S8), maternal Psych-PC1 were associated with child internalising (β=0·056 [95% CI 0·022 to 0·091]; *p*_FDR_=0·011) and total difficulties (β=0·046 [95% CI 0·016 to 0·077]; *p*_FDR_=0·018), supporting maternal IGEs but with a weaker effect by the father-reporters than the mother-reporters in most studies, except the NTR (figure 4). Child Psych-Mean and Psych-PC1 were instead associated with externalising difficulties (Psych-Mean: β=0·063 [95% CI 0·027 to 0·010]; *p*_FDR_=0·002; Psych-PC1: β=0·079 [95% CI 0·045 to 0·113]; *p*_FDR_<0·0001). Child Psych-PC1 also predicted total difficulties (β=0·056 [95% CI 0·017 to 0·096]; *p*_FDR_=0·025).

In UVMA, maternal Psych-PC1 remained significantly associated with mother-reported but not father-reported child internalising difficulties in both ages (tables S19–S22).

#### Genetic liability to emotional conditions

At preschool age (figure 2; table S7), PGSm-MDD was associated with child’s internalising difficulties (β=0·022 [95% CI 0·007 to 0·038]; *p*_FDR_=0·027) in MVMA, which was also found in maternal ratings with similar effect size in UVMA, although the paternal rating did not reach significance (tables S19–S20).

At school age (figures 3–4; table S8), PGSm-MDD remained associated with internalising (β=0·049 [95% CI 0·017 to 0·081]; *p*_FDR_=0·016) and total difficulties (β=0·052 [95% CI 0·022 to 0·081]; *p*_FDR_=0·012), showing mixed effect directions between raters among studied cohorts (figure 4). PGSc-MDD instead showed associations with externalising (β=0·060 [95% CI 0·030 to 0·090]; *p*_FDR_=0·00040) and total difficulties (β=0·053 [95% CI 0·020 to 0·087]; *p*_FDR_=0·010). In UVMA, PGSm-MDD showed significant IGEs on mother-reported but not father-reported internalising and total difficulties (tables S21–S22).

Similarly, PGSm-PPD was also associated with school-aged internalising (β=0·053 [95% CI 0·023 to 0·083]; *p*_FDR_=0·011) and total difficulties (β=0·048 [95% CI 0·018 to 0·077]; *p*_FDR_=0·014), and showed consistent effect directions between raters (figure S23), whereas PGSc-PPD was associated with externalising difficulties (β=0·041 [95% CI 0·011 to 0·071]; *p*_FDR_=0·022) (figure 3; table S8).

#### Genetic liability to neurodevelopmental conditions

In preschool-aged children (figure 2; table S7), PGSc-ADHD showed associations with externalising (β=0·070 [95% CI 0·038 to 0·10]; *p*_FDR_=0·00011) and total difficulties (β=0·030 [95% CI 0·012, 0·048]; *p*_FDR_=0·008), whereas no effects were observed for either maternal or paternal PGS. Similarly, at school age (figures 3–4; table S8), PGSc-ADHD but not corresponding parental PGSs was associated with externalising (β=0·121 [95% CI 0·091 to 0·151]; *p*_FDR_<0·0001) and total difficulties (β=0·072 [95% CI 0·037 to 0·107]; *p*_FDR_=0·00052), showing a most consistent effect between raters across studied cohorts (figure 4). The significance of PGSc-ADHD association with externalising difficulties was confirmed in both maternal and paternal ratings at preschool age and in maternal rating at school age (tables S19–22), thereby clearly demonstrating DGEs of ADHD liability.

In preschool-aged children (figure 2; table S7), PGSm-ASD was associated with higher externalising (β=0·031 [95% CI 0·015 to 0·047]; *p*_FDR_=0·002) and total difficulties (β=0·029 [95% CI 0·013 to 0·044]; *p*_FDR_=0·007), but not internalising difficulties, whereas no association was observed for PGSc-ASD. Notably, these maternal associations were only observed for mother-reported but not father-reported outcomes in UVMA (tables S19–20). Furthermore, no behavioural associations were observed for any autism PGSs at school age (figure 3; table S8).

#### Genetic liability to educational attainment

PGSc-EA was inversely associated with externalising and total difficulties at both preschool (externalising: β=−0·059 [95% CI −0·080 to −0·038]; *p*_FDR_<0·0001; total: β=−0·047 [95% CI −0·063 to −0·031]; *p*_FDR_<0·0001; figure 2) and school ages (externalising: β=−0·095 [95% CI −0·127 to −0·063]; *p*_FDR_<0·0001; total: β=−0·095 [95% CI −0·125 to −0·064]; *p*_FDR_<0·0001; figures 3–4), showing nevertheless a consistently weaker effect by fathers than mothers across studied cohorts (figure 4). The negative associations of PGSc-EA with externalising difficulties at preschool age and with total difficulties at school age were confirmed by both mother-reported and father-reported data in UVMA (tables S19–22). No significant associations were observed for parental PGSs in either age group, suggesting little evidence of IGEs driven by EA on child behavioural difficulties (figures 2–4; tables S7–S8).

#### Genetic liability to other mental health conditions and related traits

At both preschool and school ages, parental and offspring PGSs for ALC, AN, ANX, BD, CIG, INSOM, NEUROT, OCD, PTSD, and SCZ did not show any significant associations with child behaviour difficulties (figures 2–3).

### Sensitivity Analysis

Since genotyping procedures differed in fathers, compared to the mothers and the child, in the ALSPAC, we conducted a sensitivity analysis excluding this cohort to ensure the robustness of our findings for school-aged children. The direction of DGEs and IGEs were consistent, while effect estimates were slightly attenuated and less precise (e.g., for internalising difficulties, PGSm-MDD: β=0·045 [95% CI 0·010 to 0·079; PGSm-PPD: β=0·042 [95% CI 0·010 to 0·074; maternal Psych-PC1: β=0·050 [95% CI 0·013 to 0·087]; all *p*_FDR_=0·052; tables S27–S28).

## Discussion

In this multi-cohort study, we investigated the influences of parental polygenic liabilities to mental health conditions and related traits on child behavioural difficulties via direct and indirect pathways. We leveraged data from a wider range of samples, raters, time points, predictors, and outcomes than previous studies. We mainly observed evidence of DGEs on child externalising difficulties. In contrast, IGEs were found for child internalising and total difficulties in both age groups. The most robust evidence of IGEs was for maternal depression and general neuropsychiatric liabilities. This has not been previously demonstrated at similar scale but corroborates with recent within-sibship and within-family GWAS findings, showing notable attenuation of population genetic effects for depression.^23,24^ Our findings provide an insight into aetiology of externalising and internalising difficulties, thereby informing future prevention of childhood behavioural difficulties.

The consistency of the observed DGEs on externalising difficulties and IGEs on internalising difficulties is striking. Many previous parent-offspring genomic studies have focused on externalising difficulties.^20,25,26^ Corroboratively, another recent study on an independent single cohort than ours compared both dimensions and highlighted the importance of IGEs on adolescent internalising but not externalising behaviours.^21^ Previous twin-based studies also demonstrate a higher heritability of externalising traits than internalising traits.^4^ These findings call for a future focus on identifying mechanisms of IGEs on internalising traits, including potential environmental mediation, a current gap in the literature. Notably, parental IGEs do not exclusively consist of genetic nurture but also other mechanisms such as assortative mating and population stratification.^7^

Assortative mating is however less likely to explain the different patterns of results for externalising and internalising difficulties, as otherwise we would have expected to see higher spousal correlations for internalising traits (e.g., depression, anxiety) than externalising traits (e.g., ADHD, alcohol use), which we did not. If substantiated in future studies accounting for alternative mechanisms explicitly, we may better understand differences in the aetiology of internalising and externalising difficulties.

We observed the most robust evidence of IGEs for maternal liabilities. This may be explained by biological mechanisms such as DNA methylations,^27,28^ or alternatively that mothers spent more time with their children. A key issue with most previous studies in the field is that maternal effects may have been inflated by solely relying on mother-reported offspring behaviours, as a mother’s own liability to for example depression might bias reporting of her child’s outcomes. Here, we sought to account for this possibility by including data reported by both mothers and fathers. This helped us discern nurturing from reporting effects underlying IGEs of parental liabilities. We observed small but robust IGEs of maternal depression (MDD and PPD) and general neuropsychiatric liabilities on child internalising and total difficulties across maternal and paternal ratings, although the number of studies including paternal reports was small. In contrast, the observed IGE of maternal liability to autism on externalising and total difficulties appeared to be mainly present in mother-reported observations at preschool age, indicating possible rater bias influenced by maternal autism liability. However, note that differences between raters may also be compatible with random sampling variance, as statistical precision was limited due to the lower sample size of our paternal ratings, highlighting the necessity of collecting data from multiple raters to substantiate these initial findings.

We observed that child but not parental liability to ADHD was consistently and robustly associated with externalising and total behavioural difficulties across raters, age groups and model types. This corroborates previous studies^9,11,15^ and supports the notion of direct genetic inheritance of ADHD, with IGEs playing a minimal role. In addition, genetic liability to EA played a direct but protective role in child behaviours, conflicting with some earlier studies showing indirect genetic influences of EA.^9,16,18^ Note that we included maternal education as a covariate in our models, allowing us to control for wider confounding associated with this trait on child mental health outcomes, but likely deflating parental IGEs for EA and possibly other traits.

Although we lacked sufficient samples to test this directly, genetic effects appeared generally larger at school age. This may be driven by increased expression of behavioural and emotional difficulties^29^ or decreased measurement error of outcomes as children grow older. Alternatively, reliance on adult-based GWASs to derive PGSs may play a role. GWAS based on child and adolescent samples, ideally incorporating longitudinal data to disentangle stability and change in DGEs and IGEs over time, should be a key priority for this field to progress forward.

This study has some limitations which may restrict the generalisability of our findings. First, our analyses were based solely on European ancestry samples, which may not transfer well to other ancestries^30^. Second, although study-level heterogeneity was generally low, we cannot completely rule out biases caused by differences in genotyping and QC procedure, as well as phenotype measurement among cohorts. The different approach of genotyping fathers in the ALSPAC was a particularly important limitation, although point estimates only attenuated slightly in the sensitivity analysis excluding this cohort. Third, statistical power was a constraint in some analyses, especially given the low number of studies including paternal ratings, which prevented us from conducting meta-regression analyses to compare effect sizes between the raters, child ages, and behavioural dimensions. Fourth, both maternal and paternal ratings may inflate estimates of IGEs, hence future studies should also include other measures such as child self-reports, teacher ratings, or clinical diagnoses.

In summary, our findings support direct genetic influences of ADHD and EA and indirect genetic influences of maternal depression and general neuropsychiatric liabilities on offspring difficulties. The observed indirect maternal effects on child internalising difficulties across samples, raters, and time points call for further substantiating and mechanistic investigations.

## Data Availability

Correspondence and requests for materials should be addressed to

https://github.com/HappyMumWP4/Direct-and-Indirect-Genetic-Effects-/tree/master

## Author contributions

L.T., A.D.A., and M.S. drafted this article. All authors reviewed or edited this article. J.L., L.T., M.S., K.R., and A.N. conceptualized this study. L.T., M.S., A.D.A., J.L., A.N., K.R., A.K.S.H., L.F.U., B.V., and F.B. contributed to the methodology used in this study. A.D.A., E.V., B.S.P., R.P., L.D.V., M.B., S.V., J.F., E.Z., L.T., M.L.P., J.T., T.R., A.S., C.P., and D.C. contributed to data curation in this study. L.T., M.S. A.D.A., E.V., R.P., L.D.V., S.V., J.F., J.T., T.R., and A.S. contributed to formal analyses in this study. L.T. and J.L. coordinated and administered this study. J.L., A.N., A.K.S.H., C.A.M.C., B.S.P., M.B., S.H.W., M.G., K.R., C.K., D.C., and A.J.F. supervised this study. A.K.S.H., H.A., O.A.A., D.I.B., J.F., M.G., J.L., K.R., T.R., A.S., and C.P. contributed to individual cohort investigation. A.N., C.A.M.C., O.A.A., A.K.S.H., B.S.P., M.B., D.I.B., S.H.W., J.L., K.R., M.L.P., C.K., A.J.F., B.V., and F.B. contributed to funding for this work. All authors gave the final approval of the current version of this article.

## Acknowledgments

We thank Mental Health Norway represented by Jon Fabritius, Andrea Kjennerud, Camilla Gundersen, and the other participants in the lived experience feedback meetings, for their valuable contributions. In addition, we thank Martin Scheiene for helping facilitate the meetings together with A.D.A. We also thank the EAGLE consortium partners. A.D.A., A.K.S.H., and C.A.M.C. were supported by the European Union’s Horizon Europe Research and Innovation Programme (FAMILY, #101057529). C.A.M.C., J.L., K.R., B.V., and F.B were supported by the European Union (HappyMums, #101057390). J.L. was also supported by the Strategic Research Council (SRC) established within the Academy of Finland (#352700 and #372317). C.A.M.C. was also supported by the European Research Council (ERC; TEMPO, #101039672). A.K.S.H. was also supported by the Research Council of Norway (RCN; #336085) and the South-Eastern Norway Regional Health Authority (#2020022; #2026069). B.S.P. was funded by the Horizon Europe, R2D2-MH, #101057385; the UK Research and Innovation (UKRI; under the UK government’s Horizon Europe funding guarantee, R2D2-MH, #10039383); and the Swiss State Secretariat for Education, Research and Innovation (SERI; R2D2-MH, #22.00277). D.I.B. received a KNAW Academy Professor Award (PAH/6635). J.L., K.R. and M.L.P. were funded by the Research Council of Finland (RCF; J.L.: #269925, #311617 and #352700; K.R.: #128789, #1287891, and #1312670; M.L.P.: #330206, #330209, and #358095). C.K. and A.J.F. were supported by the German Research Foundation (DFG; #458609264); C.K. was also supported by the DFG (#220286500).

Views and opinions expressed here are, however, those of the author(s) only and do not necessarily reflect those of the European Union and granting authorities. Neither the European Union nor the granting authorities can be held responsible for them.

## Supplementary Materials

Supplementary appendix

## Additional information

The codes used for all analyses are available on Github: GitHub - HappyMumWP4/Direct-and-Indirect-Genetic-Effects- at master.

