## Supplementary document for "Direct and Indirect Genetic Effects of Parental Liabilities to Mental Health Conditions and Related Traits on Children’s Behavioural Difficulties: A Multi-Cohort Study"

#### Table of Contents


Rapid review of the literature p2

Systematic literature search p7

Cohort descriptions and acknowledgements p17

Genotyping and data processing in each cohort p27

Polygenic score (PGS) calculation in each cohort p34

Child behavioral phenotyping in each cohort p37

Methods used in cohort-level statistical analyses p41

Methods used in meta-analyses p41

Evidence of parental genetic effects in univariate meta-analysis (UVMA) p42

Evidence of parental genetic effects on clinically relevant child behavioral difficulties p44

Sensitivity analysis of multivariate meta-analysis (MVMA) p45

Supplementary figures and legends p46

References p73

### Rapid review of the literature

To assess the current state of the evidence, we conducted a rapid review of indirect genetic effects (IGEs) on child and adolescent mental health. To this end, we adapted the search strategy from Wang et al.[1] to identify studies that had employed trio polygenic score (PGS) designs to estimate IGEs on mental health outcomes in offspring (see detailed search strategy in the section "Systematic literature search: Which polygenic scores tend to show indirect effects on mental health-related outcomes in early life?"). No restrictions were placed on which traits were used for PGS calculation in parents and offspring. We systematically searched the MEDLINE, Embase, PsycInfo, and Web of Science databases on 6 March 2025. The search was conducted by Senior Librarian Trude Anine Muggerud at the Norwegian Institute of Public Health library, in collaboration with ADA. The search identified 309 primary studies, of which 141 remained after duplicate removal. After screening the titles and abstracts, 23 potentially relevant studies remained. Further in-depth screening of abstracts and full papers resulted in 12 studies eligible for inclusion (see table S1 below). From each study, we extracted information about the sample, methods, and whether the results provided support for IGEs based on a 5% alpha threshold (multiple testing corrected if available).

In table S29, we present the main characteristics of the reviewed studies, organised by their overall support for IGEs (top) or lack of support thereof (bottom). Overall, the most common methodology to detect IGEs was 'Trio PGS'. The most common outcomes in children were externalising and neurodevelopmental traits (10 out of 12 studies). Studies, which showed evidence of IGEs had larger sample sizes overall (approximately >19,000 parent-offspring trios). There were no published multi-cohort studies.

**Table S1.** Summary of rapid review results on indirect genetic effects on child/adolescent mental health

| Authors (Year) | Method | Sample | N trios | Outcome | Sig. IGE | Non-sig. IGE |
|---|---|---|---|---|---|---|
| Pingault et al. (2023)[2] | Trio PGS | Pop-based (EUR) | 19,506 | ADHD traits | Maternal: neuroticism, cognition and EA. | ADHD, schizophrenia, bipolar disorder, |

| | | | | | | |
|---|---|---|---|---|---|---|
| | | | | | Paternal: alcohol use. | depression, anxiety, smoking, cannabis use. |
| Hegemann et al. (2025) [3] | Trio PGS | Pop-based (EUR) | 24,692 | Hyperactivity | Maternal: autism, EA, cognitive ability. | ADHD, dyslexia. |
| | | | | Inattention | Paternal: EA. | Cognitive ability, ADHD, dyslexia. |
| | | | | Motor skills | Maternal and paternal: EA, cognitive ability. | Autism, ADHD and dyslexia. |
| | | | | Language skills | None | Autism, EA, cognitive ability, ADHD and dyslexia. |
| | | | | Social/communication skills | Maternal: EA. | Autism, cognitive ability, ADHD, dyslexia. |
| | | | | Repetitive and restrictive behaviours | Maternal: cognitive ability. | Autism, EA, ADHD, dyslexia. |
| Askelund et al. (2025) [4] | Trio PGS | Pop-based (EUR) | 33,351 | Intercept of CBCL total problems | Maternal: autism, depression. | ADHD, anorexia, Tourette's syndrome, OCD, schizophrenia, bipolar disorder, alcohol dependence, anxiety, PTSD. |

| | | | | | | |
|---|---|---|---|---|---|---|
| | | | | Intercept of differentiation | None | ADHD, anorexia, Tourette's syndrome, OCD, schizophrenia, bipolar disorder, autism, depression, alcohol dependence, anxiety, PTSD. |
| Kuo et al. (2022) [5] | Trio PGS linear | Clinical (parents with/without alcohol dependence, European and African) | 1,111 | Externalising behaviours in adolescence | Externalising (EUR). | Externalising (AFR). |
| van der Laan et al. (2023) [6] | T/NT PGS | Pop-based (EUR twins) | 3,024 | Child aggression | None | Early-life aggression, EA, ADHD. |
| Voronin et al. (2024) [7] | Trio PGS and twin | Pop-based (EUR twins) | 415 | Hyperactivity and inattention | None | ADHD, EA. |
| Axelrud et al. (2023) [8] | Trio PGS | Population (high-risk, Admixed Brazilian) | 1,017 | ADHD traits and psychotic symptoms | None | Cognition, EA, ADHD, schizophrenia. |

| | | | | | | |
|---|---|---|---|---|---|---|
| Frach et al. (2024) [9] | Trio PGS | Pop-based (EUR) | 31,290 | Conduct problems | None | ADHD, depression, anxiety, antisocial behaviour, lifetime smoking, problematic alcohol use, cannabis use disorder, cognitive performance, EA, household income, risky behaviours, age at first birth. |
| de Zeeuw et al. (2020) [10] | T/NT PGS | Pop-based (EUR twins) | 2,649 | ADHD traits | None | EA, ADHD. |
| Nayar et al. (2021) [11] | Trio PGS | Clinical (EUR autistic children) | 2,614 | Autism-related traits | None | Autism. |
| Birmaher et al. (2022) [12] | Logistic/ Cox regression | Clinical (parents and offspring w/wo bipolar disorder, EUR) | 331 | Bipolar disorder | None | Bipolar disorder. |
| Shakeshaft et al. (2024) [13] | T/NT PGS | Pop-based (EUR and SAS) | 3,063 | Emotional disorders | None | Anxiety, major depression, broad depression, bipolar disorder, schizophrenia, ADHD, autism. |

| | | | | | | |
|---|---|---|---|---|---|---|
| | | | | Emotional symptoms | None | Anxiety, major depression, broad depression, bipolar disorder, schizophrenia, ADHD, autism. |

Notes: AFR = African; EA = educational attainment; EUR = European; PGS = polygenic scores; Pop-based = Population-based; T/NT = transmitted / non-transmitted; SEA = Southeast Asian.
*Here we report only conditional associations within the trio design.

The studies included in our review used similar methods but focused on a heterogeneous set of outcomes, ranging from specific psychiatric symptoms and neurodevelopmental traits measured in the general population to paediatric bipolar disorder and autism in clinical cohorts. The three studies relying on the transmitted/non-transmitted PGS design (with cohort sizes ranging between 2,649-3,063) did not identify any significant indirect genetic effects. This may be related to insufficient statistical power due to both limited sample sizes and the method employed. The same consideration likely applies to the two studies with less than 500 trios. The remaining seven studies (of which four were based on the Norwegian Mother, Father, and Child Cohort study; MoBa) were more likely to identify significant indirect genetic effects, with a few exceptions.

Among the studies that showed evidence of indirect genetic effects, parental PGSs for autism showed an indirect effect on different child outcomes, driven by the mothers (two MoBa studies). The parental educational attainment (EA) PGS showed an indirect association with ADHD traits in children in two MoBa studies. This association was sometimes in a positive direction of association (i.e., higher PGS for EA in parents positively associated with more ADHD-related traits). As some authors have noted, however, these findings might be due to shared rater bias originating from using reports from only one parent.[9] Indeed, most previously identified significant IGEs were of maternal genetic liabilities on mother-reported offspring outcomes, raising the possibility that these IGEs are inflated by reporting effects. Finally, parental PGS for cognitive ability were positively associated with a range of neurodevelopmental traits after accounting for the child PGS. In sum, the findings that are consistent across the included studies are mainly from the same sample (MoBa). The current evidence base is thus very limited and requires both studying many unexplored traits and replication of previous findings in independent cohorts and across raters.

#### Systematic literature search: "Which polygenic scores tend to show indirect effects on mental health-related outcomes in early life?"

| | |
|---|---|
| **Institution:** | Library at the Norwegian Institute of Public Health |
| **Contact person:** | Adrian Dahl Askelund |
| **Search:** | Trude Anine Muggerud |
| **Reviewer:** | Bente Foss |
| **Duplicate check in EndNote:** | Before duplicate removal: 309 primary studies, 15 systematic reviews. After duplicate removal: 141 primary studies, 8 systematic reviews |
| **Database:** | Ovid MEDLINE(R) and Epub Ahead of Print, In-Process, In-Data-Review & Other Non-Indexed Citations, Daily and Versions <1946 to March 05, 2025> |
| **Date:** | 06.03.25 |
| **Numbers:** | 82 primary studies, 3 systematic reviews |

| | | |
|---|---|---|
| 1 | Child/ or Child, Preschool/ or Pediatrics/ or Students/ or Minors/ or Adolescent/ or (child* or pediatric* or paediatric* or toddler* or preschooler? or "pre schooler?" or preadolescen* or prepubescen* or preteen? or tween? or tweenager? or boy? or girl? or kid? or juvenil* or underage* or minor? or pubescen* or student* or adolescen* or teen? or teenage* or (young* adj (people or person*)) or youth*).tw,kf. | 4906298 |

| | | |
|---|---|---|
| 2 | Genetic Risk Score/ or (((polygenic or polygenetic or genetic? or genomic or genotype or genotypic or "genome wide" or genomewide) adj scor*) or ((polygenic or polygenetic or genetic? or genomic or genotype or genotypic or "genome wide" or genomewide) adj ("risk scor*" or "risk assessment?")) or "genetic liabilit*").tw,kf. | 12695 |
| 3 | ("nature of nurtur*" or "genetic nurtur*" or "indirect genetic*" or "virtual parent design" or "pseudo control" or "dynastic effect*" or intergeneration* or multigeneration* or ((transmit* or nontransmit*) adj allele*) or "passive gene environment correlation" or (genetic adj (inheritance or confounding*)) or ((genetic or parental or maternal or paternal or familial) adj transmission) or ((parental or maternal or paternal) adj influence?) or "social inheritance" or "social genetic effect*" or ((environmental or cultural or vertical) adj transmission) or "family based stud*" or "familybased stud*" or trio* or triad* or dual* or dyad*).tw,kf. | 465245 |
| 4 | Mental Disorders/ or Anxiety/ or Panic Disorders/ or Phobic Disorders/ or exp "Feeding and Eating Disorders"/ or Depression/ or Dysthymic Disorder/ or Bipolar Disorder/ or Suicide/ or Self-Injurious Behavior/ or Obsessive-Compulsive Disorder/ or Stress Disorders, Post-Traumatic/ or Adjustment Disorders/ or Conduct Disorder/ or exp Schizophrenia/ or Psychotic Disorders/ or Hallucinations/ or Paranoid Disorders/ or exp Substance-Related Disorders/ or Marijuana Abuse/ or exp Autism Spectrum Disorder/ or Attention Deficit Disorders with Hyperactivity/ or Neurodevelopmental Disorders/ or exp Tic Disorders/ or Dissociative Disorders/ or Depersonalization/ or exp Sleep Wake Disorders/ or (((Mental or psychiat* or psychological) adj (disorder? or disease? or illness* or problem?)) or "nervous breakdown?" or ("mental health" adj | 2082674 |

| | | |
|---|---|---|
| | (disorder? or disease? or illness* or problem?)) or ((psychological or emotional) adj (distress or stress or problem?)) or psychopatholog* or internali* or externali* or "conduct disorder?" or "oppositional defiant disorder?" or (eating adj2 disorder*) or anorexi* or bulimi* or "binge eating" or orthorexi* or anxiety or anxious or angst or "panic disorder?" or phobi* or agoraphobi* or ((bipolar or manic) adj (disorder? or illness)) or "mood disorder?" or "bipolar affective psychos#s" or depression or depressive or dysthymi* or GAD or suicid* or (self adj (harm* or injur* or wounding or mutilation)) or ("self inflicted" adj (harm or injur* or wound?)) or "self injurious behavio?r" or automutilation or "obsessive compulsive disorder?" or OCD or "compulsive obsessi* disorder?" or ((compulsive or obsessive) adj neurosis) or (("post traumatic" or posttraumatic) adj stress) or "acute stress disorder?" or "traumatic stress" or PTSD or "adjustment disorder?" or schizophren* or "psychotic disorder?" or psychoses or psychosis or "delusional disorder?" or hallucinat* or "paranoid disorder?" or paranoia? or "schizoaffective disorder?" or (substance adj ("use" or abuse or related) adj disorder?) or (alcohol adj ("use disorder?" or abuse or addiction or addict? or dependence or dependency)) or alcoholism or ((drug or opioid or cannabis or marihuana or marijuana or hashish or stimulant or sedative) adj ("use disorder?" or abuse or addiction or addict? or dependence or dependency)) or "behavio?ral addiction?" or "gambling disorder?" or neurodevelopment* or autis* or asd or "Asperger syndrom*" or adhd or "attention deficit hyperactivity disorder?" or "attention deficit disorder?" or "attention deficit and disruptive behavio?r disorder?" or tic? or Tourette* or (dissociative adj (disorder? or hysteria?)) or dereali#ation or depersonali#ation or (sleep* adj (disorder? or deprivation or deprived or problem? or difficult*)) or dyssomnia? or parasomnia? or insomnia?).tw,kf. | |
| 5 | 1 and 2 and 3 and 4 | 97 |
| 6 | limit 5 to yr="2014 -Current" | 85 |
| 7 | limit 6 to english | 85 |
| 8 | (comment or editorial or letter).pt. | 2315626 |
| 9 | 7 not 8 | 85 |
| 10 | limit 9 to "reviews (maximizes specificity)" | 1 |
| 11 | Meta-Analysis/ or Network Meta-Analysis/ or ((systematic* adj2 review*) or metaanal* or "meta anal*" or (review and ((structured or database* or | 617116 |

| | | |
|---|---|---|
| | systematic*) adj2 search*)) or "integrative review*" or (evidence adj2 review*)).tw,kf,bt. | |
| 12 | 10 or (9 and 11) | 3 |
| 13 | 9 not 12 | 82 |

**Database:** Embase <1974 to 2025 March 05>

**Date:** 06.03.25

**Numbers:** 68 primary studies, 3 systematic reviews

| | | |
|---|---|---|
| 1 | child/ or boy/ or girl/ or preschool child/ or school child/ or childhood/ or juvenile/ or pediatrics/ or toddler/ or student/ or elementary student/ or middle school student/ or minor/ or exp Adolescence/ or exp Adolescent/ or high school student/ or (child* or pediatric* or paediatric* or preadolescen* or prepubescen* or preteen? or tween? or tweenager? or boy? or girl? or kid? or juvenil* or underage* or minor? or pubescen* or student* or toddler* or preschooler? or "pre schooler?" or adolescen* or teen? or teenage* or (young* adj (people or person*)) or youth*).tw,kf. | 5296172 |
| 2 | genetic risk score/ or (((polygenic or polygenetic or genetic? or genomic or genotype or genotypic or "genome wide" or genomewide) adj scor*) or ((polygenic or polygenetic or genetic? or genomic or genotype or genotypic or "genome wide" or genomewide) adj ("risk scor*" or "risk assessment?")) or "genetic liabilit*").tw,kf. | 22295 |
| 3 | "mother to child transmission"/ or "maternal fetal transmission"/ or ("nature of nurtur*" or "genetic nurtur*" or "indirect genetic*" or "virtual parent design" or "pseudo control" or "dynastic effect*" or intergeneration* or multigeneration* or ((transmit* or nontransmit*) adj allele*) or "passive gene environment correlation" or (genetic adj (inheritance or confounding*)) or ((genetic or parental or maternal or paternal or familial) adj transmission) or ((parental or maternal or paternal) adj influence?) or "social inheritance" or "social genetic effect*" or ((environmental or cultural or vertical) adj transmission) or "family based stud*" or "familybased stud*" or trio* or triad* or dual* or dyad*).tw,kf. | 580055 |

| | | |
|---|---|---|
| 4 | mental disease/ or conduct disorder/ or oppositional defiant disorder/ or eating disorder/ or anxiety/ or anxiety disorder/ or panic/ or phobia/ or bipolar disorder/ or mood disorder/ or suicide/ or depression/ or major depression/ or automutilation/ or obsessive compulsive disorder/ or posttraumatic stress disorder/ or adjustment disorder/ or schizophrenia/ or psychosis/ or delusional disorder/ or hallucination/ or paranoia/ or schizoaffective psychosis/ or substance abuse/ or alcohol abuse/ or alcoholism/ or drug abuse/ or drug dependence/ or cannabis addiction/ or behavioral addiction/ or pathological gambling/ or autism/ or attention deficit hyperactivity disorder/ or tic/ or Gilles de la Tourette syndrome/ or dissociative disorder/ or depersonalization/ or sleep disorder/ or (((Mental or psychiat* or psychological) adj (disorder? or disease? or illness* or problem?)) or "nervous breakdown?" or ("mental health" adj (disorder? or disease? or illness* or problem?)) or ((psychological or emotional) adj (distress or stress or problem?)) or psychopatholog* or internali* or externali* or "conduct disorder?" or "oppositional defiant disorder?" or (eating adj2 disorder*) or anorexi* or bulimi* or "binge eating" or orthorexi* or anxiety or anxious or angst or "panic disorder?" or phobi* or agoraphobi* or ((bipolar or manic) adj (disorder? or illness)) or "mood disorder?" or "bipolar affective psychos#s" or depression or depressive or dysthymi* or GAD or suicid* or (self adj (harm* or injur* or wounding or | 2702345 |
| | mutilation)) or ("self inflicted" adj (harm or injur* or wound?)) or "self injurious behavio?r" or automutilation or "obsessive compulsive disorder?" or OCD or "compulsive obsessi* disorder?" or ((compulsive or obsessive) adj neurosis) or (("post traumatic" or posttraumatic) adj stress) or "acute stress disorder?" or "traumatic stress" or PTSD or "adjustment disorder?" or schizophren* or "psychotic disorder?" or psychoses or psychosis or "delusional disorder?" or hallucinat* or "paranoid disorder?" or paranoia? or "schizoaffective disorder?" or (substance adj ("use" or abuse or related) adj disorder?) or (alcohol adj ("use disorder?" or abuse or addiction or addict? or dependence or dependency)) or alcoholism or ((drug or opioid or cannabis or marihuana or marijuana or hashish or stimulant or sedative) adj ("use disorder?" or abuse or addiction or addict? or dependence or dependency)) or "behavio?ral addiction?" or "gambling disorder?" or neurodevelopment* or autis* or asd or "Asperger syndrom*" or adhd or "attention deficit hyperactivity disorder?" or "attention deficit disorder?" or "attention deficit and disruptive behavio?r disorder?" or tic? or Tourette* or (dissociative adj (disorder? or hysteria?)) or dereali#ation or depersonali#ation or (sleep* adj (disorder? or deprivation or deprived or problem? or difficult*)) or dyssomnia? or parasomnia? or insomnia?).tw,kf. | |
| 5 | 1 and 2 and 3 and 4 | 211 |
| 6 | limit 5 to embase status | 83 |
| 7 | limit 6 to yr="2014 -Current" | 72 |

| | | |
|---|---|---|
| 8 | limit 7 to english | 72 |
| 9 | (Conference Abstract or Letter or Editorial).pt. | 7569002 |
| 10 | 8 not 9 | 71 |
| 11 | limit 10 to "reviews (maximizes specificity)" | 1 |
| 12 | exp Meta-Analysis/ or "systematic review"/ or ((systematic* adj2 review*) or metaanal* or "meta anal*" or (review and ((structured or database* or systematic*) adj2 search*)) or "integrative review*" or (evidence adj2 review*)).tw,kf,bt. | 875857 |
| 13 | 11 or (10 and 12) | 3 |
| 14 | 10 not 13 | 68 |

**Database:** APA PsycInfo <1806 to March 2025 Week 1>

**Date:** 06.03.25

**Numbers:** 62 primary studies, 5 systematic reviews

| | | |
|---|---|---|
| 1 | Students/ or exp Elementary School Students/ or Middle School Students/ or Junior High School Students/ or Kindergarten Students/ or exp Preschool Students/ or Pediatrics/ or College Students/ or Middle School Students/ or Graduate Students/ or High School Students/ or Junior High School Students/ or Reentry Students/ or Special Education Students/ or Transfer Students/ or High School graduates/ or ("100" or "140" or "160" or "180" or "200").ag. or (child* or pediatric* or paediatric* or toddler* or preschooler? or "pre schooler?" or preadolescen* or prepubescen* or preteen? or tween? or tweenager? or boy? or girl? or kid? or juvenil* or underage* or minor? or pubescen* or student* or adolescen* or teen? or teenage* or (young* adj (people or person*)) or youth*).tw. | 1900035 |
| 2 | (((polygenic or polygenetic or genetic? or genomic or genotype or genotypic or "genome wide" or genomewide) adj scor*) or ((polygenic or polygenetic or genetic? or genomic or genotype or genotypic or "genome wide" or genomewide) adj ("risk scor*" or "risk assessment?")) or "genetic liabilit*").tw. | 3175 |

| | | |
|---|---|---|
| 3 | Nature Nurture/ or ("nature of nurtur*" or "genetic nurtur*" or "indirect genetic*" or "virtual parent design" or "pseudo control" or "dynastic effect*" or intergeneration* or multigeneration* or ((transmit* or nontransmit*) adj allele*) or "passive gene environment correlation" or (genetic adj (inheritance or confounding*)) or ((genetic or parental or maternal or paternal or familial) adj transmission) or ((parental or maternal or paternal) adj influence?) or "social inheritance" or "social genetic effect*" or ((environmental or cultural or vertical) adj transmission) or "family based stud*" or "familybased stud*" or trio* or triad* or dual* or dyad*).tw. | 134891 |
| 4 | Mental Disorders/ or Psychological Stress/ or Psychopathology/ or Internalization/ or Externalization/ or Conduct Disorder/ or Oppositional Defiant Disorder/ or Eating Disorder/ or Anxiety/ or Panic Disorder/ or Phobias/ or exp Bipolar Disorder/ or Affective Disorders/ or Major Depression/ or "Depression (Emotion)"/ or Suicide/ or Nonsuicidal Self-Injury/ or Self-Inflicted Wounds/ or Obsessive Compulsive Disorder/ or Posttraumatic Stress Disorder/ or Adjustment Disorder/ or Schizophrenia/ or Psychosis/ or Delusional Disorder/ or Hallucinations/ or Paranoid Psychosis/ or Paranoia/ or Schizoaffective Disorder/ or Drug Abuse/ or Drug Dependency/ or Alcohol Abuse/ or "Opioid Use Disorder"/ or Nonsubstance Related Addiction/ or Gambling Disorder/ or Neurodevelopmental Disorders/ or Autism Spectrum Disorders/ or Attention Deficit Disorder with Hyperactivity/ or Tics/ or Tourette Syndrome/ or Dissociative Disorders/ or "Depersonalization/Derealization Disorder"/ or Depersonalization/ or Sleep Wake Disorder/ or (((Mental or psychiat* or psychological) adj (disorder? or disease? or illness* or problem?)) or "nervous breakdown?" or ("mental health" adj (disorder? or disease? or illness* or problem?)) or ((psychological or emotional) adj (distress or stress or problem?)) or psychopatholog* or internali* or externali* or "conduct disorder?" or "oppositional defiant disorder?" or (eating adj2 disorder*) or | 1304457 |

1

| | | |
|---|---|---|
| 4 | Mental Disorders/ or Psychological Stress/ or Psychopathology/ or Internalization/ or Externalization/ or Conduct Disorder/ or Oppositional Defiant Disorder/ or Eating Disorder/ or Anxiety/ or Panic Disorder/ or Phobias/ or exp Bipolar Disorder/ or Affective Disorders/ or Major Depression/ or "Depression (Emotion)"/ or Suicide/ or Nonsuicidal Self-Injury/ or Self-Inflicted Wounds/ or Obsessive Compulsive Disorder/ or Posttraumatic Stress Disorder/ or Adjustment Disorder/ or Schizophrenia/ or Psychosis/ or Delusional Disorder/ or Hallucinations/ or Paranoid Psychosis/ or Paranoia/ or Schizoaffective Disorder/ or Drug Abuse/ or Drug Dependency/ or Alcohol Abuse/ or "Opioid Use Disorder"/ or Nonsubstance Related Addiction/ or Gambling Disorder/ or Neurodevelopmental Disorders/ or Autism Spectrum Disorders/ or Attention Deficit Disorder with Hyperactivity/ or Tics/ or Tourette Syndrome/ or Dissociative Disorders/ or "Depersonalization/Derealization Disorder"/ or Depersonalization/ or Sleep Wake Disorder/ or (((Mental or psychiat* or psychological) adj (disorder? or disease? or illness* or problem?)) or "nervous breakdown?" or ("mental health" adj (disorder? or disease? or illness* or problem?)) or ((psychological or emotional) adj (distress or stress or problem?)) or psychopatholog* or internali* or externali* or "conduct disorder?" or "oppositional defiant disorder?" or (eating adj2 disorder*) or | 1304457 |
| | anorexi* or bulimi* or "binge eating" or orthorexi* or anxiety or anxious or angst or "panic disorder?" or phobi* or agoraphobi* or ((bipolar or manic) adj (disorder? or illness)) or "mood disorder?" or "bipolar affective psychos#s" or depression or depressive or dysthymi* or GAD or suicid* or (self adj (harm* or injur* or wounding or mutilation)) or ("self inflicted" adj (harm or injur* or wound?)) or "self injurious behavio?r" or automutilation or "obsessive compulsive disorder?" or OCD or "compulsive obsessi* disorder?" or ((compulsive or obsessive) adj neurosis) or (("post traumatic" or posttraumatic) adj stress) or "acute stress disorder?" or "traumatic stress" or PTSD or "adjustment disorder?" or schizophren* or "psychotic disorder?" or psychoses or psychosis or "delusional disorder?" or hallucinat* or "paranoid disorder?" or paranoia? or "schizoaffective disorder?" or (substance adj ("use" or abuse or related) adj disorder?) or (alcohol adj ("use disorder?" or abuse or addiction or addict? or dependence or dependency)) or alcoholism or ((drug or opioid or cannabis or marihuana or marijuana or hashish or stimulant or sedative) adj ("use disorder?" or abuse or addiction or addict? or dependence or dependency)) or "behavio?ral addiction?" or "gambling disorder?" or neurodevelopment* or autis* or asd or "Asperger syndrom*" or adhd or "attention deficit hyperactivity disorder?" or "attention deficit disorder?" or "attention deficit and disruptive behavio?r disorder?" or tic? or Tourette* or (dissociative adj (disorder? or hysteria?)) or dereali#ation or depersonali#ation or (sleep* adj (disorder? or deprivation or deprived or problem? or difficult*)) or dyssomnia? or parasomnia? or insomnia?).tw. | |

| | | |
|---|---|---|
| 5 | 1 and 2 and 3 and 4 | 92 |
| 6 | limit 5 to yr="2014 -Current" | 77 |
| 7 | limit 6 to english | 69 |
| 8 | ("comment/reply" or editorial or letter).dt. | 216409 |
| 9 | 7 not 8 | 67 |
| 10 | limit 9 to "reviews (maximizes specificity)" | 4 |
| 11 | (meta analysis or "systematic review").md. or meta analysis/ or ((systematic* adj2 review*) or metaanal* or "meta anal*" or (review and ((structured or database* or systematic*) adj2 search*)) or "integrative review*" or (evidence adj2 review*)).tw. | 130264 |
| 12 | 10 or (9 and 11) | 5 |
| 13 | 9 not 12 | 62 |

**Database: Web of Science Core Collection: Science Citation Index Expanded (SCI-EXPANDED)--1987-present, Social Sciences Citation Index (SSCI)--1987-present, Arts & Humanities Citation Index. (AHCI)--1987-present, Emerging Sources Citation Index (ESCI)--2019present**

**Date:** 06.03.25

**Note:** Searched with "Exact search"

**Numbers:** 97 primary studies, 4 systematic reviews

| | | |
|---|---|---|
| 1 | TS=(child* or pediatric* or paediatric* or toddler* or preschooler$ or "pre schooler$" or preadolescen* or prepubescen* or preteen$ or tween$ or tweenager$ or boy$ or girl$ or kid$ or juvenil* or underage* or minor$ or pubescen* or student* or adolescen* or teen$ or teenage* or (young* NEAR/0 (people or person*)) or youth*) | 4189885 |

| | | |
|---|---|---|
| 2 | TS=(((polygenic or polygenetic or genetic$ or genomic or genotype or genotypic or "genome wide" or genomewide) NEAR/0 scor*) or ((polygenic or polygenetic or genetic$ or genomic or genotype or genotypic or "genome wide" or genomewide) NEAR/0 ("risk scor*" or "risk assessment$")) or "genetic liabilit*") | 14462 |
| 3 | TS=("nature of nurtur*" or "genetic nurtur*" or "indirect genetic*" or "virtual parent design" or "pseudo control" or "dynastic effect*" or intergeneration* or multigeneration* or ((transmit* or nontransmit*) NEAR/0 allele*) or "passive gene environment correlation" or (genetic NEAR/0 (inheritance or confounding*)) or ((genetic or parental or maternal or paternal or familial) NEAR/0 transmission) or ((parental or maternal or paternal) NEAR/0 influence$) or "social inheritance" or "social genetic effect*" or ((environmental or cultural or vertical) NEAR/0 transmission) or "family based stud*" or "familybased stud*" or trio* or triad* or dual* or dyad*) | 956415 |
| 4 | TS=(((Mental or psychiat* or psychological) NEAR/0 (disorder$ or disease$ or illness* or problem$)) or "nervous breakdown$" or ("mental health" NEAR/0 (disorder$ or disease$ or illness* or problem$)) or ((psychological or emotional) NEAR/0 (distress or stress or problem$)) or psychopatholog* or internali* or externali* or "conduct disorder$" or "oppositional defiant disorder$" or (eating NEAR/1 disorder*) or anorex* or bulimi* or "binge eating" or orthorexi* or anxiety or anxious or angst or "panic disorder$" or phobi* or agoraphobi* or ((bipolar or manic) NEAR/0 (disorder$ or illness)) or "mood disorder$" or "bipolar affective psychosis" or "bipolar affective psychoses" or depression or depressive or dysthymi* or GAD or suicid* or (self NEAR/0 (harm* or injur* or wounding or mutilation)) or ("self inflicted" NEAR/0 (harm or injur* or wound$)) or "self injurious behavior" or "self injurious behaviour" or automutilation or "obsessive compulsive disorder$" or OCD or "compulsive obsessive disorder$" or "compulsive obsessional disorder$" or ((compulsive or obsessive) NEAR/0 neurosis) or (("post traumatic" or posttraumatic) NEAR/0 stress) or "acute stress disorder$" or "traumatic stress" or PTSD or "adjustment disorder$" or schizophren* or "psychotic disorder$" or psychoses or psychosis or "delusional disorder$" or hallucinat* or "paranoid disorder$" or paranoia$ or "schizoaffective disorder$" or (substance NEAR/0 ("use" or abuse or related) NEAR/0 disorder$) or (alcohol NEAR/0 ("use disorder$" or abuse or addiction or | 2107482 |

| | | |
|---|---|---|
| | addict$ or dependence or dependency)) or alcoholism or ((drug or opioid or cannabis or marihuana or marijuana or hashish or stimulant or sedative) NEAR/0 ("use disorder$" or abuse or addiction or addict$ or dependence or dependency)) or "behavioral addiction$" or "behavioural addiction$" or "gambling disorder$" or neurodevelopment* or autis* or asd or "Asperger syndrom*" or adhd or "attention deficit hyperactivity disorder$" or "attention deficit disorder$" or "attention deficit and disruptive behavior disorder$" or "attention deficit and disruptive behaviour disorder$" or tic$ or Tourette* or (dissociative NEAR/0 (disorder$ or hysteria$)) or derealisation or derealization or depersonalisation or depersonalization or (sleep* NEAR/0 (disorder$ or deprivation or deprived or problem$ or difficult*)) or dyssomnia$ or parasomnia$ or insomnia$) | |
| 5 | #4 AND #3 AND #2 AND #1 | 112 |
| 6 | #4 AND #3 AND #2 AND #1 Timespan: 2014-01-01 to 2025-03-06 | 101 |
| 7 | (#4 AND #3 AND #2 AND #1) AND (LA==("ENGLISH")) Timespan: 2014-01-01 to 2025-03-06 | 101 |
| 8 | TS=(("systematic*" NEAR/1 "review*") or ("review" and (("structured" or "database*" or "systematic*") NEAR/1 "search*")) or "integrative review*" or ("evidence" NEAR/1 "review*")) OR TI=("metaanal*" or "meta anal*") OR AB=("metaanal*" or "meta anal*") | 704485 |
| 9 | #7 AND #8 | 4 |
| 10 | #7 not #9 | 97 |

#### Cohort descriptions and acknowledgements

The data for analysing this study was drawn from seven independent prospective early life studies: 1) The Avon Longitudinal Study of Parents and Children (**ALSPAC**): A UK-based birth cohort in Avon, following over 14,000 pregnant women and their children to examine genetic and environmental health influences.[14-18] 2) Generation R (**GenR**) study: A birth cohort in Rotterdam, Netherlands, following participants from foetal life to young adulthood to explore early environmental and genetic influences on development.[19,20] 3) The Millennium Cohort Study (**MCS**): A longitudinal population-based cohort study recruiting children born between 2000 and

2002 across England, Scotland, Wales and Northern Ireland.[21-24] 4) The Norwegian Mother, Father, and Child Cohort Study (**MoBa**), including pregnancies in Norway from 1999-2008.[25-27] 5) The Netherlands Twin Register (**NTR**): A long-term Dutch registry following over 120,000 twins and their families since the early 1980s.[28-30] 6) The Pre-, Peri-, and Postnatal Stress: [Epi-]Genetic Impact on Depression (**POSEIDON**) study: A German longitudinal cohort study recruiting children born between 2010 and 2013.[31,32] 7) Prediction and prevention of preeclampsia and intrauterine growth restriction (**PREDO**) study: A Finnish multi-centre study (2005–2009) focused on pregnant women and their children to identify risk factors for preeclampsia.[33] 8) The German Twin Family Panel (**TwinLife**): A longitudinal study of same-sex twin children (both monozygotic and dizygotic) and their families in Germany, running from 2014 since present, focused on the origins of the development of social inequality.[34-37]

**The Avon Longitudinal Study of Parents and Children (ALSPAC)**

**Cohort profile**: ALSPAC (Avon Longitudinal Study of Parents and Children) is a prospective, population-based study that enrolled pregnant women with expected delivery dates between April 1991 and December 1992. During the initial recruitment phase, a total of 14,541 pregnancies were enrolled. The study has continued as a three-generational study: G0 refers to the original pregnant women, their partners, or other carers, G1 refers to their children (the index children) and G2 refers to the children of the G1 cohort (i.e., the grandchildren). The G1 children were followed until the age of 18, with opportunities to participate in extensive measurements and questionnaires.[14-18] Ethical approval for the study was obtained from the ALSPAC Ethics and Law Committee and the Local Research Ethics Committees. Informed consent for the use of all data collected was obtained from participants in line with the recommendations of the ALSPAC Ethics and Law Committee at the time (NHS Haydock REC: 10/H1010/70). Participants can contact the study team at any time to retrospectively withdraw consent for their data to be used. Study participation was voluntary, and during all data collection sweeps, participants were provided with information on the intended use of their data.

**Funding**:

The UK Medical Research Council and Wellcome (Grant ref: MR/Z505924/1) and the University of Bristol provide core support for ALSPAC. This publication is the work of the authors and AN and MS will be guarantors for the contents of this paper. A comprehensive list of grants funding is available on the ALSPAC website (**http://www.bristol.ac.uk/alspac/external/documents/grant-acknowledgements.pdf**).
Genome wide genotyping data was generated by Sample Logistics and Genotyping Facilities at Wellcome Sanger Institute and LabCorp (Laboratory Corporation of America) using support from 23andMe.

This work was supported by the Erasmus MC Sophia Foundation ("Stiching Vrienden van het Sophia," Grant WAR24-30, AN, MS), the European Research Council (TEMPO, no.101039672, CAMC, AN; iRISK: No 863981; AN, JBP) and the European Union's Horizon Europe Research and Innovation Programme (FAMILY: no.101057529, CAMC, AN; HappyMums: no.101057390, CAMC) for both ALSPAC and GenR.

**Role of funders:**

The funders had no role in the design of the study; in the collection, analysis, and interpretation of the data; in the writing of this manuscript; or in the decision to submit the article for publication.

**Acknowledgements:**

We are extremely grateful to all the families who took part in this study, the midwives for their help in recruiting them, and the whole ALSPAC team, which includes data collection staff, data and administration staff, technical managers, and the technical staff within the Bristol Bioresource Laboratory at the University of Bristol.

**The Generation R (GenR) Study**

Cohort profile: The GenR study is an ongoing population-based prospective cohort study from early foetal life onwards. A total of 9778 pregnant women with 9901 related children were recruited from the municipality of Rotterdam, Netherlands, with an expected delivery date between April 2002 and January 2006.[19,38] These mothers, their children and partners took part in several research waves with an extensive measurements that will be followed until young adulthood The GenR study was approved by the Medical Ethical Committee of Erasmus MC, University Medical Center Rotterdam and is conducted in compliance with the Helsinki Declaration of the World Medical Association, and written informed consent was provided by mothers and parents on behalf of their child.

**Funding**: The GenR Study is made possible by financial support from the Erasmus Medical Center, Rotterdam, the Erasmus University Rotterdam, the Netherlands Organization for Health Research and Development and the Ministry of Health, Welfare and Sport, the European Union's Horizon 2020 research and innovation programme under grant agreements no.633595 (DynaHEALTH) and no.733206 (LifeCycle), and the European Joint Programming Initiative "A Healthy Diet for a Healthy Life" (JPI HHL, NutriPROGRAM project, ZonMw the Netherlands no.529051022). This work was also supported by the Erasmus MC Sophia Foundation ("Stiching Vrienden van het Sophia," Grant WAR24-30, AN, MS).

**Role of funders:** The funders had no role in the planning or execution of the study nor the interpretation or publication of its results.

**Acknowledgements:** The GenR Study is conducted by the Erasmus Medical Center in close collaboration with the Faculty of Social Sciences of the Erasmus University Rotterdam, the Municipal Health Service Rotterdam area, the Rotterdam Homecare Foundation, and the Stichting Trombosedienst & Artsenlaboratorium Rijnmond (STAR-MDC), Rotterdam. We gratefully acknowledge the contribution of children and parents, general practitioners, hospitals, midwives and pharmacies in Rotterdam. The generation and management of GWAS genotype data for the Generation R Study was done at the Human Genomics Facility, HuGe-F, housed within the Laboratory for Population Genomics of the Department of Internal Medicine at Erasmus MC. Genetic Laboratory of the Department of Internal Medicine, Erasmus MC, The Netherlands. We thank Zahra Alawi, Marijn Verkerk, Dr. Katerina Trajanoska, Costanza Vallerga, Samuel Gathan, Dr. Carolina Medina-Gomez, Dr. Linda Broer and Jard de Vries for their help in creating, managing and quality control (QC) the GWAS database.

**The Millennium Cohort Study (MCS)**

**Cohort profile:** The Millennium Cohort Study (MCS)[21-24] is a nationally representative longitudinal population-based cohort study that was set up to follow the lives of children born between September 2000 and January 2002 across England, Scotland, Wales and Northern Ireland. Eligible children were identified using government child benefit records and the sample was constructed to be representative of the total United Kingdom (UK) population, containing 18,552 families (18,827 children) at baseline.[39] The MCS sample members were first surveyed when they were around 9 months of age, and cohort members continue to remain eligible to be surveyed if they remain living in, or return to, the UK. Additional rounds of data collection were performed at ages 3, 5, 7, 11, 14, 17 and 23 years. Saliva samples were collected at an approximate child age of 14 years[40] using the Oragene® 500 DNA Self-Collection Kit.[22] The MCS data are available to bona fide researchers under standard access conditions via the UK Data Service (**http://ukdataservice.ac.uk**) and the MCS website provides detailed information on the study (**http://www.cls.ioe.ac.uk/mcs**).

The study was ethically approved by an NHS Research Ethics Committee. Informed consent was obtained from the parents, as well as from the children themselves as they grow up. Written consent was required for DNA collection from parents for their own samples and for their child to provide a sample, and the 14-year-old cohort members themselves had to provide verbal consent.[22] Ethical approval (23N.008298) for the analysis of MCS secondary data at the MPI was also obtained from the Ethics Committee Social Sciences (ECSS) at Radboud University.

**Funding:** The MCS is core funded by a consortium of government departments and the three devolved administrations (i.e. the Welsh Government, the Scottish Government and the Northern Ireland Executive), as well as the Economic and Social Research Council funds the Centre for Longitudinal Studies (CLS) Resource Centre (ES/W013142/1), which provides core support for the CLS cohort studies, including the Millennium Cohort Study.

**Role of funders:** The CLS Resource Centre makes MCS data available but does not bear any responsibility for the analysis or interpretation of these data.

**Acknowledgements:** The CLS cohorts, including MCS, are only possible due to the commitment and enthusiasm of their participants, their time and contribution is gratefully acknowledged. We are also grateful to the Centre for Longitudinal Studies (CLS), UCL Social Research Institute, for the use of these data and to the UK Data Service for making them available.

##### The Norwegian Mother, Father and Child Cohort (MoBa) Study

**Cohort profile:** MoBa is a population-based pregnancy cohort study conducted by the Norwegian Institute of Public Health.[25-27] Participants were recruited from all over Norway from 1999 to 2008. The women consented to participation in 40.6% of the pregnancies. The cohort includes approximately 114,500 children, 95,200 mothers, and 75,200 fathers. MoBa is regulated by the Norwegian Health Registry Act. Ethical approval for this work was given by the Regional Committees for Medical and Health Research Ethics (REK) (2016/1702). We also used data from the Medical Birth Registry of Norway (MBRN), a national health registry containing information about all births in Norway.

**Role of funders:** The funders have/had no role in study design, data collection and analysis, the decision to publish, or preparation of the manuscript.

**Acknowledgements:** For generating high-quality genomic data, we thank the Norwegian Institute of Public Health (NIPH), the HARVEST collaboration, the NORMENT Centre at the University of Oslo, the Center for Diabetes Research at the University of Bergen, deCODE Genetics, the Research Council of Norway, the SouthEastern and Western Norway Regional Health Authorities, the ERC AdG, Stiftelsen KG Jebsen, the Trond Mohn Foundation, and the Novo Nordisk Foundation. We are grateful to all the families in Norway who have taken part in this ongoing study.

##### The Netherlands Twin Register (NTR) Study

**Cohort profile:** The Netherlands Twin Register is a longitudinal population-wide cohort.[28-30] The NTR was established by the Department of Biological Psychology, Vrije Universiteit Amsterdam in 1986 and has since then collected survey and experimental data in Dutch twins and their families. Using the Twin-pair as the probands, recruitment in both Adult NTR (ANTR) and Young

NTR (YNTR) initially included parents, and in later years also siblings, spouses and offspring of twins. This has resulted in a database with roughly equal proportions of participants who are and who are not twins. Over the years, a total of 280,569 participants were registered at the NTR, 231,088 of whom are still contactable. The NTR collects data in children and adolescents, i.e. the YNTR and in adults, i.e. the ANTR. This data includes self-report data, survey data supplied by parents or teachers, and data collected in dedicated projects such as cardiovascular and magnetic resonance imaging (MRI) studies (for more information see[30]). Ethical approval for this work was given by the Central Ethics Committee on Research Involving Human Subjects of the University Medical Centers Amsterdam (YNTR3/YNTR5/YNTR7/YNTR10/YNTR12 (94/105, 21-06-1994; 96/205, 14-01-1997; 99/068, 11-08-1999; 2003/182, 18-12-2003; and 2010/359, 18-02-2011).

**Funding:** The Young NTR is and has been funded by a NWO large investment grant (NTR: 480-15-001/674)), Twin family database for behaviour genomics studies (NWO 480–04-004), and Developmental Study of Attention Problems in Young Twins (National Institute of Mental Health, Grant No. RO1 MH58799-03).

**Role of funders:** The funders had no role in the planning or execution of the study nor the interpretation or publication of its results.

**Acknowledgements:** We thank all participants of the Netherlands Twin Register who take part in this on-going study.

**The Pre-, Peri-, and Postnatal Stress: Epigenetic Impact on Depression (POSEIDON) Study**

**Cohort profile:** The POSEIDON study is a longitudinal cohort study comprising five assessment time points: the third trimester of pregnancy (T1), perinatal and a few days postpartum (T2), six months postpartum (T3), 45 months postpartum (T4), and 7-10 years (T5).[31,32] A total of 410 mothers were recruited from hospitals in the Rhine-Neckar Region in Germany between 2010 and 2013. For genetic analyses, parental saliva samples were collected at T1, and EDTA cord blood samples from the newborns were obtained at T2. To compensate for the number of dropouts at T4, the cohort was supplemented with 101 additional children and their parents. At T4, saliva samples were collected from both the new children and their parents for genetic analysis. T5 was conducted as an online survey during the COVID-19 pandemic. The study was registered in the German Clinical Trials Register (DRKS00006338), conducted in accordance with the Declaration of Helsinki, and approved by the Ethics Committee of the Medical Faculty Mannheim, University of Heidelberg. Written informed consent was obtained from all participating families and reaffirmed at T4 and T5.

**Funding:** This work was supported by the German Federal Ministry of Education and Research (BMBF) and the ministry of Badem-Württemberg within the initial phase of the German Center for Mental Health (DZPG) (grant: 01EE2304D), by the BMBF through ERA-NET NEURON (grant: 01EW1904), by grants of the Dietmar-Hopp Foundation, and the by Health + Life Science Alliance, Heidelberg, Mannheim.

**Role of funders:** The funders had no role in the planning or execution of the study nor the interpretation or publication of its results.

**Acknowledgements:** We thank all parents and children who take part in this ongoing study.

**The Prediction and prevention of preeclampsia and intrauterine growth restriction (PREDO) Study**

**Cohort profile:** The PREDO study is a prospective, multi-centre study of Finnish women who were pregnant between 2005 and 2009 and their children.[33] PREDO recruited 1079 women with a singleton, intrauterine pregnancy, who visited antenatal clinics at any of 10 study hospitals for their first routine ultrasound screening at 12 to 13 weeks of gestation, of whom 969 had one or more and 110 had none of known risk factors for preeclampsia and intrauterine growth restriction. All participating mothers provided written informed consent. The study protocol was approved by the Ethics Committee of Obstetrics and Gynaecology and Women, Children and Psychiatry of the Helsinki and Uusimaa Hospital District and by the participating hospitals (487/E7/04). The study has been registered as ClinicalTrials.gov identifier ISRCTN14030412.[41]

**Funding:** The PREDO Study has been funded by the Academy of Finland (J.L.: 269925 and 311617; K.R.: 128789, 1287891, and 1312670), EraNet Neuron, EVO (a special state subsidy for health science research), University of Helsinki Research Funds, the Signe and Ane Gyllenberg foundation, the Emil Aaltonen Foundation, the Finnish Medical Foundation, the Jane and Aatos Erkko Foundation, the Novo Nordisk Foundation, the Päivikki and Sakari Sohlberg Foundation, Juho Vainio foundation, Yrjö Jahnsson foundation, The Finnish Society of Sciences and Letters, Jalmari and Rauha Ahokas foundation, Sigrid Juselius Foundation granted to members of the Predo study board. Methylation assays were funded by the Academy of Finland (269925). Dr. Lahti has received research support from the Strategic Research Council (SRC) established within the Academy of Finland (decision number: 352700).

**Role of funders:** The funders had no role in the planning or execution of the study nor the interpretation or publication of its results.

**Acknowledgements:** The PREDO study would not have been possible without the dedicated contribution of the PREDO study group members: E. Hamäläinen, E. Kajantie, H Laivuori, P.M. Villa, A-K. Pesonen, A. Aitokallio-Tallberg, A-M. Henry, V.K. Hiilesmaa, T. Karipohja, R. Meri, S. Sainio, T. Saisto, S. Suomalainen-Konig, V-M. Ulander, T. Vaitilo (Department of Obstetrics and Gynaecology, University of Helsinki and Helsinki University Central Hospital, Helsinki, Finland), L. Keski-Nisula, M-R. Orden (Kuopio University Hospital, Kuopio Finland), E. Koistinen, T. Walle, R. Solja (Northern Karelia Central Hospital, Joensuu, Finland), M. Kurkinen (Päijät-Häme Central Hospital, Lahti, Finland), P. Taipale, P. Staven (Iisalmi Hospital, Iisalmi, Finland), J. Uotila (Tampere University Hospital, Tampere, Finland). We thank all the PREDO children and their parents for their enthusiastic participation. We also thank all the research nurses, research assistants, and laboratory personnel involved in the PREDO study.

**The TwinLife Study**

Cohort profile: TwinLife study is currently a 10-year longitudinal, cross-sequential study with an extended twin family design, including annual survey data from mono- (MZ) and dizygotic (DZ) same-sex twin pairs and their families (N = 4,096 families at first data collection in 2014) across Germany. A detailed description of the TwinLife study design and survey format is provided.[34-37] The twin sample comprises four age groups with twins born in 2009/2010 (cohort 1), 2003/2004 (cohort 2), 1997/1998 (cohort 3) and 1990-1993 (cohort 4). Each of the four birth cohorts includes two birth years (subsample A and B) interviewed in consecutive years. The first data collection took place in the form of a household interview (face-to-face 1= F2F1) at the age 5, 11, 17, and 23 years from October 2014 to April 2016 followed by a computer-assisted telephone interview (CATI1) from November 2015 to April 2017. The time interval between two data collections is always one year, and participants are interviewed alternately at home and by telephone (i.e., F2F 2 in 2016–2018, CATI 2 in 2017–2019, F2F 3 in 2018–2020, CATI 3 in 2019–2021, and F2F 4 in 2020–2022). From December 2018 to April 2020, TwinLife participants were recruited to donate a first saliva sample for genotyping as part of the TwinSNPs satellite project. As part of the TwinLife Epigenetic Change Satellite, TwinLife participants were recruited to donate a second saliva sample in September 2021 to May 2022.

The TwinLife study was reviewed and approved by the German Psychological Society (Deutsche Gesellschaft für Psychologie; protocol number: RR 11.2009). The Ethics Committee of the Medical Faculty of the University of Bonn (No. 113/18) reviewed and approved the protocols for the genetic sampling via saliva samples. Prior to the genetic sampling, the participants received extensive written information on, e.g., the sampling procedures, the scope of the study, their right to refuse participation, and their right to withdraw their consent at any given time. For sampling genetic data, written informed consent was obtained from all participants and the participant's legal guardian (for minors).

**Funding:**

**Survey data (Scientific Use File)**

The TwinLife study was funded by the German Research Foundation (DFG) (Grant number 220286500). Funding was awarded to Martin Diewald, Christian Kandler, Frank M. Spinath, Bastian Mönkediek, and Rainer Riemann. The funder had no role in the study design, data collection and analysis, the decision to publish, or the preparation of the manuscript.

**TwinSNPs**

The molecular genetic extension project of TwinLife was funded by the German Research Foundation (DFG; Grant number 428902522). Funding was awarded to Martin Diewald, Peter Krawitz, Markus M. Nöthen, Rainer Riemann, and Frank M. Spinath. The funders had no role in the study design, data collection and analysis, the decision to publish, or the preparation of the manuscript.

**TECS**

The epigenetic change satellite project of TwinLife was funded by the German Research Foundation (DFG; Grant number 458609264, https://gepris.dfg.de/gepris/projekt/458609264). Funding was awarded to Elisabeth Binder, Martin Diewald, Andreas J. Forstner, Christian Kandler, Markus M. Nöthen, and Frank M. Spinath. The funders had no role in the study design, data collection and analysis, the decision to publish, or the preparation of the manuscript.

**Role of funders:** The funders had no role in the planning or execution of the study nor the interpretation or publication of its results.

**Acknowledgement:** We would like to express our sincere gratitude to the TwinLife data management team (in particular, Dr. Anita Kottwitz and Dr. Mirko Ruks) for their invaluable support with the TwinLife data and Shirin Zare for performing the DNA extraction. Moreover, the TwinLife study and all its satellites would not have been possible without the dedicated collaboration of all the TwinLife Study group members from the University of Bielefeld, University of Saarland, and University of Bremen as well as the University of Bonn and Max Planck Institute of Psychiatry regarding its molecular satellite projects. We also thank the TwinLife cohort mothers, fathers and children for their dedicated and continued participation.

#### Genotyping and data processing in each cohort

Each cohort genotyped their DNA samples and processed their genotype data internally and processed in Plink 1.9[42,43] and R[44] (https://www.R-project.org). All cohorts met the recommended QC criteria, e.g., single-nucleotide polymorphism (SNP) with minor allele frequency (MAF) > 1%, call rate > 95% (or > 99% if a the SNP had MAF > 5%), Hardy-Weinberg equilibrium (HWE) exact test passing $p > 1e^{-6}$, in autosomal chromosomes, and with low autosomal heterozygosity and imputation quality ($R^2$) ≥ 80% were included; multi-allelic or duplicate SNPs and structural variants (InDels) were removed.

**The Avon Longitudinal Study of Parents and Children (ALSPAC)**

The genotyping of ALSPAC participants performed using the three genotyping platforms, namely: Illumina HumanHap550-quad chip for children, Illumina human 660W-quad for mothers and Illumina HumanCoreExome chip genotyping for fathers. The genotype sample went through a population stratification and only European samples remained in the data.

The genotype underwent a QC step excluding SNPs with MAF < 1%, call rate < 95%, more than 5% missingness or evidence for violations of HWE for each genotyping batch. Cryptic relatedness was measured as proportion of identity by descent (IBD > 0.125). Related subjects were removed. Samples were excluded if they displayed more than 5% missingness, had indeterminate X chromosome heterozygosity or extreme autosomal heterozygosity.

The genotype of mothers and related children, after QC steps and exclusion of related subjects using cryptic relatedness measures described previously, resulted in 8,237 eligible children and 8,196 eligible mothers (465,740 SNPs). The sample underwent imputation using the 1000 Genomes Phase 1 version 3 reference panel. Additionally, 2,911 unrelated partner genotypes at 507,586 SNPs were imputed using the 1000 Genomes Phase 1 version 3 reference panel as a different batch.

**The Generation R (GenR) Study**

A detailed description of the Generation R genotyping procedure has been provided previously.[20,45,46] In summary, children were genotyped using either the Illumina 610k, 660k, or GSA-MD v2.0 SNP array (Illumina, San Diego, CA), while their parents were genotyped using either the GSA-MD v2.0 or GSA-MD v3.0 array.

The samples underwent QC checks including assessments for HWE, sample and SNP call rates, excess heterozygosity, and mismatches between genetically derived and reported sex or relatedness. For all participants imputation was performed using the 1000 Genomes Project reference (Phase 3, Version 5). Post-imputation QC was performed to assess common autosomal

SNPs with high imputation quality ($R^2 \geq 0.80$) and a MAF of at least 1%. This process resulted in 7,673,245 SNPs (Illumina 610k, 660k) and 9,508,795 SNPs (GSA-MD v2.0) for children, and 9,520,497 SNPs for parents, which were used in subsequent PGS computations. Ten genomic components were calculated using multidimensional scaling (MDS).

**The Millennium Cohort Study (MCS)**

Genotyping was performed using the Infinium Global Screening Array-24 v.1.0, genotypes were called using Genome Studio v2.0.4 graphical user interface,[22] and QC was applied using PLINK (v1.9b6). Individuals were excluded in case of a mismatch between reported and genetic sex information, > 3% missing SNP information, > 3 standard deviations (SD) in heterozygosity rate, mendelian errors (maximum error rate of 0.2% per trio and per variant), and non-European ancestry (informed by MDS analyses). In addition, relationships between different parent-child families were restricted to IBD < 0.125. SNPs were excluded if they had a low call rate (< 95%), were rare (< 0.5%) and/or deviated from HWE ($p < 5 \times 10^{-7}$). Imputation was carried out with the Sanger imputation server (https://imputation.sanger.ac.uk), using the EAGLE2[47] and PBWT imputation pipeline for pre-phasing, and the HRC r1.1 reference panel[48]. Subsequently, best-guess genotypes were constructed for high-quality imputed SNPs (INFO > 0.8; 95%-posterior genotyping probability > 0.9; MAF > 0.5%). This resulted in 6,947,751 autosomal markers for 2,517 complete trios (irrespective of whether SDQ data is available).

**The Norwegian Mother, Father and Child Cohort (MoBa) Study**

Blood samples were obtained from both parents during pregnancy and from mothers and children at birth.[26] The genotyping and QC procedures have been described elsewhere.[49]

**The Netherlands Twin Register (NTR) Study**

Genotyping of NTR DNA samples was done on 6 SNP array platforms, namely Affymetrix 5-Perlegen (N = 1904), Affymetrix 6.0 (N = 10377), Affymetrix Axiom (N = 3536), Illumina Omni

1M (N = 445), Illumina Human Quad Array 660W (N = 1501) and Illumina GSA NTR array (N = 20060). Genotype calling was done following the manufacturers protocols and white papers. Older genome builds 35 and 36 of Illumina Omni, 660 and Affymetrix-Perlegen were lifted-over to build 37.

For each individual platform, DNA samples were checked for gender mismatches, heterozygosity with Plink F value being between -0.10 to 0.10 and Plink estimated IBD mismatches in comparison to the known family structure. For call rate in the samples, each sample needed to have at least 90% genotyped and furthermore, at least 80% of the genotypes needed to be present on each separate chromosome 1-22 plus X for each person. Problematic samples were removed given all the above criteria. In addition, multiple typed samples were removed from the platforms, the first non-problematic sample was taken for subsequent steps. The SNP QC was based on the following filters applied in each platform: call rate should be over 95%, HWE *p*-value should be over 0.0001, MAF should be over 0.01 and Mendelian error rate should be less than 1%. Based on several plate control samples in Affymetrix 6 (N = 4 typed 38-84 times) and Axiom (N = 2 typed 33-37 times) and the at least 2 times typed samples mentioned above in the separate platform sets, SNPs were removed if the genotypes differed more than 1% between these multiple measurements.

The GONL reference panel 5.4 was used for SNP name - and positive strand alignment on build 37 (HG19) for each platform. If the platform allele frequency was more than +/-0.10 the reference allele frequency, SNPs were removed. Finally, palindromic SNPs with an allele frequency of 0.35-0.65 were also removed. After this alignment of the data, the platforms are also inherently aligned with each other. Therefore, at this point the data of the 6 platforms were merged into a single dataset and based on the remaining overlapping SNPs (N = 7478), IBD was checked again against the known family structure (now across platforms). Mismatching samples and samples which had discordant DNA across platforms were also removed in this step.

In a recent effort nearly all samples, except 44, of the older platforms being Affymetrix-Perlegen, Illumina 660 and Omni 1M have been retyped on the Illumina GSA array. Therefore, after applying the above QC steps to filter cross platform bad samples, the 3 older platforms were not used further. As these platforms are also informative about the monozygotic state of twins, the DNA of these individuals was duplicated from twin one to twin two for 1024 individuals in the GSA, Affymetrix 6.0 and Axiom arrays if needed. Then over platforms, keeping both MZ twins on a single array, samples were selected for each participating individual in the following order of arrays: Axiom, Affymetrix 6.0 and then Illumina GSA. Invalid NTR samples (controls), withdrawn consents and overlapping samples across the 3 arrays were removed at this point. This led to the final set of 3644 individuals with 534405 SNPs on Axiom, 9049 individuals with 537992 SNPs on Affymetrix 6.0 and 16276 individuals with 481898 SNPs on Illumina GSA, a total of 28969 individuals.

Before imputation, for each of the 3 genotype platforms separately, SNP name and reference allele were aligned now to the reference panels 1000 genomes Phase III v5 and HRC 1.1, and data were converted to VCF format with Plink. The data were then imputed with Beagle 5.4. Since GPDR restricts us from using imputation servers, the HRC panel of the Ega Sanger website was used. This panel misses the Sardina, Gecco and GONL cohorts. After imputation the resulting VCF data of the 3 platforms were merged into single chromosome sets 1-22 plus X using BCFtools for each reference panel. With QCtool version 2.20 these data were then also converted to BGEN format, as well as best guess genotypes using Plink 1.9.

Twenty 1000 genomes projected principal components (PCs) for the genotype data were calculated using the EIGENSTRAT smartpca tool. We selected the genotyped SNPs that passed QC present in one of the three platforms from the 1000 Genomes imputed data (as the overlap between platforms is too small to take only genotyped SNPs). These SNPs were then filtered to have MAF > 0.05, HWE $p > 0.001$, call rate > 0.98, Mendelian error rate < 1% and imputation info >= 90%. These SNPs were subsequently pruned with plink (option –indep 50 5 2) and SNPS in long range LD blocks were removed. This left 110558 SNPs for analysis. From the 1000 genomes reference panel all samples with the same SNPs were selected and then merged with the NTR data. Subsequently PCs were calculated in the 1000 genomes set and then projected upon the NTR data with the smartpca software.

For polygenic scoring we used the NTR data imputed to the HRC reference panel. Prior to starting with scoring also here a post-imputation SNP QC selection was employed. This included the following SNP filters: MAF > 0.01, HWE $p > 0.00001$, Mendel error rate < 1% and genotype call rate over 98%. This selection was made on the merged best guess 3 platforms data. Furthermore, the imputation info for the 3 individual platforms all needed to be above 0.

#### The Pre-, Peri-, and Postnatal Stress: Epigenetic Impact on Depression (POSEIDON) Study

Genome-wide genotyping was carried out using the Illumina Infinium Psych Array and the Illumina Global Screening Array (Illumina, Inc., San Diego, CA). Prior to genotype imputation, standard QC procedures were applied using the RICOPILI pipeline (Rapid Imputation and COmputational PIpeLIne for Genome-Wide Association Studies, from Broad Institute). QC steps included the following filters: SNP missing rate < 0.05 (before filtering individuals), individual missingness < 0.02, removal of individuals with sex mismatches, autosomal heterozygosity deviation Fhet| < 0.2, SNP missing rate < 0.02 (after filtering individuals), MAF > 0.01, HWE p $\geq 1e^{-6}$, and exclusion of SNPs lacking valid association p-value. Genotype imputation was then performed using the RICOPILI imputation pipeline with the 1000 Genomes reference sample. Post-imputation, both parental and offspring datasets were further filtered to retain variants with SNP-missing rate < 0.02 and INFO score ≥ 0.8. A subset of autosomal SNPs, filtered for quality (MAF > 0.2, HWE $p > 0.02$, SNP missingness = 0), was used for relatedness check and principal component analysis (PCA). Linkage disequilibrium (LD) pruning was performed by removing SNPs with pairwise $R^2 > 0.05$ within sliding windows of 250 SNPs. Individuals showing a pairwise IBD estimate (Pi Hat) > 0.1 were excluded to remove related samples. To identify and exclude genetic outliers, individuals exceeding six SDs on any of the first 20 PCs were removed.

#### The Prediction and prevention of preeclampsia and intrauterine growth restriction (PREDO) Study

Genotyping was performed with DNAs extracted from the full blood samples in both parents using IlluminaGlobal Screening array (Illumina Inc, San Diego, CA) and the cord blood samples in the offspring using Illumina Human Omni Express Exome Arrays according to the manufacturer's guidelines (Illumina Inc., San Diego, CA). QC was performed in Plink 1.9 and R. In the pre-imputation QC, samples were excluded if genotype call rate < 95%, heterozygosity F < -0.1, sex mismatch between phenotyping and genotyping. If relatedness IBD > 0.125 between any pair of samples, one sample was excluded from further analysis. Population outliers were excluded based on visual inspection. SNP imputation was performed using IMPUTE v2.3.2 and Eagle v2.3 against the Finnish specific SiSu v2 reference panel (GRCh37) comprising 2690 high coverage whole-genome and 5093 high-coverage whole-exome sequences, which introduces less false polymorphisms than using global reference panels.[50] SNPs were removed if genotype call rate < 95%, MAF > 0.35, minor allele count < 19, HWE $p < 1 \times 10^{-6}$, outside autosomes non-PAR region of the chromosome X, ambiguous strand when compared to the imputation reference, chromosome X variants with heterozygous male calls, or with imputation INFO score < 0.8. There were 15,536,178 (of which 82.4% with INFO > 0.8) and 15,544,584 (of which 86.1% with INFO > 0.8) variants after imputation in the offspring and any parental samples, respectively. For ancestry-related information, MDS analysis on the identity by the IBS state matrix of QCed genotypes was performed to adjust for population structure.[51] The first ten PCs were extracted and included as covariates in statistical analyses

**The TwinLife Study**

Genotyping and data processing of the TwinLife study have been described in detail elsewhere.[52] In summary, saliva samples for DNA extraction were collected as part of both molecular genetic TwinLife satellite projects, TwinSNPs and TECS. Twins and their family members were invited to provide saliva samples for the above-mentioned satellite projects during TwinLife face-to-face interviews and by mail. Respondents were provided with informed consent forms, Oragene® saliva self-collection kits (OG-500 or OG-600; DNA Genotek, Canada), and instructions.

DNA was extracted according to standard procedures. Genome-wide genotyping was conducted using Global Screening Arrays (GSA+MD-24v3.0-Psych-24v1.1, Illumina, San Diego, CA, USA), customized to include additional markers relevant to psychiatric disorders, according to the manufacturer's protocol.

The QC of the genotype data was performed using PLINK and R. Raw genotype data were available for 5,927 samples resulting in 5,861 individuals after filtering based on a high overall genotyping rate of 98% and autosomal heterozygosity deviation (FHET) within +/- 0.20. Variants with a call rate < 98%, a deviation from HWE with a p-value $< 1 \times 10^{-6}$ among unrelated individuals (considering one sample per family selected according to the highest genotyping rate) or a MAF < 0.001 were removed from the dataset. Post-QC data were phased using Eagle (v2.4.1)[53] and imputed using Minimac (v4)[47] considering the 1000 Genomes Phase 3 data (N = 2,504)[54] as the reference panel after removing ambiguous variants (A/T, C/G) to avoid potential strand issues alignment between the raw genotypes and the reference haplotypes. From the post-imputation data, only variants with an imputation accuracy ($R^2$ imputation) > 0.8 were retained for further analyses.

Sex checks were performed using PLINK 1.9 genetically inferred sex and self-reported sex. Samples with undetermined genetically inferred sex and samples with a mismatch between the genetically inferred and self-reported sex were excluded.

As the TwinSNPs sample consists of twins and their family members, a relatedness check was included in the QC. For this purpose, KING[55] was used to compute the kinship coefficients between samples and infer familial relationships from genotype data. Samples with a discordant genetically inferred family structure (reflected by pairwise KING relatedness coefficients and KING-generated family IDs) with respect to reported relatedness (reflected by the fid and ptyp SUF variables) were excluded, except for zygosity mismatches. We retained monozygotic pairs with an inferred genetic zygosity discordant with the reported zygosity if the genetically inferred family structure was otherwise consistent with the self-report. In these cases, the zygosity variable was corrected to include the genetically inferred zygosity. Ten genetic PCs were derived by projecting the samples into the PC space defined using the 1000 Genomes Phase 3 reference panel and used to flag PC outliers, defined as samples whose PC values deviated more than 6 SDs from the mean of the first 10 PCs.

#### Polygenic score (PGS) calculation in each cohort

##### Genome-wide association study (GWAS) summary statistics and QC

The largest available GWAS summary statistics used in the current study included attention deficit/hyperactive disorder (ADHD),[56] anorexia nervosa (AN),[57] anxiety disorder (ANX),[58] autism spectrum disorder (ASD),[59] bipolar disorder (BD),[60] cross-psychiatric disorders (CDG),[61] insomnia (INSOM),[62] major depressive disorder (MDD),[63] neuroticism (NEUROT),[64] obsessive compulsive disorder (OCD),[65] postpartum depressive (PPD),[66] post-traumatic stress disorder

(PTSD),[67] and schizophrenia (SCZ).[68] General mental health traits included substance use (e.g., daily consumption of alcohol (ALC) or cigarettes (CIG)[69] and EA.[70]

**PGS calculation**

PGSs were calculated based on the above mentioned GWAS summary statistics with computed SNP weights using extensive sets of Hapmap3+ SNPs as the reference panel in LDpred2-auto,[71] after the QC of the GWAS summary statistics. The QC procedure involved removal of variants based on their SDs of allele frequencies. Specifically, SDs inferred from the GWAS summary statistics were compared with those computed from a reference panel (e.g., UKB), and variants with their SDs either above or below the expected value were removed.[72]

PGSs from children (PGSc), mothers (PGSm), and fathers (PGSf) were calculated based on the respective QCed GWAS summary statistics using imputed plink bed files by PLINK (v1.9b.6)[42,43] that directly estimated two key model parameters from genotype data: the SNP heritability and polygenicity. These SNP weights were used to calculate the summation scores.

Extreme outliers in PGSs were winsorized according to the 3* interquartile range (IQR), non-transformed PGSs were used and standardized with mean of 0 and the SD of 1. Winsorization has been shown to greatly reduce false positives by popular differential expression methods when analyzing human population samples.[73]

**The Avon Longitudinal Study of Parents and Children (ALSPAC)**

PRSc, PRSm, and PRSf were calculated.

**The Generation R (GenR) Study**

PRSc, PRSm, and PRSf were calculated.

**The Millennium Cohort Study (MCS)**

PRSc, PRSm, and PRSf were calculated.

**The Norwegian Mother, Father and Child Cohort (MoBa) Study**

PGSc, PGSm, and PGSf were calculated. For BD, EA, and PPD, SNP weights were recomputed excluding MoBa from the discovery GWASs before calculating the PGSs.

**The Netherlands Twin Register (NTR) Study**

PGSc, PGSm, and PGSf were calculated. PGSs for ALC, CIG, and NEUROT were calculated using re-computed summary statistics excluding the NTR from the discovery GWASs, and PGS for ANX was not calculated since NTR was a major part of the discovery GWAS.

**The Pre-, Peri-, and Postnatal Stress: Epigenetic Impact on Depression (POSEIDON) Study**

PGSc, PGSm, and PGSf were calculated.

**The Prediction and prevention of preeclampsia and intrauterine growth restriction (PREDO) Study**

PGSc, PGSm, and PGSf were calculated.

**The TwinLife Study**

PGSc, PGSm, and PGSf were calculated.

#### Child behavioral phenotyping and covariates in each cohort

##### Behavioural measure instruments: Child Behaviour Check List (CBCL) and Strengths and Difficulties Questionnaire (SDQ)

CBCL for ages 1½ to 5 years (CBCL/1½ - 5) and 6 to 18 years (CBCL/6 - 18) were filled in by the child's mother or father. CBCL comprises 99 problem items rated on a scale of not true (0) to very true or often true (2).[74] The CBCLs has good test-retest reliability, internal consistency, and criterion validity.[74,75] The CBCLs yield scores for three main scales (internalizing, externalizing, and total difficulties), seven syndrome scales (emotionally reactive, anxious/depressed, somatic complaints, withdrawn, sleep difficulties, attention difficulties, and aggressive behaviour), and five DSM-oriented scales (affective, anxiety, pervasive developmental, attention-deficit/hyperactivity, and oppositional defiant problems). Total emotional/behavioural difficulties are reflected by a summative total CBCL t-score, where higher value indicates greater difficulties; Externalizing difficulties constellate a summative t-score of subscales for impulsivity, aggressiveness, and conduct difficulties; Internalizing difficulties encapsulate a summative t-score of subscales for emotional withdrawal, anxiety, and depression. A categorical CBCL variable can also be created by dichotomizing the total difficulties t-scores at a cutoff value of 60. A t-score of ≥ 60 on the main scale indicates borderline clinically significant difficulties.

SDQ for ages 2 to 4 years (SDQ2 - 4) and 4-17 years (SDQ4 - 17) were filled in by the child's mother or father.[76,77] Total emotional/behavioural difficulties are assessed using a summative total SDQ score, where higher value indicates greater difficulties; Externalizing difficulties are assessed by combining the inattention/hyperactivity subscale and the conduct difficulties subscale scores; Internalizing difficulties are assessed by combining the emotional symptoms subscale and the peer difficulties subscale scores. A categorical SDQ variable can also be created by dichotomizing the total difficulties scores at a cutoff value of 14. A total SDQ score ≥ 14 indicates elevated symptom levels, while a total SDQ score ≥ 17 is consistent with high/very high clinical risk.

Child's age at assessment point, child sex, maternal age at childbirth, maternal educational level at childbirth or child assessment, based on the US standard (UNESCO UIS),[78] as well as the 10 first genetic PCs calculated from the child's genotype data were included as covariates. The maternal EA was covaried as it has previously been shown to partly explain IGEs for children's academic achievement and behavioural difficulties independent of the immediate environment of the nuclear family.[79-81]

Extreme outliers were winsorized according to the 3*IQR, non-transformed outcome variables and covariates were standardized to the mean of 0 and the SD of 1.

**The Avon Longitudinal Study of Parents and Children (ALSPAC)**

The Strengths and Difficulties Questionnaire (SDQ) was completed by mothers for children aged 9 years.[18] The internalizing score was calculated as the sum of the emotional and peer difficulties subscales, while the externalizing score was calculated as the sum of the hyperactivity and conduct difficulties subscales. A categorical SDQ variable was created, classifying children as cases (1) if the total difficulties score was greater than the cutoff value of 14, and as controls (0) otherwise.

**The Generation R (GenR) Study**

Fathers and mothers completed the CBCL/1½–5 and CBCL/6–18 to report child behaviours. T-scores for internalizing, externalizing, and total difficulties in children were computed. Both continuous T-score variables and a binary variable for total difficulties (0 = CBCL < 60; 1 = CBCL ≥ 60) at ages 3 and 9 years were created for analysis.

**The Millennium Cohort Study (MCS)**

SDQs filled by one of the parents (predominantly by mothers, whose exact information was unavailable to us) at a child age of three[23] and eleven[24] years were included in the current study. Difficulties scores were only computed for children with complete data on the required scales (e.g. only children with data on both emotional symptoms and peer relationships difficulties have a score for internalising difficulties). A categorical SDQ variable was created by dichotomising the total difficulties score at a cutoff value of 14.

#### The Norwegian Mother, Father and Child Cohort (MoBa) Study

In MoBa, a short version of the CBCL was used. The total problem score was based on all available CBCL items from the 3-year (26 items) and 5-year (26 items) questionnaires, respectively.[25] In all cases, mothers responded to items on the CBCL using a 3-point scale ranging from "Not true" to "Often true" rating their children's behavioural and emotional difficulties. The behavioural difficulties subscale (11 items from both questionnaires) captures aggressive behaviour and attention difficulties. The emotional difficulties subscale (9 items from 3-year and 11 items from 5-year questionnaire) assesses emotional reactivity and anxiety/depression. Since it was based on a short version of the CBCL, standardised raw scores (rather than T-scores) were used as outcomes. MoBa was not included in the analysis based on the categorised total score for the same reason. The 896 trios included in the CBCL/6-18 analyses were those who returned the 5-year questionnaire after age 6. Anyone responding after age 6 were excluded from the CBCL/1-5 analyses.

If participants ticked multiple boxes on a single-response item, their response on this item was set to be missing. If respondents completed less than half of the items for a given scale, their scale score was not computed and was considered missing.

In MoBa, genotyping batches were included as technical covariates (dummy coded) in addition to the covariates described above. We also did not exclude related individuals. Instead, we included mother ID as a clustering variable to account for siblings as the main source of relatedness.

#### The Netherlands Twin Register (NTR) Study

CBCL/1½ - 5 CBCL/6 - 18 t-scores for internalizing, externalizing, and total difficulties were available for the age groups 3 and 10 years, both mother- and father-reported in NTR.[29] Both continuous t-score variables and a binary variable of child total difficulties t-score (e.g., 0 = CBCL $< 60$, 1 = CBCL $\geq 60$) were created.

**The Pre-, Peri-, and Postnatal Stress: Epigenetic Impact on Depression (POSEIDON) Study**

CBCL t-scores for internalizing, externalizing, and total difficulties were available for the age group 1.5-5 years (average 3.8 years, mother-reported). Both continuous t-score variables and a binary variable of child total difficulties t-score (e.g., 0 = CBCL < 60, 1 = CBCL ≥ 60) were created.

SDQ summative raw scores for internalizing, externalizing, and total difficulties were available for the age group 6-17 years (average 7.7 years, mother-reported). Both continuous score variables and a binary variable of child total difficulties score (e.g. 0 = SDQ < 14, 1 = SDQ ≥ 14) were created.[82] One extreme outlier in the SDQ score for internalizing difficulties and one for total difficulties were winsorized according to the 3*IQR, non-transformed CBCL t-scores and SDQ total scores were used and standardized.

**The Prediction and prevention of preeclampsia and intrauterine growth restriction (PREDO) Study**

CBCL t-scores for internalizing, externalizing, and total difficulties were available for the age group encompassing 1.5-6 years (average 3.5 years, mother-reported) and 7-12 years (average 9 years, both mother- and father-reported) in PREDO. Both continuous t-score variables and a binary variable of child total difficulties t-score (e.g., 0 = CBCL < 60, 1 = CBCL ≥ 60) were created.[33]

**The TwinLife Study**

SDQ was collected during the face-to-face (F2F) interviews.[37] For children < 10 years of age at F2F1, SDQ items were filled in by either the mother or the father. A total SDQ score was calculated using the sum of all available SDQ items for each participant. SDQ total scores for internalizing, externalizing, and total difficulties were available for the age group encompassing 4-5 years (mother- or father-reported) in TwinLife. A categorical SDQ variable was created by dichotomizing the total difficulties scores at a cutoff value of 14.

#### Methods used in cohort-level statistical analyses

All cohorts used the same scripts in R[44] for statistical analyses. Four regression models were set up:

**M1** and **M3** were linear and logistic regression models for single PGS predictors, respectively:

$Yc \sim iXi+covXcov$, where $Yc$ was the child's behavioural outcome, $Xi$ was an independent PGS predictor, and $Xcov$ were covariates.

**M2** and **M4** leveraged trio-PGSs as predictors of each outcome for linear and logistic regression models, respectively:

$Yc \sim \beta cXc+mXm+fXf+covXcov$, where $Yc$ was the child's behavioural outcome, $Xc$, $Xm$, and $Xf$ were PGSc, PGSm, PGSf predictors, respectively. $Xc$ reflects direct genetic effect (DGE), whereas $Xm$ and $Xf$ reflect parental indirect genetic effect (IGE). $Xcov$ were covariates.

Multicollinearity among predictors and covariates were tested and variance inflation factors (VIFs) were examined for each model, and all VIFs were $< 10$.

Pearson's correlation coefficients among the trio-PGSs as well as between mother- and father-reported scores for each behavioural outcome in applicable cohorts were also calculated.

#### Methods used in meta-analyses

Cohort-level summary statistics were further used for meta-analyses. Our main meta-analysis model was multivariate meta-analysis (MVMA) pooling results across cohorts and raters. For the pre-school-aged group, GenR3, GenR4, MCS, MoBa, NTR, POSEIDON, PREDO and TwinLife were included in MVMA. For the school-aged group, ALSPAC, GenR3, GenR4, MCS, MoBa, NTR, POSEIDON, and PREDO were included in MVMA. The simultaneous analysis of both maternally and paternally rated psychopathology has the advantage of lessening potential biases due to shared-rater variance, as well as higher statistical power due to increased rating accuracy. To account for the correlation between maternal and paternal ratings, a variance-covariance matrix incorporating cross-rater Pearson's correlation coefficients between mother-reported or father-reported outcomes was used to adjust the inverse variance (IV)-based coefficients. In addition, a random effect (RE) method was used to account for heterogeneity between cohorts. Meta-analysis models were fitted using the rma.mv function in the *metafor* package in R.[83]

For verification of the multivariate models and to obtain informant-specific coefficients, we additionally conducted simpler traditional RE univariate meta-analyses (UVMA) on either mother-reported or father-reported outcomes using the *metafor* rma.uni function.[83] For mother-reported outcomes at pre-school age, GenR3, GenR4, MoBa, NTR, POSEIDON, PREDO and TwinLife were included in UVMA. For mother-reported outcomes at school age, ALSPAC, GenR3, GenR4, MoBa, NTR, POSEIDON, and PREDO were included in UVMA. For father-reported outcomes at pre-school age, GenR3, GenR4, NTR, and TwinLife were included in UVMA. For father-reported outcomes at school age, GenR3, GenR4, NTR, and PREDO were included in UVMA. Standardized beta coefficients and confidence intervals (CI) were used to evaluate overall predictive effects. For logistic regressions, cohorts having a sample size < 10 in the high-risk group were excluded in meta-analyses. All reported *p*-values are two-sided and were corrected for multiple testing using the Benjamini-Hochberg False Discovery Rate (FDR) among each set of the 18 predictors (*i.e.*, 18 tests) for child, mother, and father, respectively. The $I^2$ statistic was used to evaluate cohort heterogeneity. Meta-analyses were conducted at the University of Helsinki, and shadow meta-analyses were conducted independently at the Norwegian Institute of Public Health for result validation.

#### Evidence of parental genetic effects in univariate meta-analysis (UVMA)

UVMA was conducted to estimate separately either maternal (N=27828 preschool-age/5669 school-age from 7 European cohorts) or paternal reporters (N=3131 preschool-age/2820 school-age from 4 European cohorts) on child behaviours (tables S11-S26).

##### Genetic liability for general psychopathology

At preschool age, maternal IGEs of general psychopathology were found in mother-reported internalizing (PGSm-CDG: β=0·024 [95% CI 0.007 to 0.040]; $p_{\mathrm{FDR}}$=0·037; Maternal Psych-Mean: β=0·025 [95% CI 0.008 to 0.042]; $p_{\mathrm{FDR}}$=0·037; Maternal Psych-PC1: β=0·026 [95% CI 0.009 to 0.043]; $p_{\mathrm{FDR}}$=0·037; table S19) and total difficulties (PGSm-CDG: β=0·025 [95% CI 0.008 to 0.042]; $p_{\mathrm{FDR}}$=0·020; Maternal Psych-Mean: β=0·025 [95% CI 0.008 to 0.043]; $p_{\mathrm{FDR}}$=0·020; Maternal Psych-PC1: β=0·029 [95% CI 0.012 to 0.047]; $p_{\mathrm{FDR}}$=0·008; table S19). In contrast, father-reported data yielded weaker and insignificant estimates (table S20). Like at preschool age, maternal Psych-PC1 was associated with mother-reported internalizing difficulties at school age (β=0·056 [95% CI 0.017 to 0.095]; $p_{\mathrm{FDR}}$=0·029; table S21), but not with father-reported outcomes (table S22).

**Genetic liability for emotional disorders**

At preschool age, PGSm-MDD's association with child's internalizing difficulties was consistent between mother reporters (β=0·027 [95% CI 0.005 to 0.048]; $p_{\mathrm{FDR}}$=0·052; table S19) and father reporters (β=0·029 [95% CI -0.048 to 0.105]; $p_{\mathrm{FDR}}$=0·95; table S20), although neither reaching statistical significance. Nevertheless, at preschool age, PGSm-MDD was significantly associated with mother-reported internalizing (β=0·057 [95% CI 0.020 to 0.094]; $p_{\mathrm{FDR}}$=0·025; table S21) and total difficulties (β=0·062 [95% CI 0.025 to 0.099]; *pFDR*=0·010; table S21). Although not reaching significance, maternal IGEs were similarly seen in father-reported internalizing (β=0·037 [95% CI -0.021 to 0.095]; $p_{\mathrm{FDR}}$=0·515; table S22) and total difficulties (β=0·060 [95% CI 0.007 to 0.112]; $p_{\mathrm{FDR}}$=0·144; table S22). Similarly to MDD, PGSm-PPD was significantly associated with mother-reported internalizing (β=0·059 [95% CI 0.021 to 0.096]; $p_{\mathrm{FDR}}$=0·025; table S21) and total problem (β=0·067 [95% CI 0.030 to 0.104]; $p_{\mathrm{FDR}}$=0·007; table S21), although not with father-reported ones (both $p_{FDR}$>0.05; table S22).

**Genetic liability for neurodevelopmental conditions**

In preschool-aged children, PGSm-ASD showed significant associations with mother-reported child externalizing (β=0·032 [95% CI 0.016 to 0.049]; $p_{\mathrm{FDR}}$=0·002; table S19) and total difficulties (β=0·031 [95% CI 0.014 to 0.047]; $p_{FDR}$=0·005; table S19) (Figure 2), but much weaker and insignificant effects with father-reported externalizing (β=0·006 [95% CI -0.044 to 0.056]; $p_{FDR}$=0·820; table S20) and total problem (β=0·007 [95% CI -0.041 to 0.055]; $p_{FDR}$=0·993; table S20). At school age, PGSm-ASD showed no significant effects on neither mother-reported nor father-reported outcomes (all $p_{FDR}$>0.05; tables S21-S22).

#### Evidence of parental genetic effects on clinically relevant child behavioral difficulties

We also performed logistic regression analysis of the clinical threshold-relevant dichotomized total difficulties in preschool- and school-aged children and meta-analysed all available cohorts by MVMA (tables S5-S6 for single PGS, and S9-S10 for trio-PGSs) and UVMA (tables S15-18 and S23-S26). The MVMA results on the trio-PGSs model are described below.

Similar to the linear regression results, maternal psychiatric PC1 and mean scores as well as PGSs for MDD and PPD showed significant IGEs in school-aged children (**MDD**: β=0·133 [95% CI 0.042, 0.223]; $p_{FDR}$=0·024; **PPD**: β=0·114 [95% CI 0.024 to 0.204]; $p_{FDR}$=0·047; **Psychiatric PC1**: β=0·155 [95% CI 0.060 to 0.250]; $p_{FDR}$=0·023; **Psychiatric Mean**: β=0·128 [95% CI 0.034 to 0.223]; $p_{FDR}$=0·035; table S10). By contrast, no significant IGE was found in preschool-aged children (table S9).

PGSc-ADHD showed a DGE at preschool (β=0·245 [95% CI 0.081 to 0.409]; $p_{FDR}$=0·038; table S9) but not school age (table S10). Meanwhile, PGSc-EA demonstrated a protective DGE at both ages (preschool: β = -0.172 [95% CI -0.291 to -0.054]; $p_{FDR}$=0·038; table S9; school age: β = -0.259 [95% CI -0.380 to -0.139]; $p_{FDR}$<0.001; table S10). Intriguingly, PGSm-EA showed an enhancing IGE on behavioral difficulties in school-aged children (β=0·157 [95% CI 0.055 to 0.259]; $p_{FDR}$=0·023; table S10).

#### Sensitivity analysis of multivariate meta-analysis (MVMA)

Since ALSPAC paternal genotype data covered only exome variants, we conducted a sensitivity analysis excluding this cohort to ensure the robustness of our findings. While there were slight attenuations in the effect sizes compared to the MVMA results including the ALSPAC, the overall pattern was held for school-aged children.

The key DGEs from child PGSs remained mostly consistent. These include the enhancing effects from PGSc-ADHD, PGSc-MDD, child Psych-Mean, and child Psych-PC1, as well as the protective effect of PGSc-EA on externalizing difficulties (tables S27-S28) (PGSc-ADHD: β=0·111 [95% CI 0.079 to 0.144]; $p_{FDR}$<0.001; PGSc-MDD: β=0·067 [95% CI 0.035 to 0.099]; $p_{FDR}$<0.001; Psych-Mean: β=0·063 [95% CI 0.022 to 0.103]; $p_{FDR}$=0·008; Psych-PC1: β=0·078 [95% CI 0.040 to 0.115]; $p_{FDR}$<0.001; PGSc-EA: β=-0.096 [95% CI -0.131 to -0.060]; $p_{FDR}$<0.001) and on total difficulties (PGSc-ADHD: β=0·064 [95% CI 0.026 to 0.103]; *pFDR*=0·006; PGSc-MDD: β=0·059 [95% CI 0.024 to 0.095]; $p_{FDR}$=0·006; Psych-PC1: β=0·058 [95% CI 0.015 to 0.102]; $p_{FDR}$=0·040; PGSc-EA: β = -0.102 [95% CI -0.134 to -0.070]; $p_{FDR}$<0.001).

None of the maternal IGEs reached statistical significance (table S27). However, the direction of the effects was consistent with previous findings, though the effect sizes were slightly attenuated for internalizing difficulties (PGSm-MDD: β=0·045 [95% CI 0.010 to 0.079]; $p_{FDR}$=0·052; PGSm-PPD: β=0·042 [95% CI 0.010 to 0.074]; $p_{FDR}$=0·052; Psych-PC1: β=0·050 [95% CI 0.013 to 0.087]; $p_{FDR}$=0·052) and total difficulties (PGSm-MDD: β=0·047 [95% CI 0.015 to 0.078]; $p_{FDR}$=0·056; PGSm-PPD: β=0·039 [95% CI 0.007 to 0.070]; $p_{FDR}$=0·072; Psych-PC1: β=0·043 [95% CI 0.011 to 0.076]; $p_{FDR}$=0·056).

In logistic regression (table S28), the associations of PGSc-EA and PGSm-EA with dichotomized child total difficulties remained (PGSc-EA: β =-0.288 [95% CI -0.411 to -0.164]; $p_{FDR}$<0.001; PGSm-EA: β=0·178 [95% CI 0.072 to 0.283]; $p_{FDR}$=0·018), showing even stronger effect sizes compared to the prior findings.

#### Supplementary figures and legends

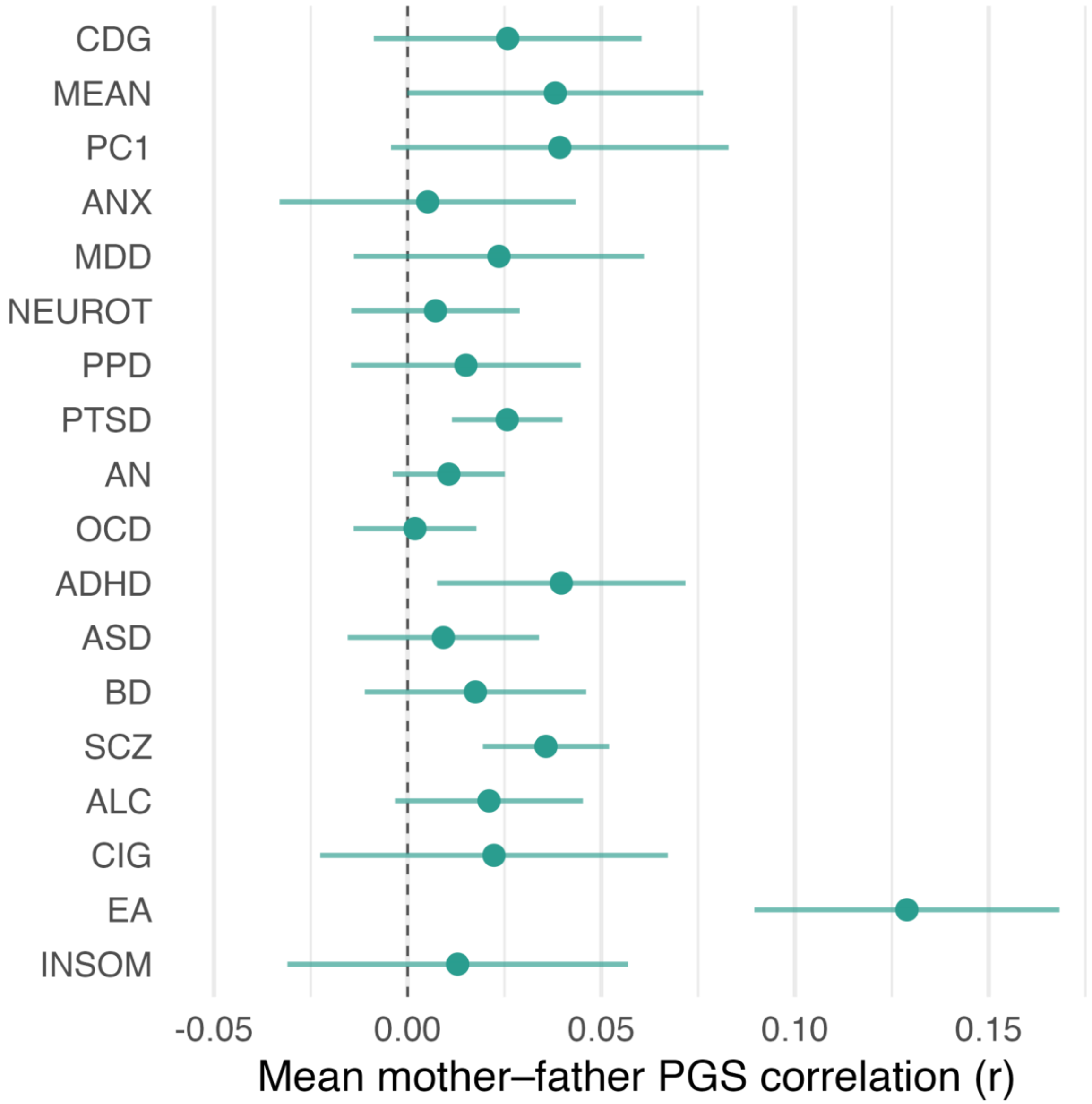


**Figure S1. Mother-father polygenic score correlations (assortative mating) across cohorts.**

Within each cohort, we retained same-trait mother–father polygenic score (PGS) correlations (Pearson's) at the youngest available child age. Points show the *N*-weighted mean correlation across cohorts; error bars indicate 95% CIs based on the cross-cohort variability (SD/√k). All cohorts contributed (ALSPAC, GenR, MCS, MoBa, NTR, POSEIDON, PREDO, and TwinLife).

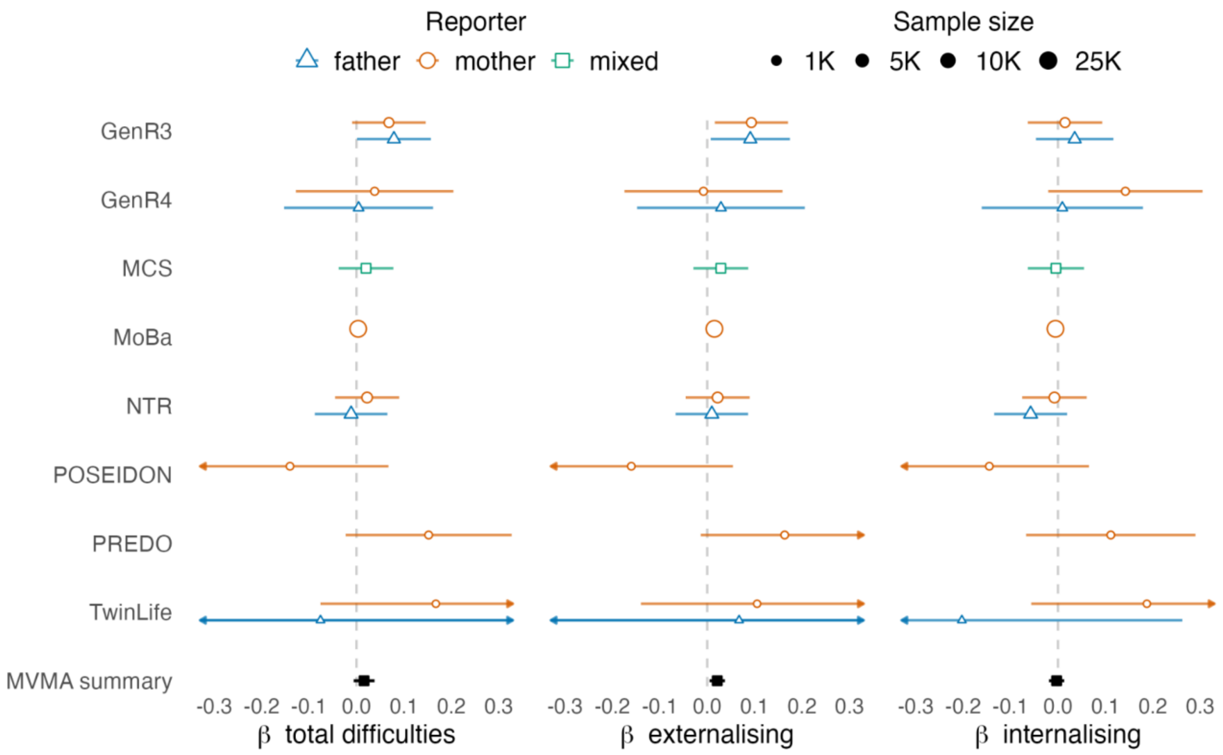


**Figure S2. Cohort- and reporter-specific associations between child Psych-Mean PGS and offspring difficulties at preschool age.**

Note: child Psych-Mean PGS associations with preschool age total, externalising, and internalising difficulties; the multivariate meta-analysis (MVMA) summary indicates the pooled coefficient across cohorts and raters; Psych-Mean = mean score capturing general neuropsychiatric liability.

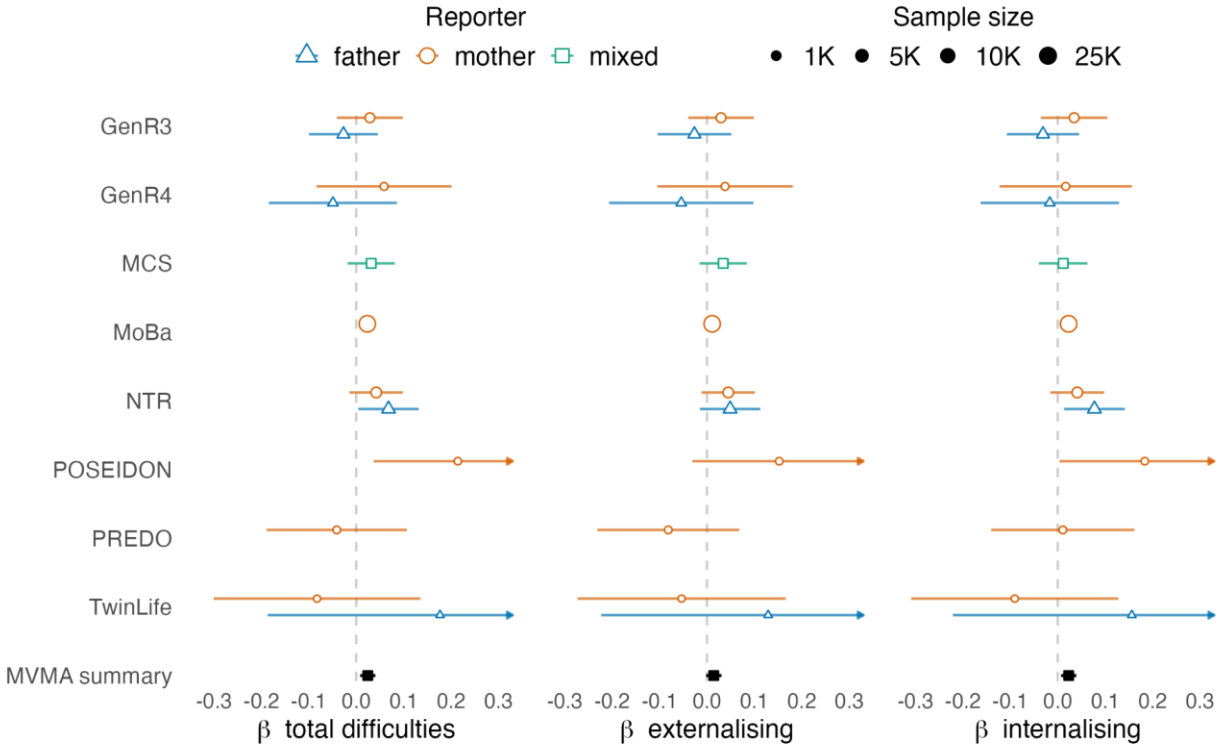


**Figure S3. Cohort- and reporter-specific associations between maternal Psych-Mean PGS and offspring difficulties at preschool age.**

Note: maternal Psych-Mean PGS associations with preschool age total, externalising, and internalising difficulties; paternal PGS associations are not shown since no effects passed multiple testing correction; the multivariate meta-analysis (MVMA) summary indicates the pooled coefficient across cohorts and raters; Psych-Mean = mean score capturing general neuropsychiatric liability.

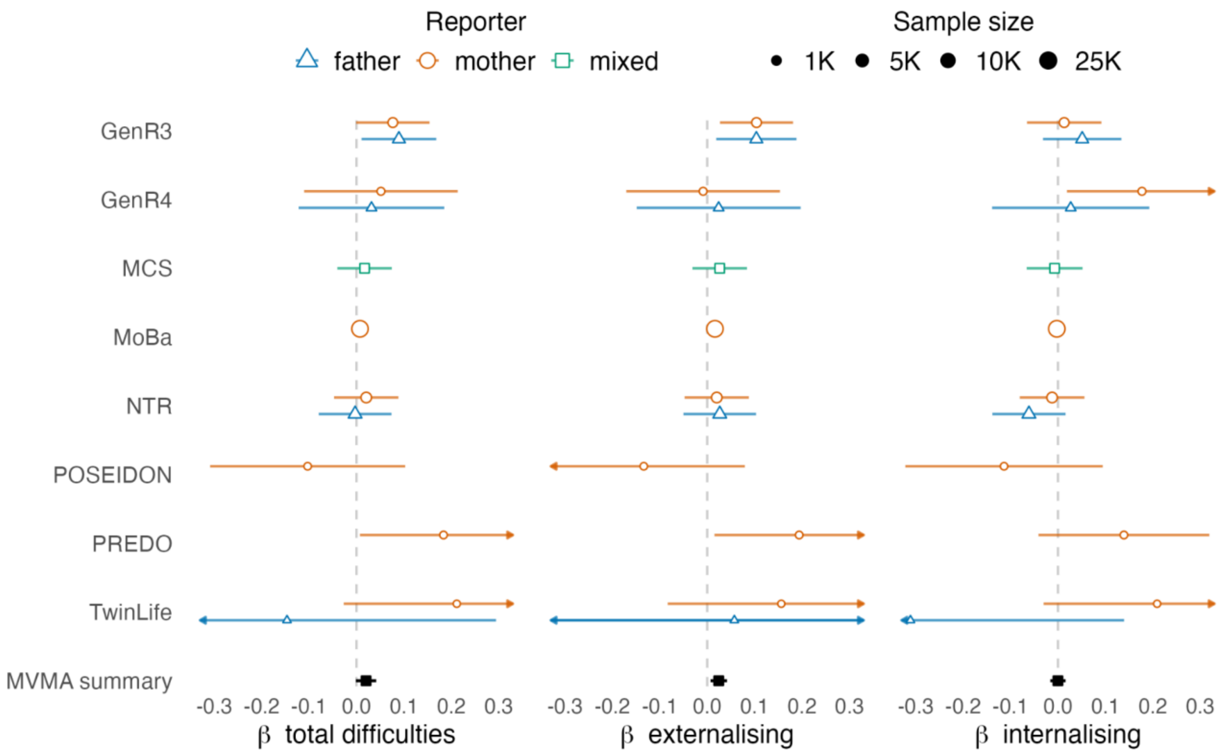


**Figure S4. Cohort- and reporter-specific associations between child Psych-PC1 PGS and offspring difficulties at preschool age.**

Note: child Psych-PC1 PGS associations with preschool age total, externalising, and internalising difficulties; the multivariate meta-analysis (MVMA) summary indicates the pooled coefficient across cohorts and raters; Psych-PC1 = first principal component capturing general neuropsychiatric liability.

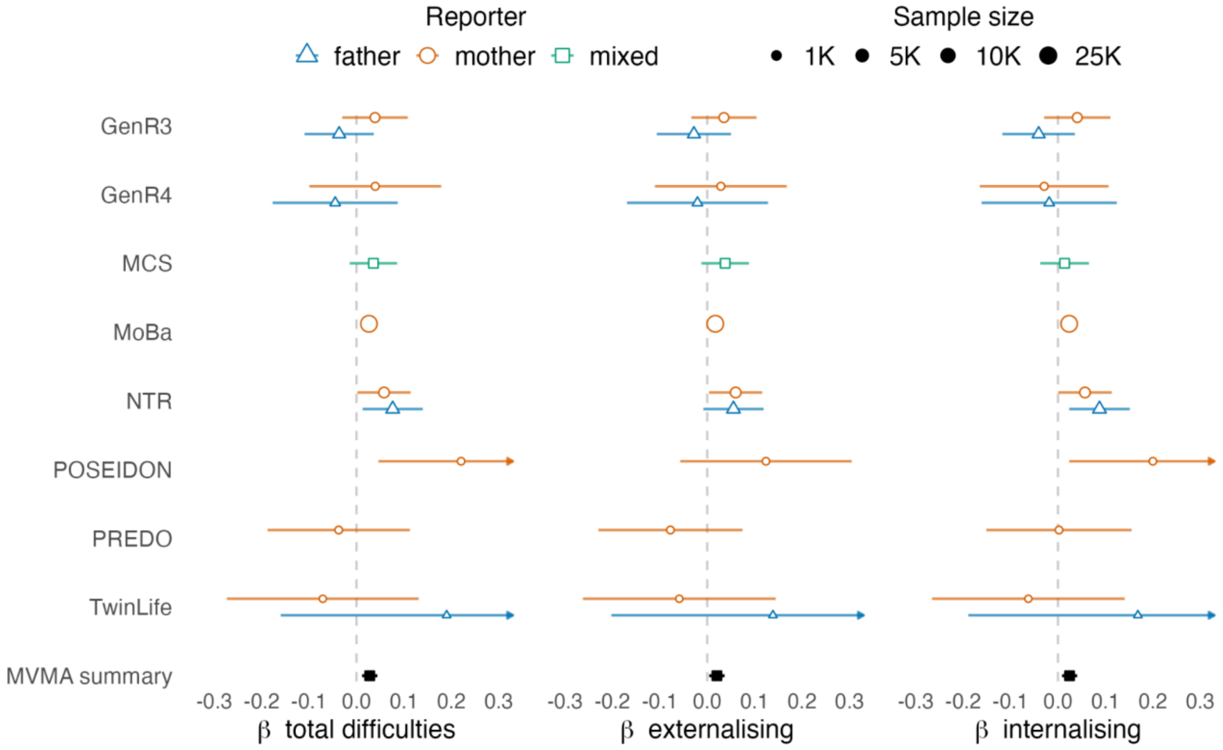


**Figure S5. Cohort- and reporter-specific associations between maternal Psych-PC1 PGS and offspring difficulties at preschool age.**

Note: maternal Psych-PC1 PGS associations with preschool age total, externalising, and internalising difficulties; paternal PGS associations are not shown since no effects passed multiple testing correction; the multivariate meta-analysis (MVMA) summary indicates the pooled coefficient across cohorts and raters; Psych-PC1 = first principal component capturing general neuropsychiatric liability.

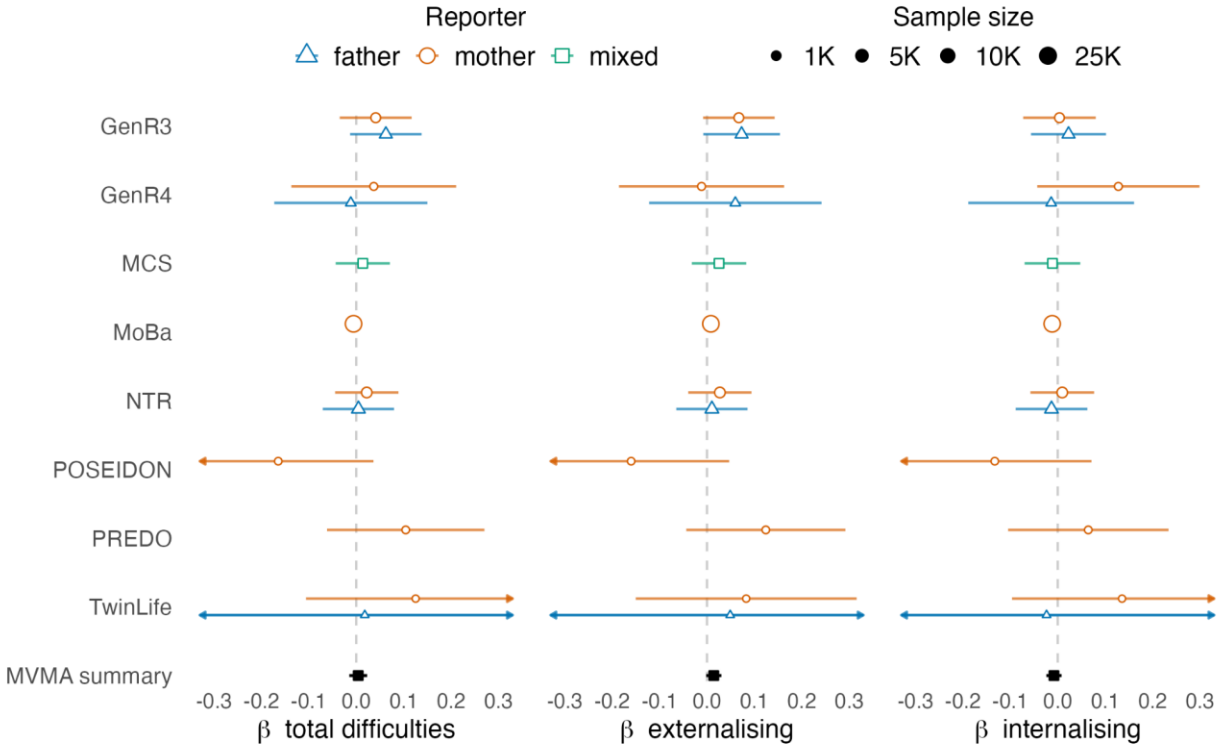


**Figure S6. Cohort- and reporter-specific associations between child CDG PGS and offspring difficulties at preschool age.**

Note: child CDG PGS associations with preschool age total, externalising, and internalising difficulties; the multivariate meta-analysis (MVMA) summary indicates the pooled coefficient across cohorts and raters; CDG = cross-disorder genetic liability.

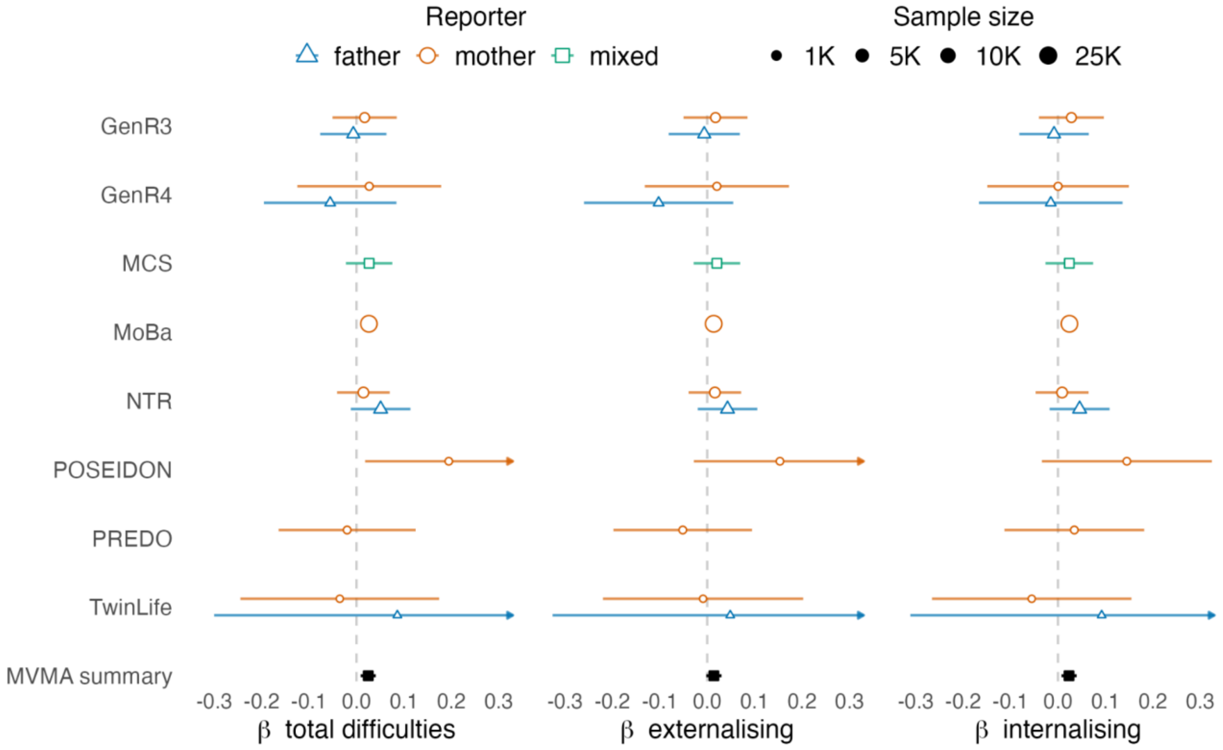


**Figure S7. Cohort- and reporter-specific associations between maternal CDG PGS and offspring difficulties at preschool age.**

Note: maternal CDG PGS associations with preschool age total, externalising, and internalising difficulties; paternal PGS associations are not shown since no effects passed multiple testing correction; the multivariate meta-analysis (MVMA) summary indicates the pooled coefficient across cohorts and raters; CDG = cross-disorder genetic liability.

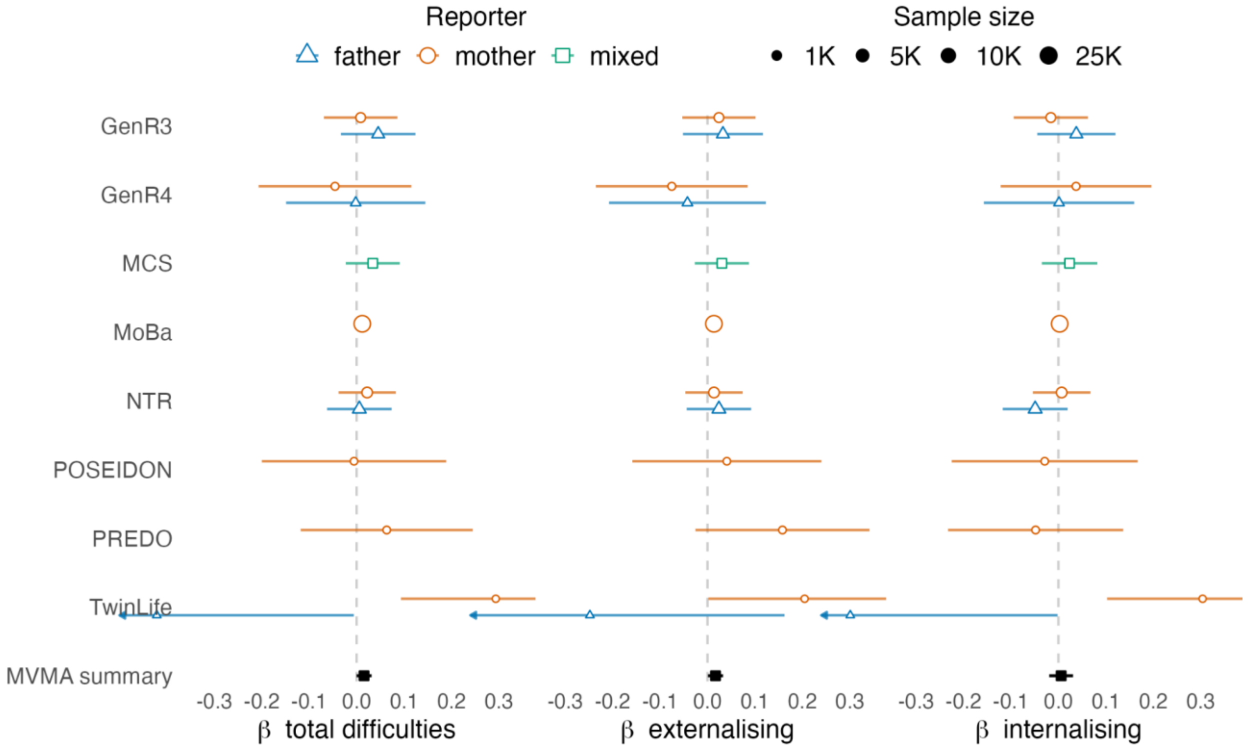


**Figure S8. Cohort- and reporter-specific associations between child MDD PGS and offspring difficulties at preschool age.**

Note: child CDG PGS associations with preschool age total, externalising, and internalising difficulties; the multivariate meta-analysis (MVMA) summary indicates the pooled coefficient across cohorts and raters; MDD = major depressive disorder.

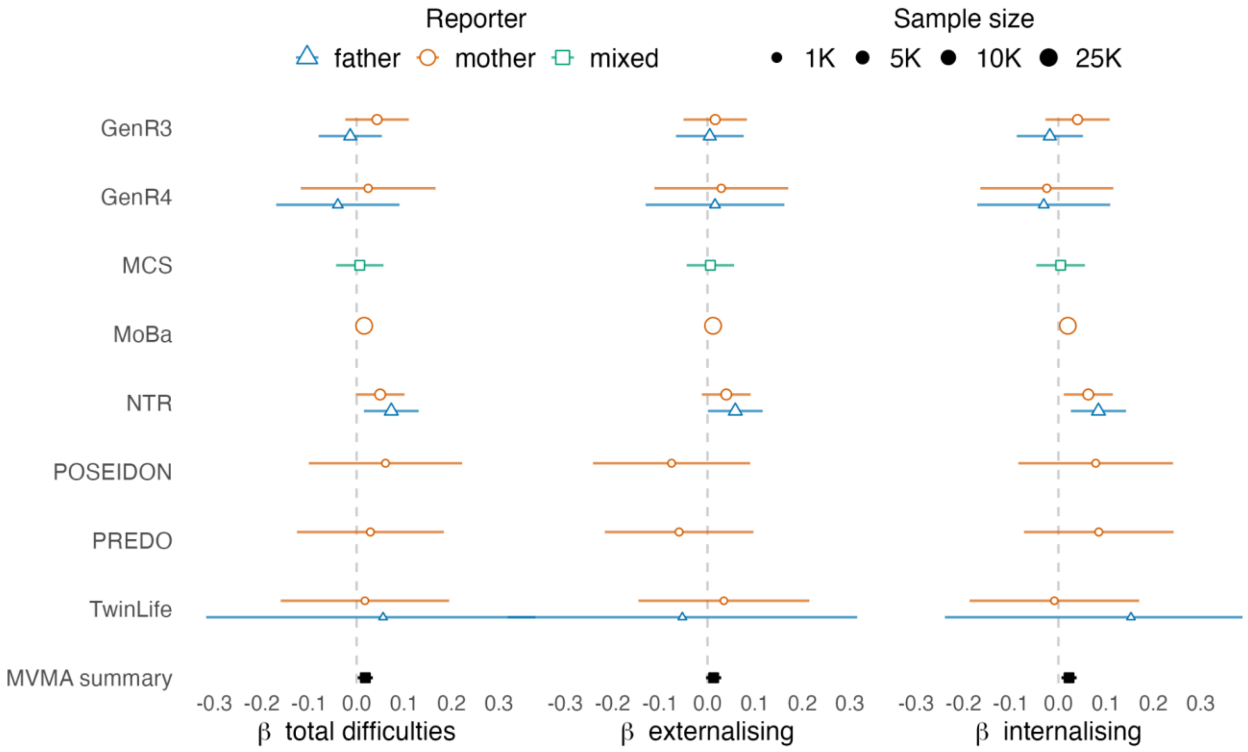


**Figure S9. Cohort- and reporter-specific associations between maternal MDD PGS and offspring difficulties at preschool age.**

Note: maternal MDD PGS associations with preschool age total, externalising, and internalising difficulties; paternal PGS associations are not shown since no effects passed multiple testing correction; the multivariate meta-analysis (MVMA) summary indicates the pooled coefficient across cohorts and raters; MDD = major depressive disorder.

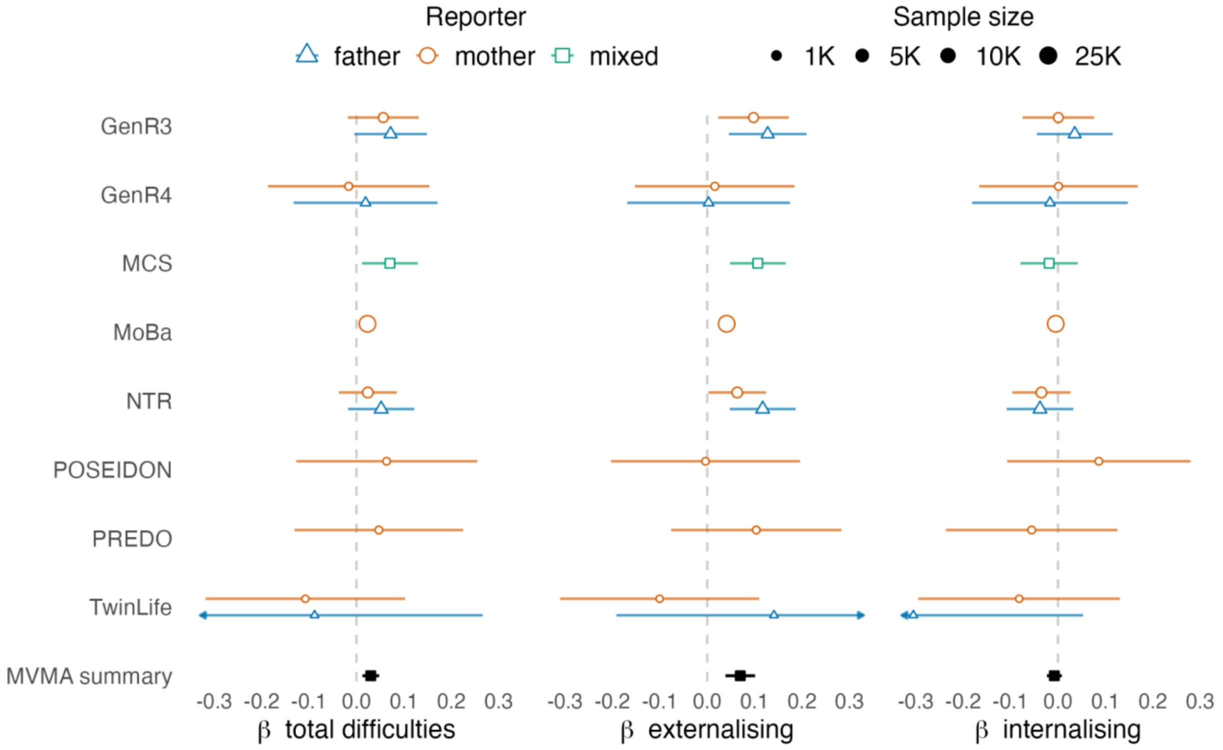


**Figure S10. Cohort- and reporter-specific associations between child ADHD PGS and offspring difficulties at preschool age.**

Note: child ADHD PGS associations with preschool age total, externalising, and internalising difficulties; the multivariate meta-analysis (MVMA) summary indicates the pooled coefficient across cohorts and raters; ADHD = attention-deficit/hyperactivity disorder.

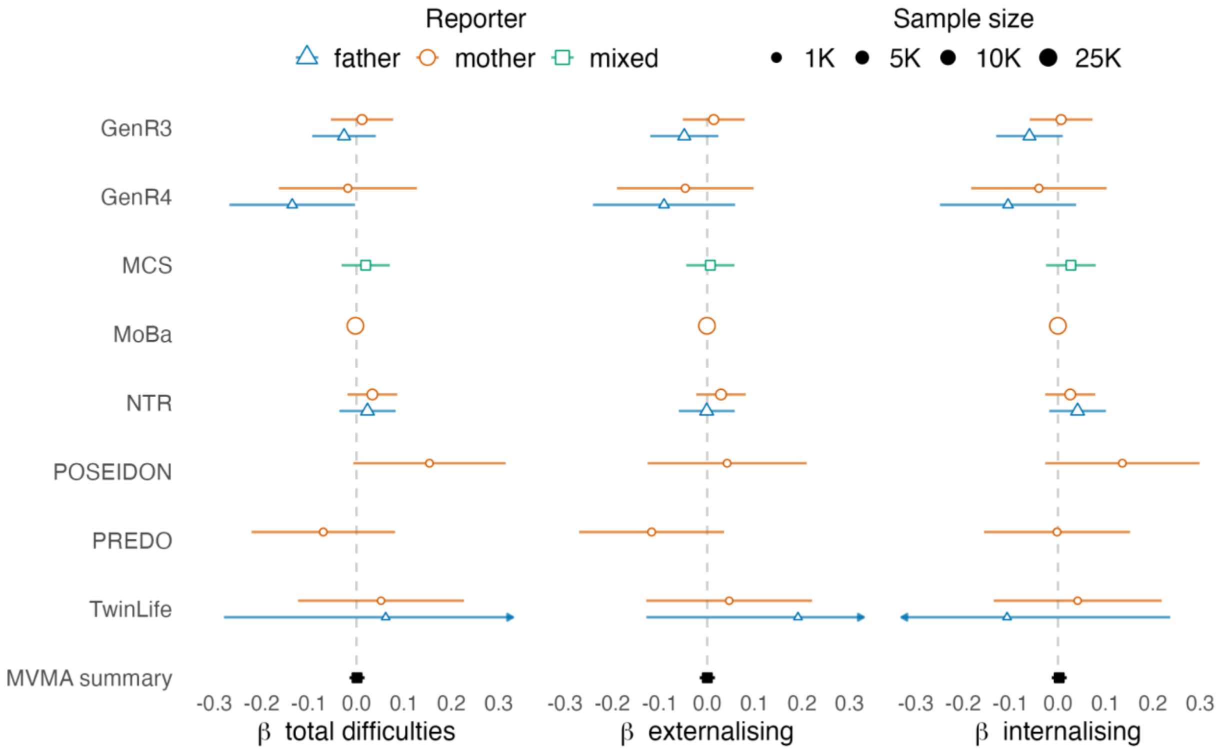


**Figure S11. Cohort- and reporter-specific associations between maternal ADHD PGS and offspring difficulties at preschool age.**

Note: maternal ADHD PGS associations with preschool age total, externalising, and internalising difficulties; paternal PGS associations are not shown since no effects passed multiple testing correction; the multivariate meta-analysis (MVMA) summary indicates the pooled coefficient across cohorts and raters; ADHD = attention-deficit/hyperactivity disorder.

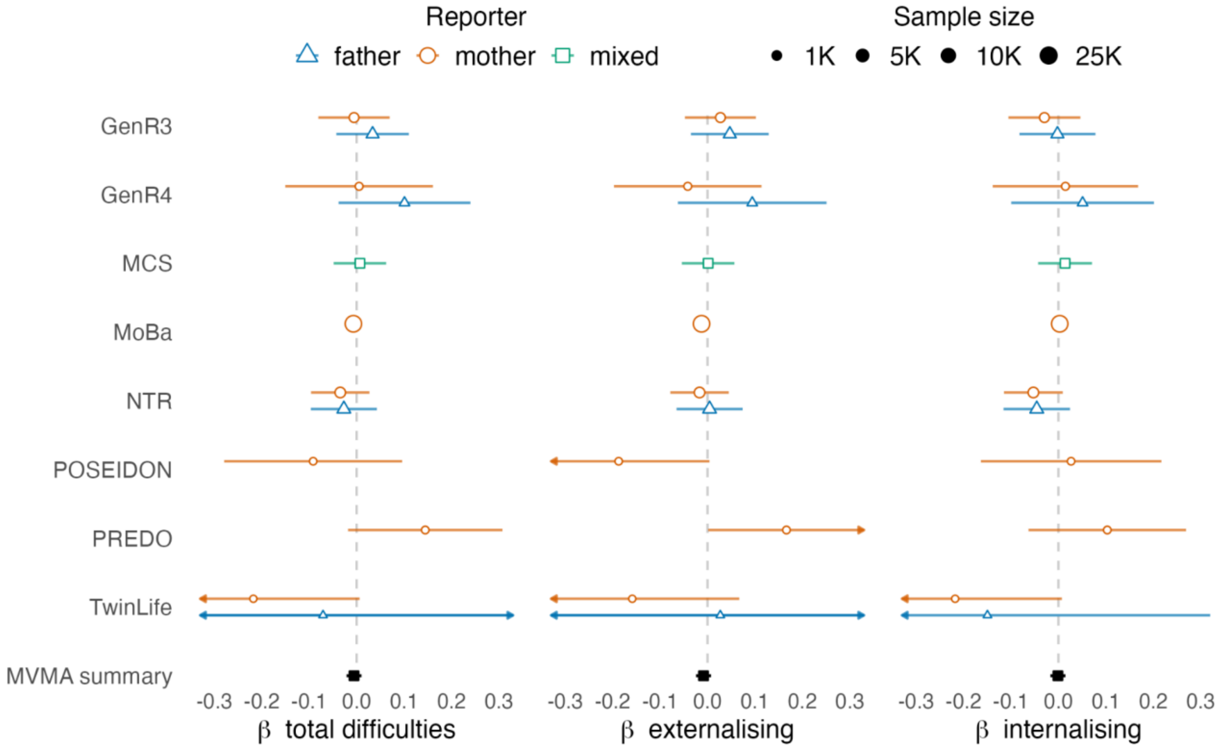


**Figure S12. Cohort- and reporter-specific associations between child ASD PGS and offspring difficulties at preschool age.**

Note: child ASD PGS associations with preschool age total, externalising, and internalising difficulties; the multivariate meta-analysis (MVMA) summary indicates the pooled coefficient across cohorts and raters; ASD = autism spectrum diagnosis.

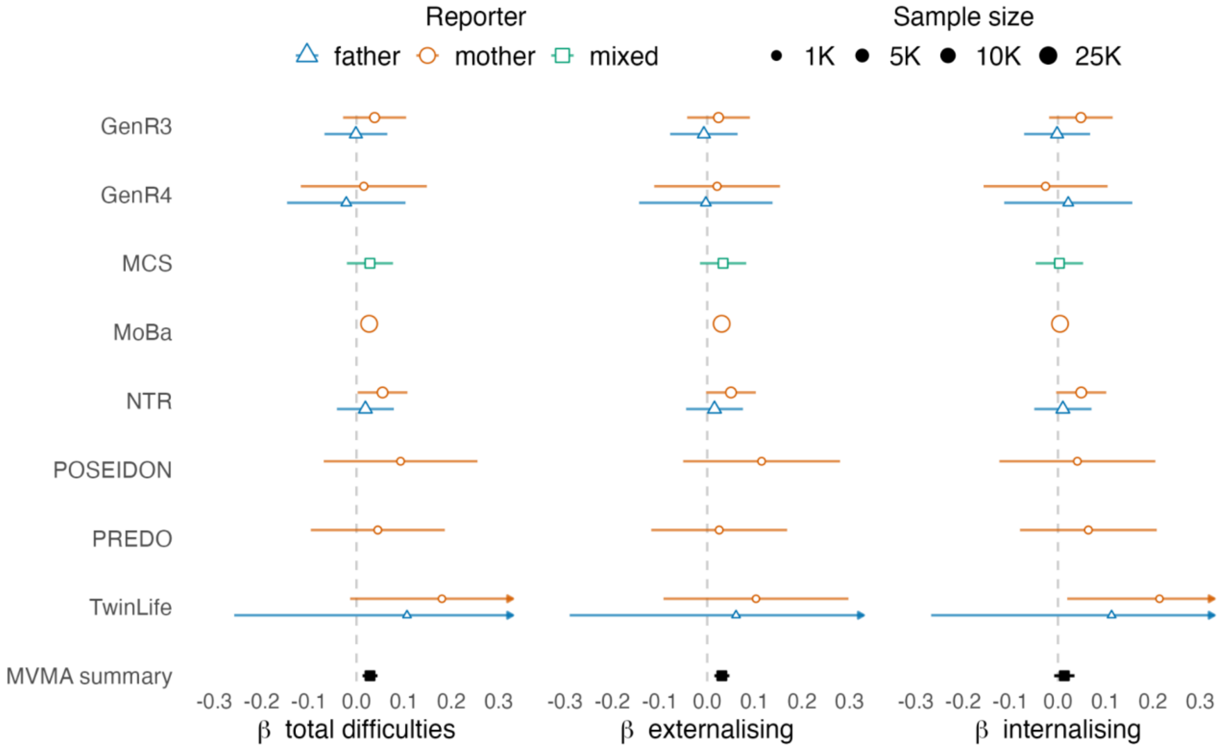


**Figure S13. Cohort- and reporter-specific associations between maternal ASD PGS and offspring difficulties at preschool age.**

Note: maternal ASD PGS associations with preschool age total, externalising, and internalising difficulties; paternal PGS associations are not shown since no effects passed multiple testing correction; the multivariate meta-analysis (MVMA) summary indicates the pooled coefficient across cohorts and raters; ASD = autism spectrum diagnosis.

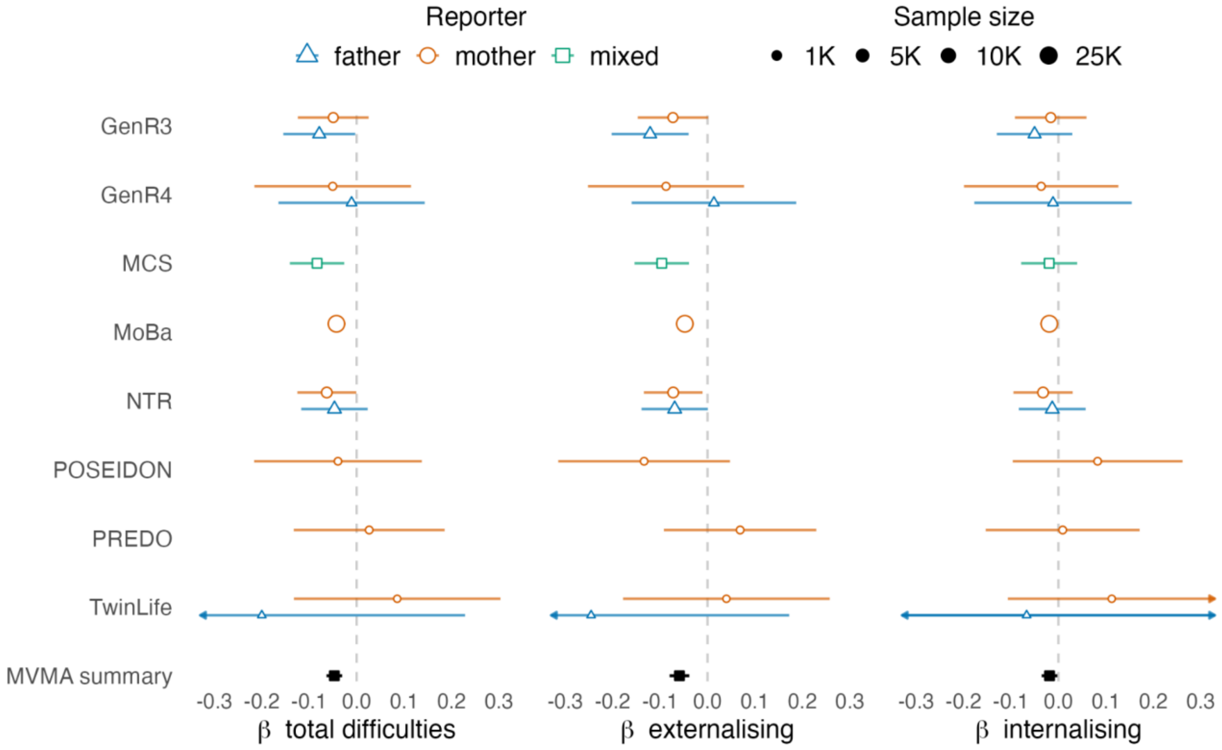


**Figure S14. Cohort- and reporter-specific associations between child EA PGS and offspring difficulties at preschool age.**

Note: child EA PGS associations with preschool age total, externalising, and internalising difficulties; the multivariate meta-analysis (MVMA) summary indicates the pooled coefficient across cohorts and raters; EA = educational attainment.

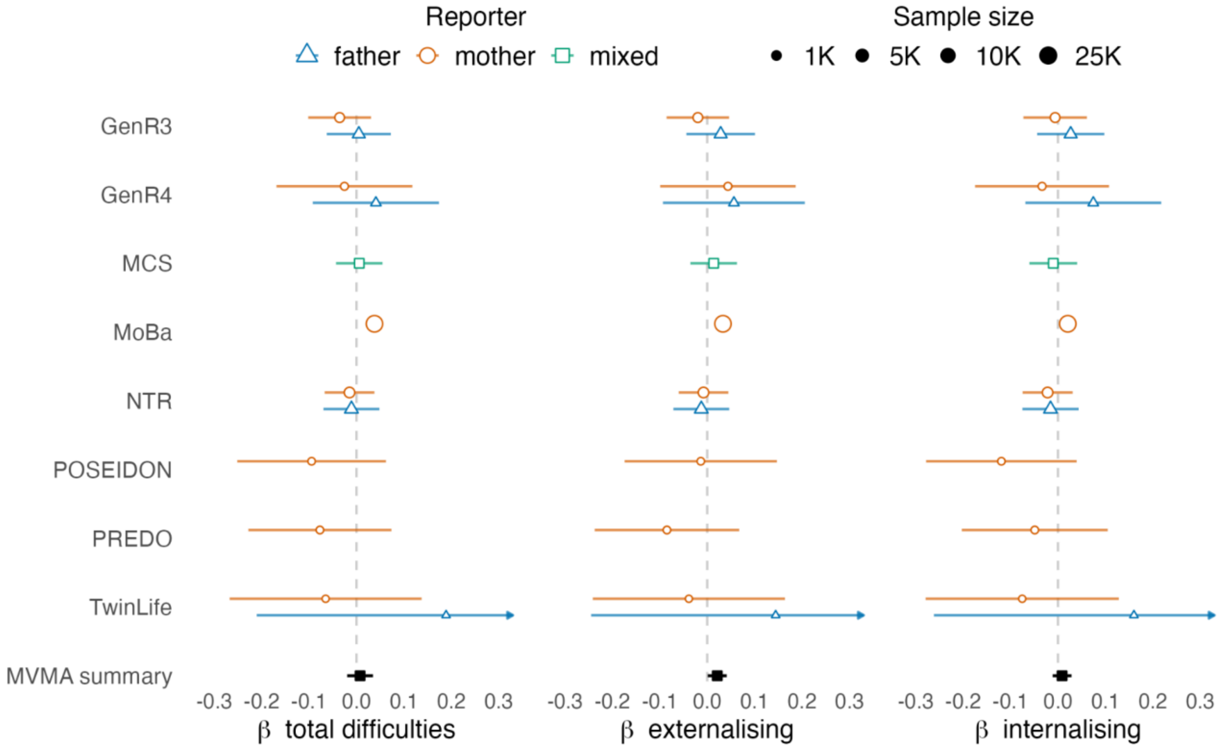


**Figure S15. Cohort- and reporter-specific associations between maternal EA PGS and offspring difficulties at preschool age.**

Note: maternal EA PGS associations with preschool age total, externalising, and internalising difficulties; paternal PGS associations are not shown since no effects passed multiple testing correction; the multivariate meta-analysis (MVMA) summary indicates the pooled coefficient across cohorts and raters; EA = educational attainment.

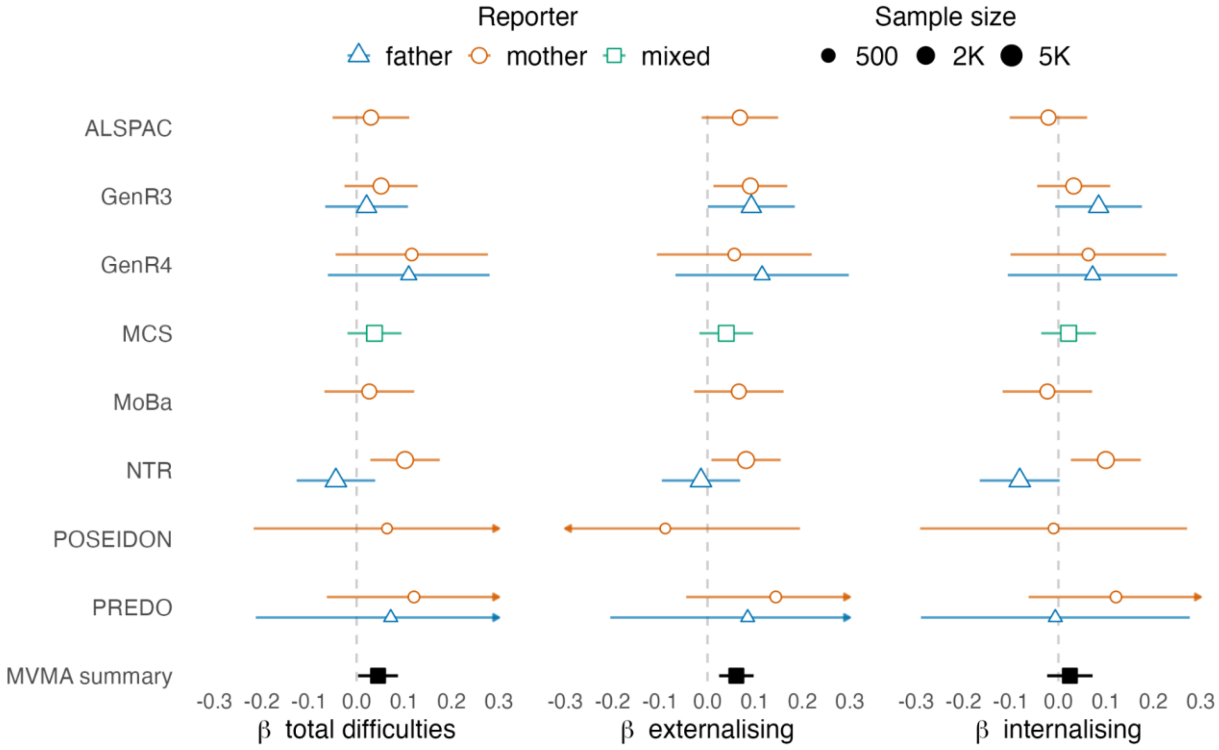


**Figure S16. Cohort- and reporter-specific associations between child Psych-Mean PGS and offspring difficulties at school age.**

Note: child Psych-Mean PGS associations with school age total, externalising, and internalising difficulties; the multivariate meta-analysis (MVMA) summary indicates the pooled coefficient across cohorts and raters; Psych-Mean = mean score capturing general neuropsychiatric liability.

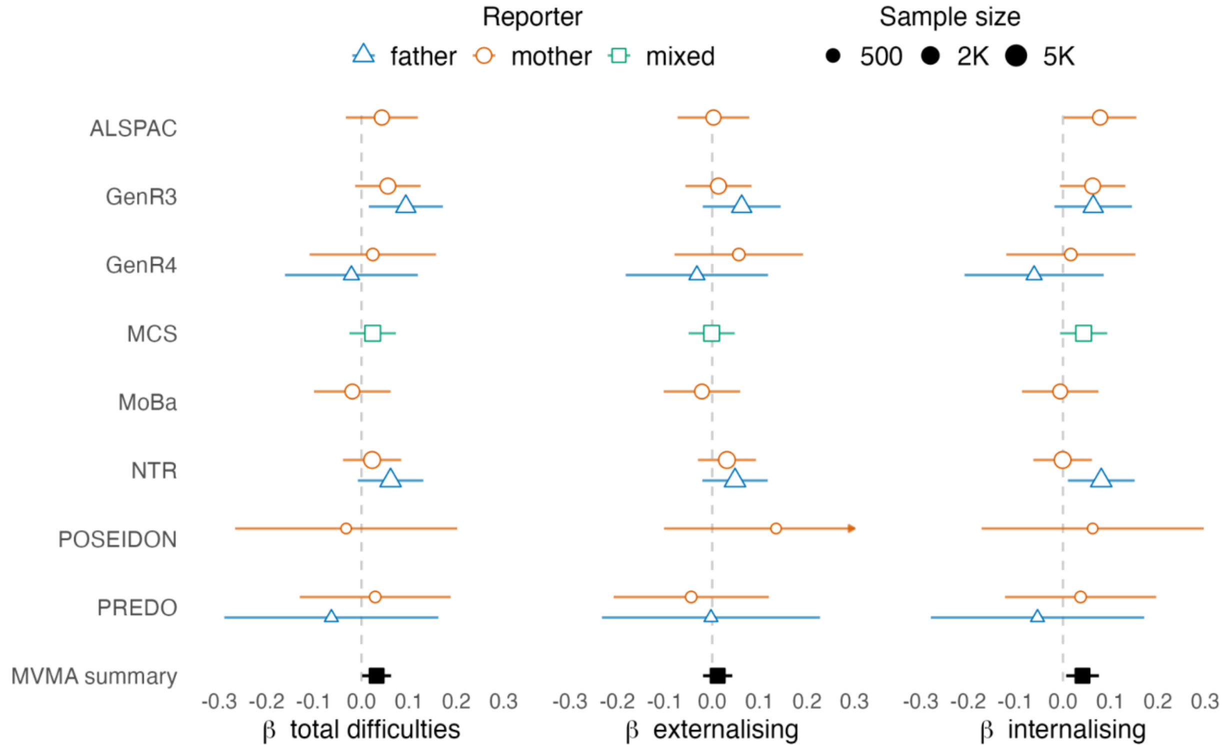


**Figure S17. Cohort- and reporter-specific associations between maternal Psych-Mean PGS and offspring difficulties at school age.**

Note: maternal Psych-Mean PGS associations with school age total, externalising, and internalising difficulties; paternal PGS associations are not shown since no effects passed multiple testing correction; the multivariate meta-analysis (MVMA) summary indicates the pooled coefficient across cohorts and raters; Psych-Mean = mean score capturing general neuropsychiatric liability.

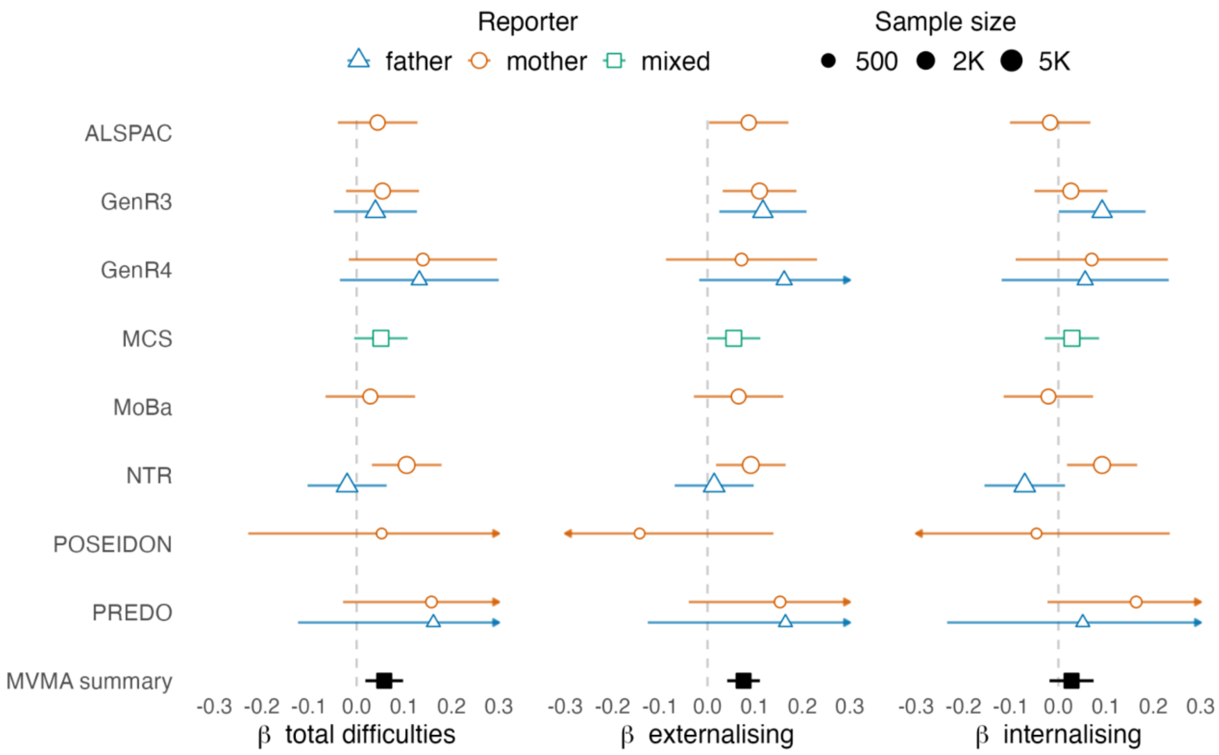


**Figure S18. Cohort- and reporter-specific associations between child Psych-PC1 PGS and offspring difficulties at school age.**

Note: child Psych-PC1 PGS associations with school age total, externalising, and internalising difficulties; the multivariate meta-analysis (MVMA) summary indicates the pooled coefficient across cohorts and raters; Psych-PC1 = first principal component capturing general neuropsychiatric liability.

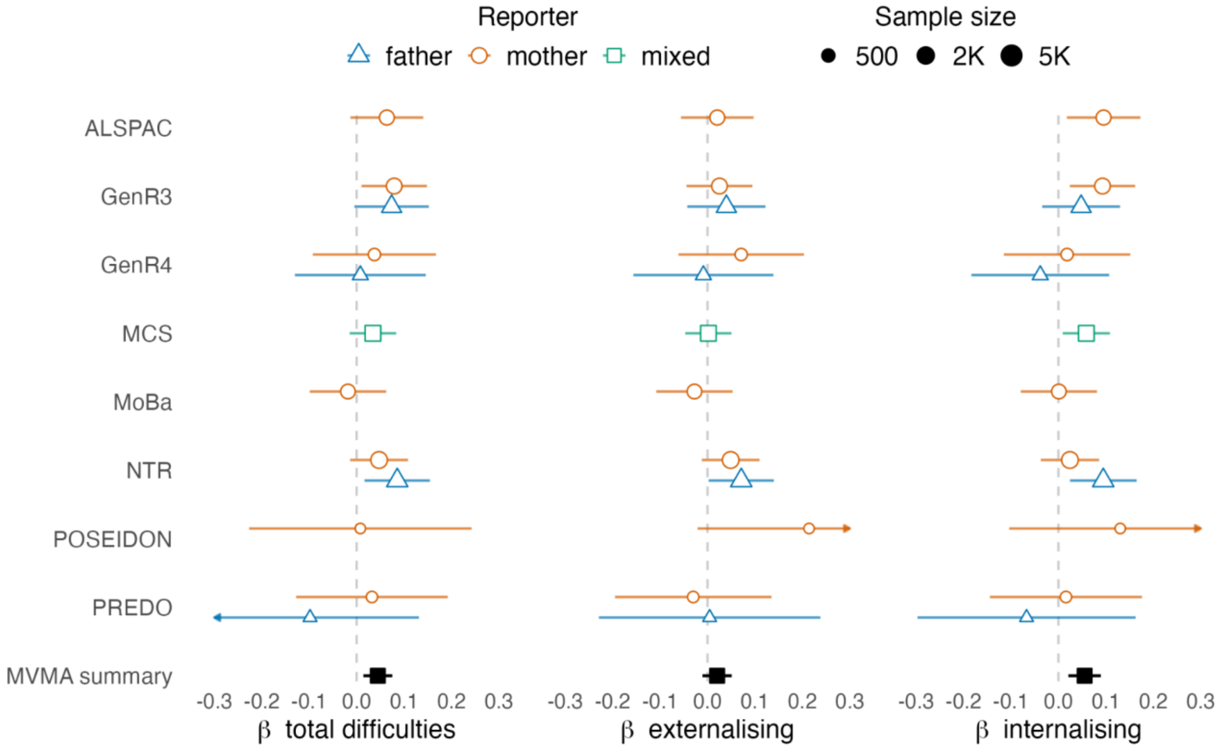


**Figure S19. Cohort- and reporter-specific associations between maternal Psych-PC1 PGS and offspring difficulties at school age.**

Note: maternal Psych-PC1 PGS associations with school age total, externalising, and internalising difficulties; the multivariate meta-analysis (MVMA) summary indicates the pooled coefficient across cohorts and raters; Psych-PC1 = first principal component capturing general neuropsychiatric liability.

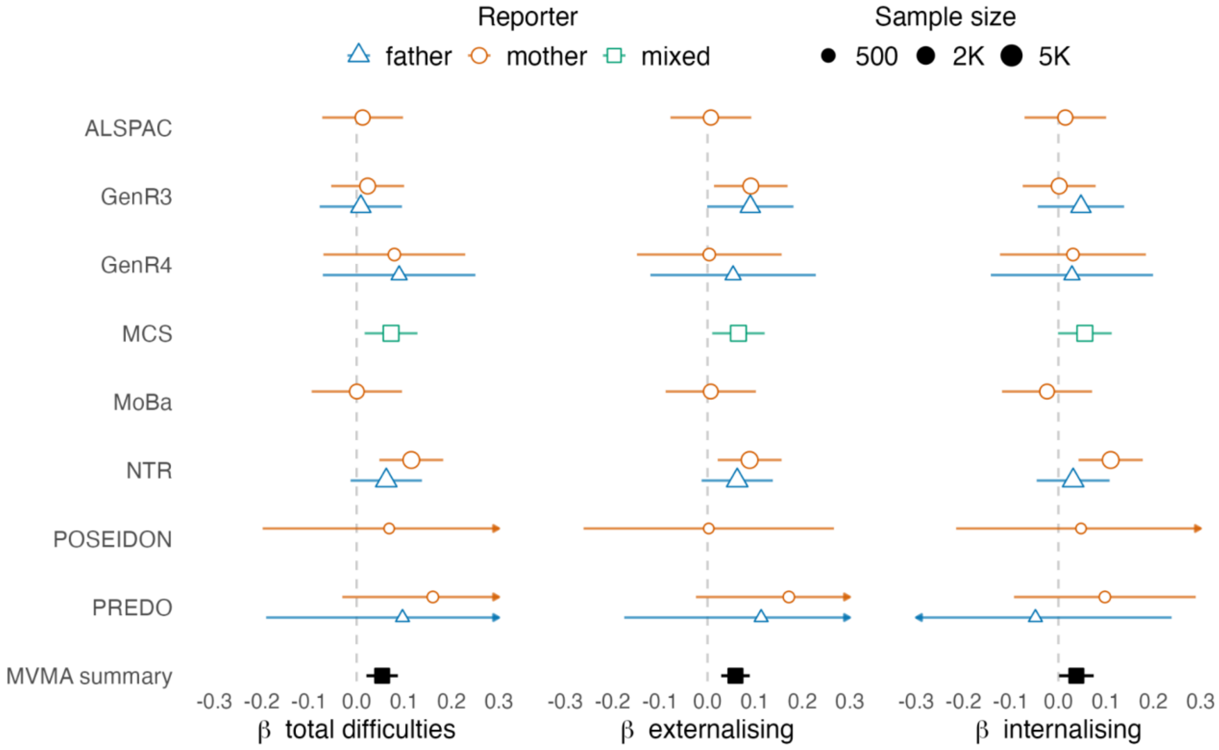


**Figure S20. Cohort- and reporter-specific associations between child MDD PGS and offspring difficulties at school age.**

Note: child MDD PGS associations with school age total, externalising, and internalising difficulties (conditional on parental PGS); the multivariate meta-analysis (MVMA) summary indicates the pooled coefficient across cohorts and raters; MDD = major depressive disorder.

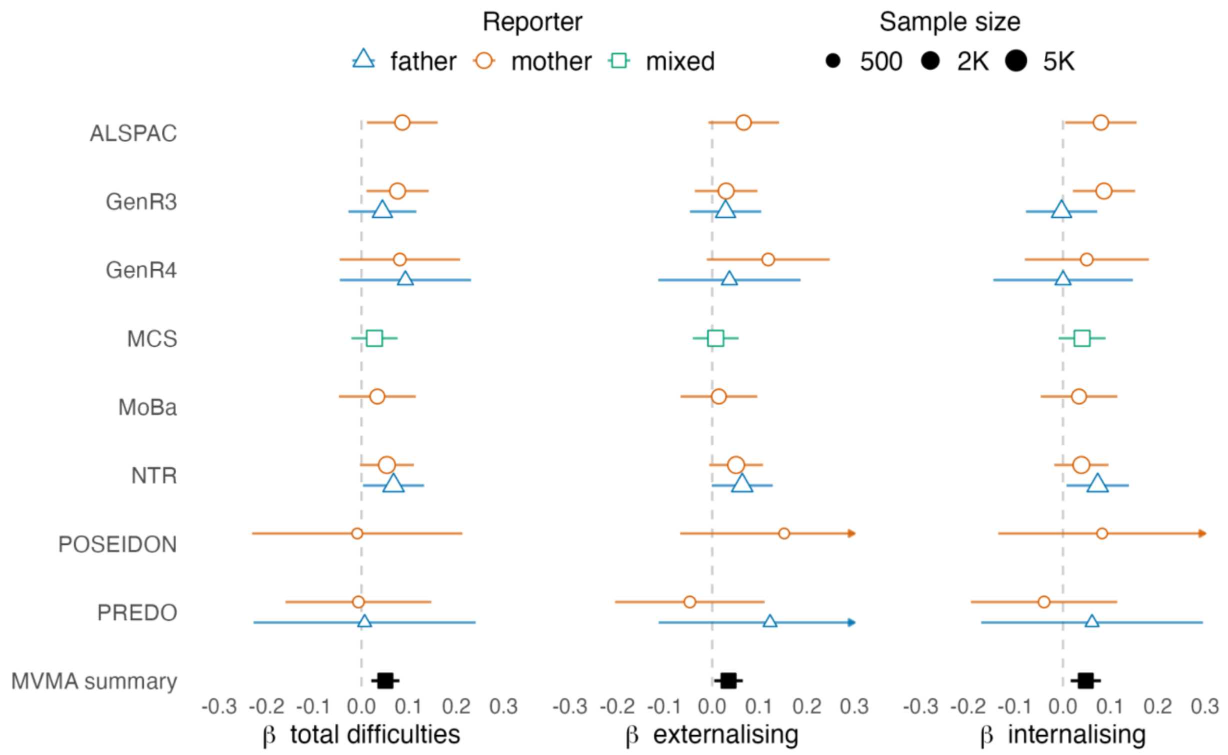


**Figure S21. Cohort- and reporter-specific associations between maternal MDD PGS and offspring difficulties at school age.**

Note: maternal MDD PGS associations with school age total, externalising, and internalising difficulties; paternal PGS associations are not shown since no effects passed multiple testing correction; the multivariate meta-analysis (MVMA) summary indicates the pooled coefficient across cohorts and raters; MDD = major depressive disorder.

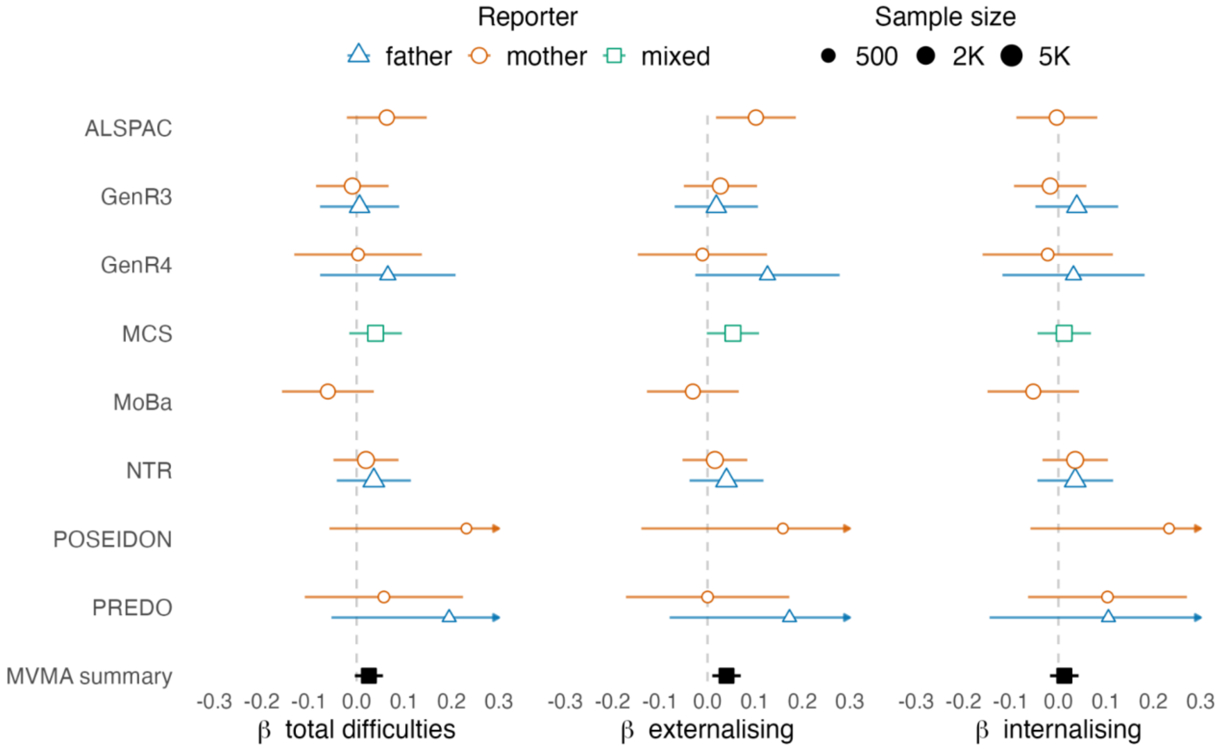


**Figure S22. Cohort- and reporter-specific associations between child PPD PGS and offspring difficulties at school age.**

Note: child PGS associations with school age total, externalising, and internalising difficulties (conditional on parental PGS); the multivariate meta-analysis (MVMA) summary indicates the pooled coefficient across cohorts and raters; PPD = postpartum depressive disorder.

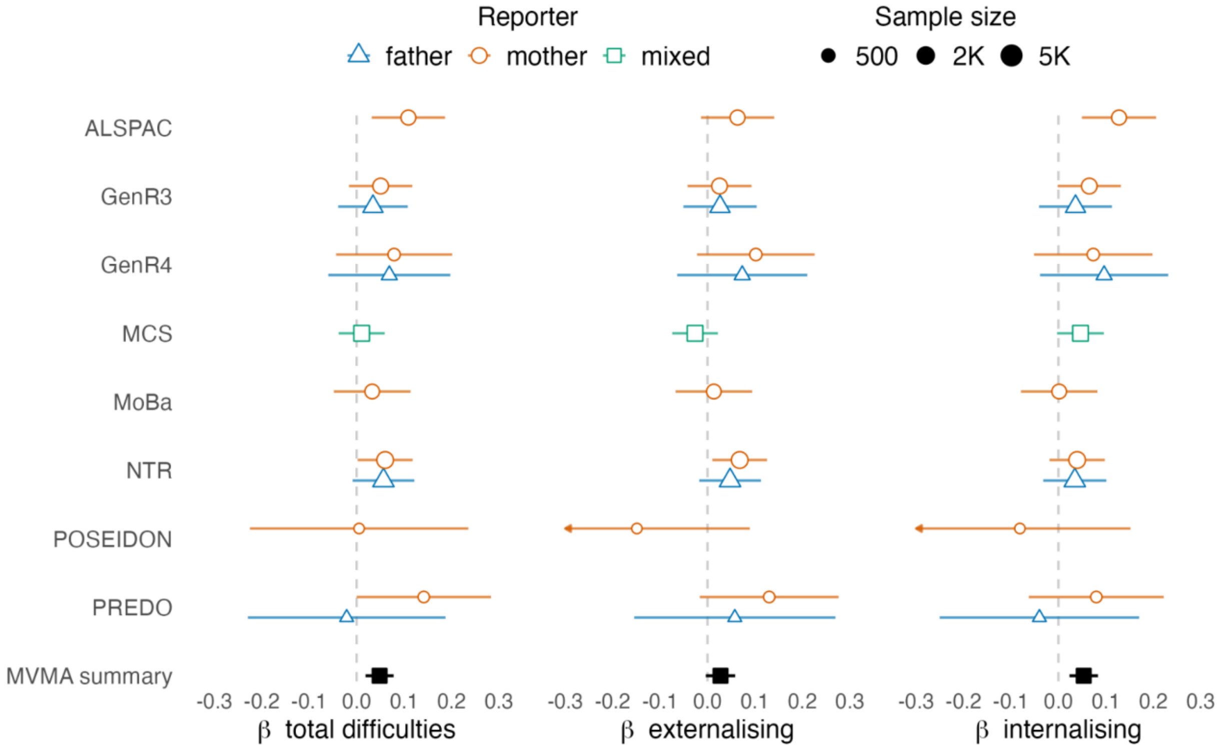


**Figure S23. Cohort- and reporter-specific associations between maternal PPD PGS and offspring difficulties at school age.**

Note: maternal PPD PGS associations with school age total, externalising, and internalising difficulties; paternal PGS associations are not shown since no effects passed multiple testing correction; the multivariate meta-analysis (MVMA) summary indicates the pooled coefficient across cohorts and raters; PPD = postpartum depressive disorder.

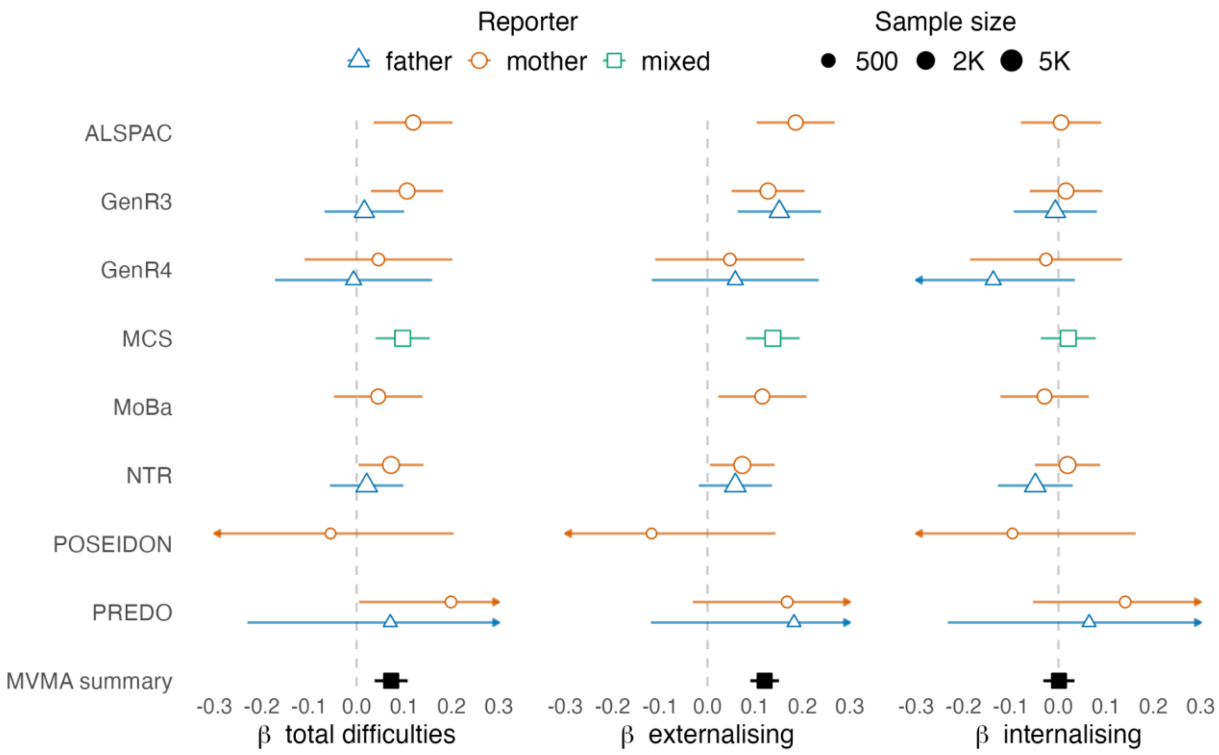


**Figure S24. Cohort- and reporter-specific associations between child ADHD PGS and offspring difficulties at school age.**

Note: child ADHD PGS associations with school age total, externalising, and internalising difficulties (conditional on parental PGS); the multivariate meta-analysis (MVMA) summary indicates the pooled coefficient across cohorts and raters; ADHD = attention-deficit/hyperactivity disorder.

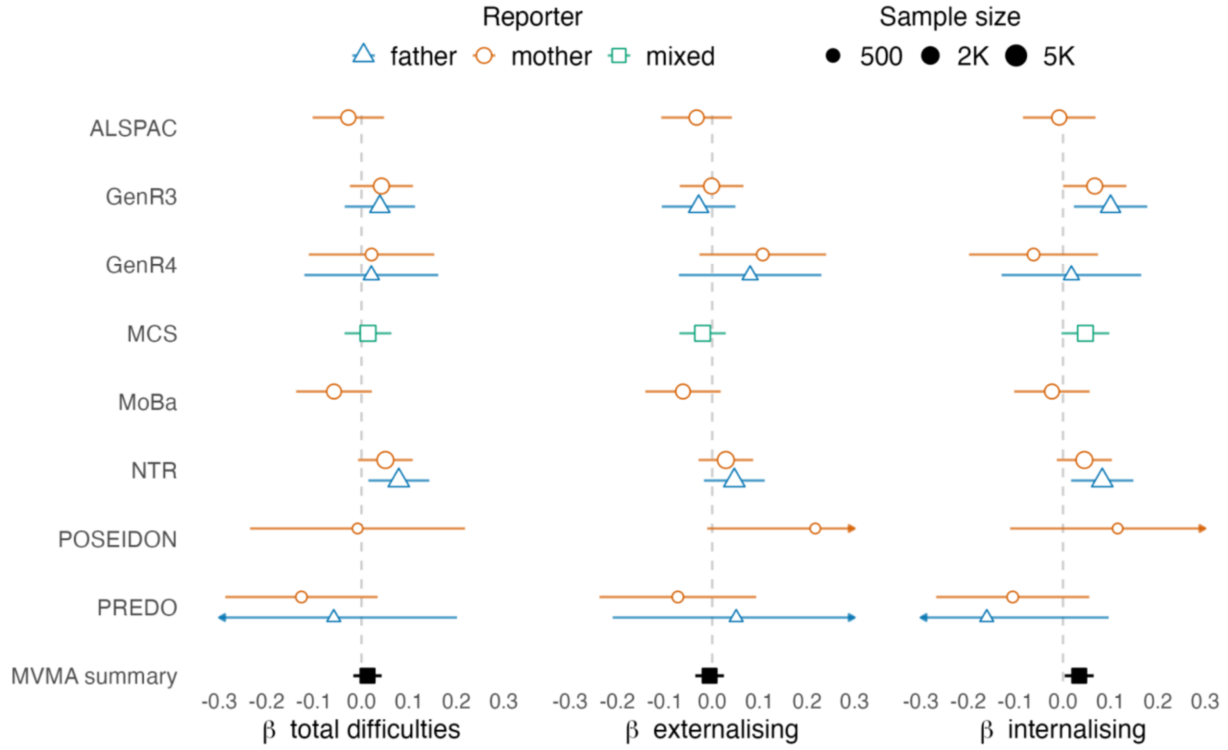


**Figure S25. Cohort- and reporter-specific associations between maternal ADHD PGS and offspring difficulties at school age.**

Note: maternal ADHD PGS associations with school age total, externalising, and internalising difficulties; paternal PGS associations are not shown since no effects passed multiple testing correction; the multivariate meta-analysis (MVMA) summary indicates the pooled coefficient across cohorts and raters; ADHD = attention-deficit/hyperactivity disorder.

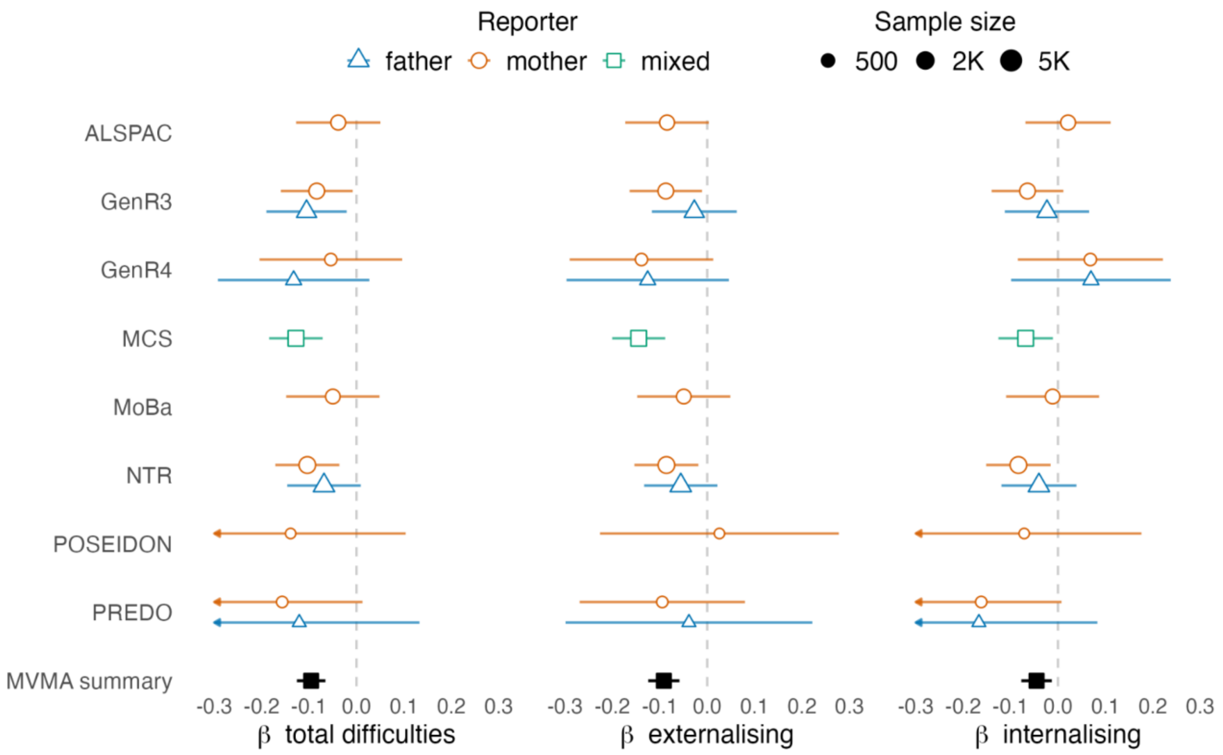


**Figure S26. Cohort- and reporter-specific associations between child EA PGS and offspring difficulties at school age.**

Note: child EA PGS associations with school age total, externalising, and internalising difficulties (conditional on parental PGS); the multivariate meta-analysis (MVMA) summary indicates the pooled coefficient across cohorts and raters; EA = educational attainment.

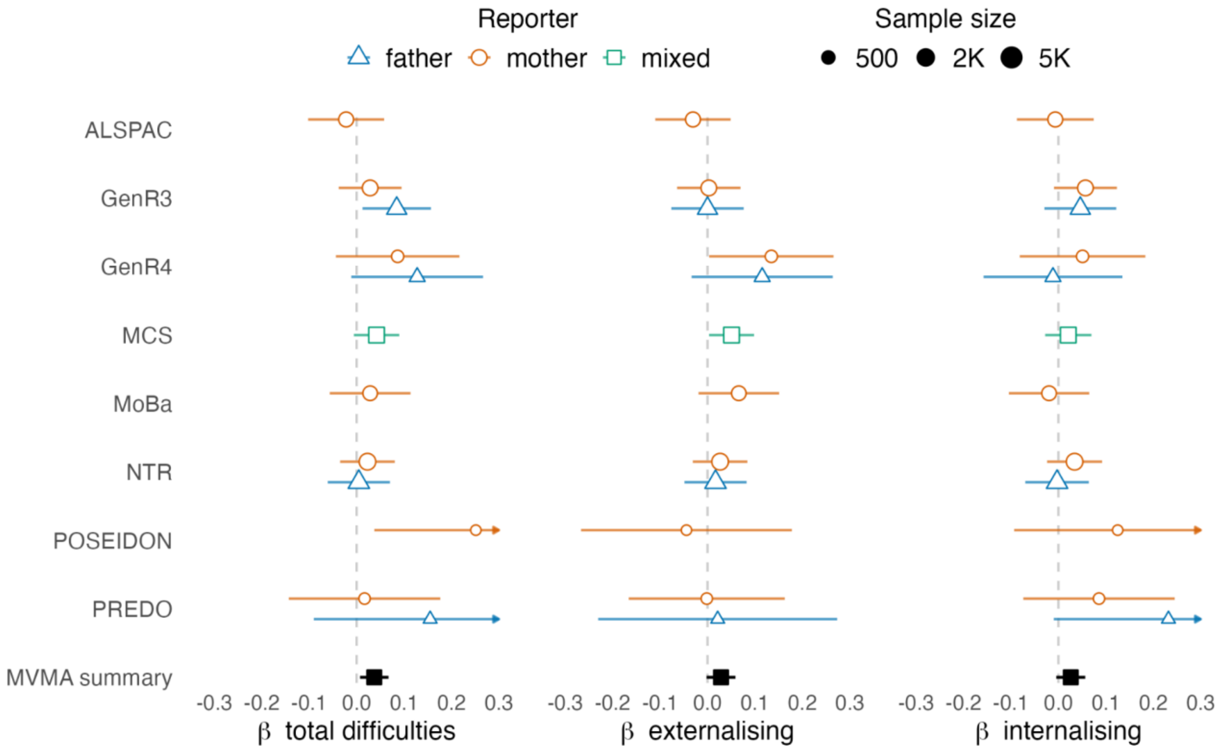


**Figure S27. Cohort- and reporter-specific associations between maternal EA PGS and offspring difficulties at school age.**

Note: maternal EA PGS associations with school age total, externalising, and internalising difficulties; paternal PGS associations are not shown since no effects passed multiple testing correction; the multivariate meta-analysis (MVMA) summary indicates the pooled coefficient across cohorts and raters; EA = educational attainment.
